# Time-Specific Associations of Wearable, Sensor-Based Cardiovascular and Behavioral Readouts with Disease Phenotypes in the Outpatient Setting of the Chronic Renal Insufficiency Cohort (CRIC)

**DOI:** 10.1101/2022.01.09.22268966

**Authors:** Nicholas F. Lahens, Mahboob Rahman, Jordana B. Cohen, Debbie L. Cohen, Jing Chen, Matthew R. Weir, Harold I. Feldman, Gregory R. Grant, Raymond R. Townsend, Carsten Skarke

## Abstract

Patients with chronic kidney disease (CKD) are at risk of developing cardiovascular disease. To facilitate out-of-clinic evaluation, we piloted wearable device-based analysis of heart rate variability and behavioral readouts in patients with CKD participating in the Chronic Renal Insufficiency Cohort and (n=49) controls. Time-specific partitioning of HRV readouts indicate higher parasympathetic nervous activity during the night (mean RR at night 14.4±1.9 ms versus 12.8±2.1 ms during active hours; n=47, ANOVA q=0.001). The α2 long-term fluctuations in the detrended fluctuation analysis, a parameter predictive of cardiovascular mortality, significantly differentiated between diabetic and non-diabetic patients (prominent at night with 0.58±0.2 versus 0.45±0.12, respectively, adj. p=0.004). Both diabetic and nondiabetic CKD patients showed loss of rhythmic organization compared to controls, with diabetic CKD patients exhibiting deconsolidation of peak phases between their activity and SDNN (standard deviation of interbeat intervals) rhythms (mean phase difference CKD 8.3h, CKD/T2DM 4h, controls 6.8h). This work provides a roadmap toward deriving actionable clinical insights from the data collected by wearable devices outside of highly controlled clinical environments.

## Introduction

Cardiovascular disease is the leading cause of morbidity and mortality in patients with chronic kidney disease (CKD). Heart failure (CHF) is the most common non-fatal CV morbidity seen in patients with CKD. Data collected from the Chronic Renal Insufficiency Cohort (CRIC) study have identified a number of factors associated with the development of CHF. These risk factors include arterial stiffness ^1^, elevated serum bicarbonate ^2^, increased pulse pressure (in CKD stage 4 and 5) ^3^, and elevated levels of troponin T and NT-proBNP, as well as left ventricular hypertrophy on echocardiography ^4^. To date, these analyses have relied exclusively on data obtained through evaluations carried out on CRIC participants while in-center at yearly visits. However, the ability to measure biometric signals in ambulatory settings, and its acceptability in industry and academia, is expanding rapidly ^5^, and there are now several easy-to-use, wearable devices that record such signals. Furthermore, as health systems increase their use of telemedicine in response to the ongoing SARS-CoV-2 pandemic, remote sensing devices can help bridge a diagnostic gap between clinical settings and patients’ homes. Work in the preventive healthcare area currently employs biometric monitoring in the outpatient evaluation and management of heart failure ^6^. However, there is very little known about the prognostic value of biometric monitoring in CKD ^7^. Given the increasing emphasis on out-of-clinic health assessments, we conducted a biometric pilot study to evaluate cardiovascular function and physical activity in CRIC study participants using a single wearable device, the Zephyr BioPatch. The BioPatch is a 2-lead cardiac monitoring device mounted on a patient’s sternum using adhesive tape patches.

We chose heart rate variability (HRV) as the parameter of interest to assess cardiovascular function as a prior report from the CRIC study showed an association between HRV and risk of mortality ^8^. In this previous study, Drawz *et al*. derived HRV data from 10 seconds of QRS complexes from a 12 lead EKG from participants at rest during a clinic visit. HRV is defined as a group of parameters derived from waveform EKG tracings that evaluate the intervals between consecutive normal heart beats as a proxy for autonomic nervous system function^9^. In addition, arterial baroreflex function also influences HRV. Existing literature shows that cardiac autonomic function in patients with coronary artery disease (CAD) ^10^, diabetes mellitus ^11^, existing CHF ^12^, and increasing age ^13^ differs systematically from that of non-CAD patients. Both heart rate and its variability demonstrate robust endogenous circadian rhythms ^14^ with sexually dimorphic effect sizes ^15^, an age-dependent decrease ^16^, and possibly seasonal differences ^17^. Notably, previous experiments showed subjects exhibit a reduced HRV under conditions of an evoked inflammatory response ^18,19^.

Progression of CKD tracks with a decrease in physical function in patients under pre-dialysis conditions ^20^ and is associated with all-cause mortality ^21^ and reduced quality of life ^22^. The National Health and Nutrition Examination Survey (NHANES) III estimated that physical inactivity was more prevalent among CKD patients (28%), than among non-CKD controls (13.5%); however, a limitation is the insignificant discriminatory impact on mortality in this questionnaire-based observational study ^23^. A Cochrane review emphasizes the beneficial effects of regular increased physical activity on risk factors in patients with CKD ^24^ which led to refined exercise recommendations for this patient population ^25,26^. However, data on the level of physical activity maintained in the home environment of CKD patients are not routinely assessed.

The aim of the present two-center pilot study within the Chronic Renal Insufficiency Cohort (CRIC) and an external reference cohort of healthy controls was to determine acceptance of wearable biosensor technology among participants and to discern whether data streams associate with disease phenotypes, despite the noise introduced by activities of daily life, to generate clinical insight.

## Results

### Study participants

The CRIC Study is an observational study that examined risk factors for progression of chronic renal insufficiency (CRI) and cardiovascular disease (CVD) among CRI patients. For this subprotocol, the University of Pennsylvania and University Hospitals of Cleveland centers recruited n=39 patient volunteers from the CRIC cohort with the following characteristics: 14 females (36%), 63.9±7.7 years of age, 17 African-American and 22 Caucasian (Figure S 1, Table S 1). Comorbidities included hypertension (33/39), type 2 diabetes mellitus (18/39; T2DM), asthma (8/39), COPD (6/39) and arthritis (3/39); but the degree of heterogeneity driven by medical history, prescription drugs, and laboratory tests is much higher, as listed in Table S 3. We leveraged the near balanced distribution of T2DM among CKD patients to address the hypothesis that biometric signals can differentiate between patients comorbid for CKD and T2DM (n=18; labeled CKD/T2DM for the remainder of this manuscript), compared to CKD patients (n=21) and controls (n=10, Figure S 1). In clinical and laboratory assessments, CKD/T2DM patients (n=21, eGFR of 55.1m^2^±26.7m^2^ and HbA1c of 7.4%±1.2%) displayed lower GFR compared to CKD patients (n=18, eGFR of 59.9 m^2^±22.9 m^2^ and HbA1c of 5.6%±0.3%), (Figure S 3, Table S 2). On average, controls were younger than the cases (7 females, 30.4±10.5 years of age, 8 Caucasian and 2 Asian, Table S 1), an approach that increased the likelihood to detect differences between cases and controls in cardiovascular-behavioral outputs.

### High degree of compliance and data quality for biometric data streams

On average, CRIC participants wore the device for 49±12.8 hours and healthy controls for 48.3±6.7 hours. Variable wear times resulted from charging the device in the CRIC cohort, switching between devices in the controls, or personal reasons (e.g. shower breaks) in both cohorts (Figure S 2). Data from two CRIC participants showed a low level of compliance (wear time of 1.9 and 9.1 hours). As a result, we excluded these two subjects from all BioPatch analyses. Data from the majority of participants (48/49) showed a high proportion of reliable heart rate (HR) readings (78.1±16.8%), defined as HR confidence >20% provided in the BioPatch readouts. High correlations within subjects for time and frequency domain readouts underscore internal consistency for these time series. This is evident, for example, between SDNN compared to total power (R^2^≥0.6) and powers of HF (R^2^≥0.73) as well as LF (R^2^≥0.44); or RMSSD compared to SD1 (R^2^=1, Figure S 9). One CRIC participant showed particularly noisy EKG signals where HR confidence was below the 20%-threshold during night hours (1.4±1.1%). Taken together, these data suggest that the BioPatch is well suited to collect cardiovascular data under outpatient conditions.

### Biometric data differentiate day versus night

To assess the internal validity of our dataset, we were interested to see if this biometric sampling approach can differentiate between day (06:00-22:00) and night (22:00-06:00) hours. As expected, day versus night differences in the BioPatch data streams were most pronounced in activity (0.06±0.02 g vs. 0.002±0.01 g; n=47, ANOVA q=1.3×10^−17^), peak acceleration (0.13±0.04 g vs. 0.06±0.01 g; n=47, ANOVA q=2.9×10^−17^), heart rate (68.6±12.7 bpm vs. 55.2±16 bpm; n=47, ANOVA q=6.3×10^−9^), breathing rate (14.4±1.9 bpm vs. 12.8±2.1 bpm; n=47, ANOVA q=0.001), and core temperature (37.02±0.34°C vs. 36.7±0.48°C; n=47, ANOVA q=3.2×10^−5^). To put the activity and acceleration data into context, commercial airline passengers experience an additional 0.1-0.3 force of gravity during takeoff, so that above stated levels may correspond more to the much slower acceleration of a train. The difference in posture (−26.6±22.2° and -36.6±37.1° for day versus night, where values towards zero indicate vertical posture and negative values indicate supine torso positions; n=47) follows expectation (ANOVA p=0.04, q=0.18). The BioPatch-SDNN showed little diurnal difference with a high degree of variability (47.9±23.2 ms vs. 50.4±23.7 ms; n=47, ANOVA p=0.09, q=0.25). These data are shown in Figure 1, Table S 3 and Table S 8.

**Figure 1.**
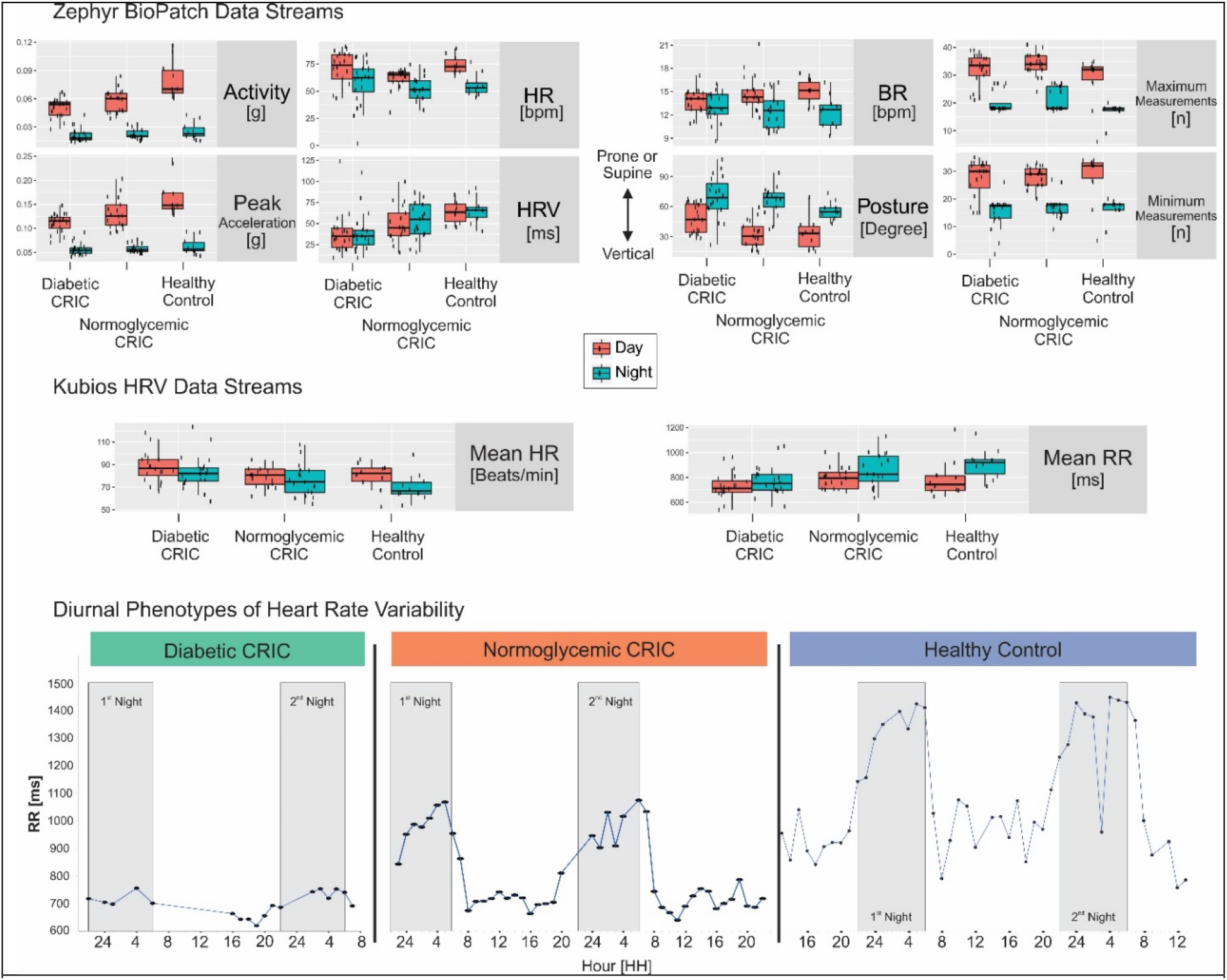
Diurnal Phenotypes on Cohort & Patient Level. **Top:** Boxplots of BioPatch data streams are stratified by cohort diabetic (left) and normoglycemic (center) patient with CKD compared to healthy controls (right) as well as by day (orange) and night (green) for the following readouts: activity (g), peak acceleration (g), heart rate (HR, bpm), SDNN (standard deviation of normal-to-normal RR intervals calculated as rolling heart rate variability value in ms), breathing rate (BR, bpm), and posture (degree where values towards zero indicate vertical posture and negative values indicate prone or supine torso positions). **Center:** Boxplots of EKG waveform data streams analyzed by Kubios. Heart rate (bpm) and interbeat intervals (RR, ms) stratified by cohort as well as day and night. **Bottom:** Time-of-day dependent modulation, or absence thereof, of interbeat intervals (RR, ms) at participant-level for a diabetic (left) and normoglycemic (center) patient with CKD compared to healthy control (right). Grey rectangulars indicate first and second night (22:00-06:00).

We adopted the scientific-grade software Kubios to extend our HRV analyses (readouts detailed in supplemental materials). In the Kubios HRV data, we found that the 250Hz EKG waveform derived heart rate decreased at night (70±23.7 bpm) compared to day (79.3±18.8 bpm; n=47, ANOVA p=0.03, q=0.14). Various HRV summary measures also showed an increase during night hours, as exemplified by mean RR (14.4±1.9 ms vs. 12.8±2.1 ms during active hours; n=47, ANOVA q=0.001) and RMSSD (53.7±32.8 ms vs. 47.8±32.5 ms; n=47, ANOVA p=0.07, q=0.21). The Kubios-SDNN was comparable for day and night, 50.4±24 ms versus 51.4±24.1 ms, n=47, ANOVA q=0.54. Day versus night modulated the normalized spectral heart rate variability for HFnu (36.3±13.1 versus 42±15.4; ANOVA q=0.005) and LFnu (63.6±13.1 versus 58±15.4; ANOVA q=0.005). These shifts indicated an increase in parasympathetic nervous activity throughout the night. The LF/HF ratio was lower at night (1.9±1.6 vs. 2.2±1.9; n=47, ANOVA p=0.04, q=0.18). These metrics display findings in the Fast Fourier Transform (FFT) spectrum, and results for the autoregressive (AR) spectrum are similar. The non-parametric measure of autonomic nervous balance, the SD2/SD1 ratio, decreased from 2.1±0.6 during the day to 1.7±0.6 at night (ANOVA p=0.04, q=0.17). This appears to be the result of an increase in SD2 during the day (62.1±26.2 ms versus 61.36±26.6 ms, ANOVA p=0.007, q=0.05) and an increase in SD1 during the night (33.8±23 ms versus 38±23.2 ms, ANOVA p=0.07, q=0.21), though only the change in SD2 met our cutoff for statistical significance. Table S 5 and Table S 8 contain the complete summary output of the Kubios HRV analysis. Taken together, these data suggest that cardiovascular signals can be reliably collected through wearable technology in the natural settings of the participants’ individual conditions.

### Biometric phenotypes track with health condition

Next, we sought to examine whether the biometric signals can differentiate between CKD and CKD/T2DM (CKD patients comorbid with type II diabetes mellitus) patients, compared to controls, despite the noise introduced by sampling in the wild, confounding co-morbidities, and therapeutic management. Here, we addressed this using a two-way ANOVA (with Benjamini-Hochberg correction) to test for differences in biometric signals across cohort and time of day, followed by post-hoc Tukey tests. Phenotypic differences among cohorts are most pronounced for activity (ANOVA q=9.6×10^−5^) and peak acceleration (ANOVA q=0.0003) and emergent for BioPatch-SDNN (ANOVA p=0.01, q=0.079). Post-hoc tests indicate this significant difference between cohorts for activity is largely driven by divergence between CKD versus control (adj. p=4.7×10^−9^) and CKD/T2DM versus control (adj. p=9.8×10^−13^). Furthermore, by looking at the interactions between cohort and time of day, we find the differences between the controls and CRIC patients are significant during the day (adj. p=3.8×10^−8^ for CKD and adj. p=2×10^−12^ for CKD/T2DM), but not at night (adj. p=0.99 for CKD and adj. p=0.94 for CKD/T2DM). The post-hoc comparisons between CKD and CKD/T2DM did not achieve statistical significance (adj. p=0.43).

A similar pattern appears for peak acceleration with differences driven in the post-hoc test by CKD or CKD/T2DM versus control (adj. p=4.9×10^−8^ and adj. p=3.5×10^−11^, respectively), pronounced during the day (adj. p=2×10^−7^ and adj. p=2.5×10^−11^, respectively) and absent at night. The BioPatch-SDNN trended toward a cohort-level difference (ANOVA q=0.079) with differences between CKD/T2DM and controls (adj. p=0.008) during both day (adj. p=0.024) and night (adj. p=0.021). The remaining BioPatch variables showed no significant differences between cohorts. Table S 6 provides the summary outputs.

The Kubios time-domain results identified several features trending toward significant differences between cohorts: mean heart rate (ANOVA q=0.143), minimum heart rate (ANOVA q=0.142), standard deviation of heart rate (ANOVA q=0.107), and mean RR interval lengths (ANOVA q=0.165) as listed in Table S 7. The numerical values in mean heart rate during the night showed directional differences with 83.5±27.3 bpm for CKD/T2DM patients compared to 77.1±29 bpm for CKD patients and 69.9±23.7 bpm for controls. Similarly, the mean RR during the night was lowest for CKD/T2DM patients (775.4±201.3 ms) compared to CKD patients (852.6±216.6 ms) and controls (915.2±191.9 ms).

In the Kubios frequency domain results, the relative Fast Fourier-transformed very low frequency (% VLF, Table S 7), proposed among other HRV features to predict survival in patients with myocardial infarction ^27^, differed between cohorts (ANOVA q=0.045). This was driven by the divergence between controls and CKD/T2DM patients (adj. p=0.005) for both day (13±4.9 and 18.3±7.4, respectively) and night (13.3±4.6 and 18.9±9.7, respectively). The relative, autoregressive (AR) modelled VLF showed a trend toward significant divergence between cohorts (ANOVA q=0.142), potentially driven by the comparison of controls to CKD/T2DM patients (adj. p=0.037, Table S 7).

The EKG derived respiration (EDR) in the Fast Fourier transformed metrics (Table S 7) was different between cohorts (ANOVA q=5×10^−5^). Here, both sets of CKD patients diverged significantly from controls during day (0.23±0.04 in CKD and 0.22±0.06 in CKD/T2DM versus 0.3±0.05 in controls, adj. p=8.4×10^−7^ and adj. p=3.2×10^−8^, respectively) and night (0.23±0.05 in CKD and 0.22±0.06 in CKD/T2DM versus 0.28±0.06 in controls, adj. p=0.007 and adj. p=0.0002, respectively). The natural logarithmically transformed absolute powers of high and low frequency bands trended toward differences across cohorts (ANOVA q<0.18 and ANOVA q<0.14, respectively, for both parametric autoregressive [AR] modeling and Fast Fourier transformation [FFT]). Post-hoc tests suggest an emerging difference between controls and CKD/T2DM patients (adj. p<0.012 for both transformations, Table S 7).

Among the Kubios nonlinear results, the dimensionless α2 long-term fluctuations in the detrended fluctuation analysis (α2-DFA) differed between cohorts (ANOVA q=0.02). The post-hoc test attributes this to the divergence between CKD and CKD/T2DM (adj. p=0.015) during both the day (0.5±0.13 and 0.58±0.17, respectively, adj. p=0.047) and night (0.45±0.12 and 0.58±0.2, respectively, adj. p=0.004). α2-DFA also showed a significant difference between controls and CKD/T2DM patients (adj. p=0.034) only during the night (0.45±0.11 and 0.58±0.2, respectively, adj. p=0.034). This is visualized in Figure 2. DFA assesses non-stochastic self-similarity by quantifying how current values in time-series data are determined by their past values, a concept coined ‘long-memory’ processes ^28^. Disease conditions, such as diabetes ^29^ and severe obstructive sleep apnea ^30^, have been associated with higher α2-DFA compared to controls. Importantly, increases in α2-DFA were associated with higher all-cause mortality in a Japanese cohort of about 300 septuagenarians and octogenarians ^31^. In the Framingham Heart Study, abnormal cardiac control quantified by a low “DFA index” and other HRV markers was associated with poor survival in a cohort of 69 septuagenarians with chronic congestive heart failure (CHF) ^32^. Of note is that modulation of DFA by age did not emerge in 114 healthy volunteers ^33^.

**Figure 2.**
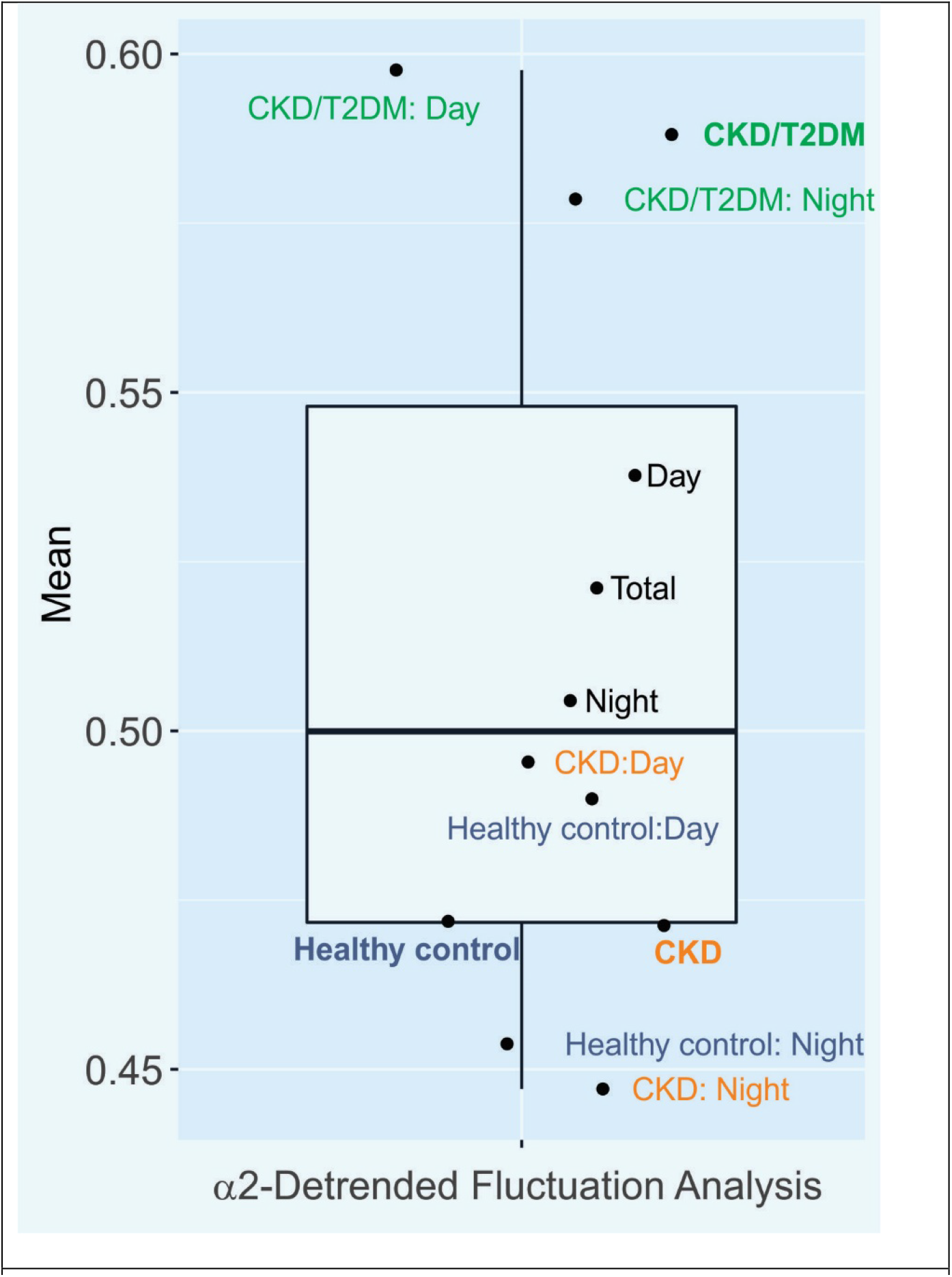
α2-Detrended Fluctuation Analysis (α2-DFA) Boxplot with mean values for the Kubios heart rate variability readout, α2-Detrended Fluctuation Analysis (α2-DFA), are shown for all participants (black dots) as well as for CKD/T2DM patients (green dots), CKD patients (orange dots) and healthy volunteers (blue dots) including day versus night differences.

The correlation dimension D2, a nonlinear HRV feature shown to decrease under acute stress conditions in students ^34^ and lower in patients with dilated cardiomyopathy (DCM) compared to controls ^35^, discriminated well between cohorts (ANOVA q=0.001). Here, controls (2.5±1.6 during day; 2.48±1.54 during night) differ from CKD (adj. p=0.001) and CKD/T2DM (adj. p=1.5×10^−6^), most prominently during the day (1.05±1.27, adj. p=0.004 and 0.58±0.93, 5.5×10^−6^, respectively) and less so during the night (1.33±1.29, adj. p=0.06 and 0.94±1.41, adj. p=0.001, respectively). The CKD and CKD/T2DM patients failed to show a significant difference in correlation dimension D2 (adj. p=0.26).

In summary, we conclude that phenotypic features composed of a subset of BioPatch data streams and Kubios HRV metrics differentiate between patients and controls. Given that these differences were driven largely by comparisons between CKD/T2DM patients and controls, as suggested by the post-hoc tests, but not by differences between CKD patients and controls, it is possible that disease severity affects these relationships in addition to the expected age-related difference between patient and control cohorts. We also note that despite the abundance of parameters, a total of 62 HRV features, and correction for multiple testing, the α2-DFA emerges as a candidate marker to differentiate between CKD and CKD/T2DM cohorts in the present study.

### Loss of rhythmicity in patients

In light of the observation in Figure 1 that diurnal changes in HRV are lost in a CKD/T2DM patient and dampened in a CKD patient compared to a healthy participant, and supported by prior evidence ^11^, we hypothesize that CKD patients have altered rhythm characteristics. We found a significant difference in the MESOR (rhythm-adjusted mean) of activity levels between cohorts (Bonferroni-Holm corrected ANOVA p=1.5×10^−05^, Table S 12). The post-hoc tests suggest that controls are more physically active (0.057±0.012 g) than patients with CKD (0.044±0.026 g, adj. p=5.2×10^−05^) or CKD/T2DM (0.042±0.007 g, adj. p=1.6×10^−06^). Note, that CKD patients did not differ significantly from patients with CKD/T2DM (adj. p=0.5). This difference in activity MESOR is potentially confounded by the age differences between the CRIC patients and the controls, and will require further investigation in future studies., Though the between group differences are substantial for the MESOR of BioPatch-SDNN, lowest in CKD/T2DM patients (38.5±23.6 ms) followed by CKD patients (48±18.9 ms) and controls (62.5±14.1 ms), only the comparison between controls and CKD/T2DM patients attained significance (Bonferroni-Holm corrected ANOVA p=0.01). Differences in MESOR for breathing and heart rate were less pronounced, as shown in Figure 3. The mean amplitude of activity was higher in controls (0.033±0.011 g) compared to both CKD (0.023±0.01, Bonferroni-Holm corrected ANOVA p=0.03) and CKD/T2DM (0.022±0.007, Bonferroni-Holm corrected ANOVA p=0.01) patients, respectively. Statistically, the two patient groups did not differ (Bonferroni-Holm corrected ANOVA p=0.9) from each other. Amplitudes for breath, heart rate, and SDNN failed to show a significant difference between cohorts.

**Figure 3.**
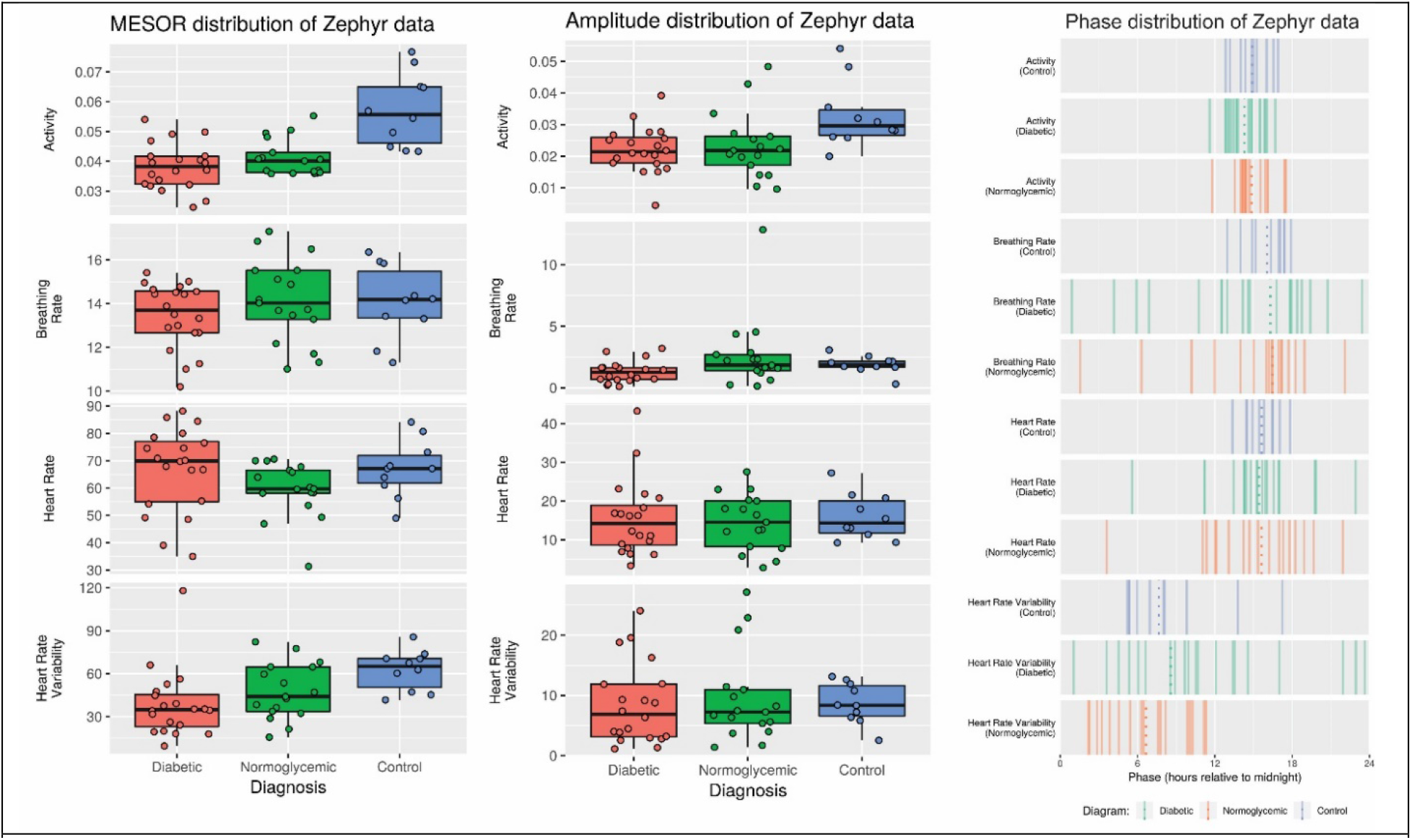
Cosinor metrics of BioPatch data streams. The rhythm-adjusted mean, MESOR, (**Left**) and amplitude (**Center**) for activity (g), breathing (bpm) and heart (bpm) rate and BioPatch SDNN (standard deviation of normal-to-normal RR, ms) as heart rate variability readout are stratified by cohort, i.e. diabetic (left) and normoglycemic (center) patient with CKD compared to healthy controls (right). **Right:** The time-of-day when physiological readouts peak, acrophase, is shown for activity (g), breathing (bpm) and heart (bpm) rate and BioPatch SDNN (ms) for controls (top, blue), diabetic (center, green) and normoglycemic (bottom, orange) patient with CKD.

We observed negligible divergence in the acrophases (peak times) of activity across the three cohorts, with all subjects peaking between 14:30-15:00 in the afternoon, with a consistent error of 1-2 hours in each cohort. This is likely because all the subjects entrained to similar light-dark schedules. Breathing and heart rates show similar acrophases, though the variability in CKD/T2DM and CKD patients is much higher than in controls, which might be reflective of the loss of rhythmicity in these patients. In contrast, the mean acrophase of SDNN occurs much earlier in the day, at around 08:30 for CKD/T2DM patients compared to 06:20 for CKD patients and 08:00 for controls. Again, we observe increased variability in SDNN acrophases among CKD patients, as shown in Figure 3. Supporting data is shown in Table S 11.

Overall, these data suggest that the diurnal rhythmicity of CKD patients is dampened relative to controls.

### Phase relationships deconsolidate in patients

Next, by comparing the acrophases across different physiological data streams (like activity and SDNN) within each subject, we sought to assess whether disease states can alter or disrupt an individual’s circadian organization. This is similar to a previous study which found shift work decreased the acrophase angle between HRV readouts and physical activity ^36^. Using the acrophase of physical activity as a reference in the present study (Figure S 8), we calculated the within-subject differences between the acrophase of activity, and the acrophases of breathing rate, heart rate, and BioPatch-SDNN. The healthy controls showed minimal differences for both breathing rate (1h, i.e. Cosinor PhaseActivity of 14.9h versus Cosinor PhaseBR of 15.9h in Table S 10) and heart rate (0.6h), but a substantial difference for the BioPatch-SDNN (mean phase difference of 6.8h). CKD and CKD/T2DM patients showed similar acrophase differences between activity and breathing or heart rate. Notably, the difference in acrophase between activity and BioPatch-SDNN showed a substantial amount of variability in CKD (mean phase difference of 8.3h) and CKD/T2DM patients (mean phase difference of 4h) compared to healthy controls, as shown in Figure S 8, left. To address the concern that loss of rhythmicity may confound the cosinor determination of acrophases, we applied a more stringent weighted smoothing method, LOESS, to confirm the results as shown in Figure S 8, right and Table S 10. Strikingly, the dispersion of acrophases of BioPatch-SDNN spans the entire 24-hour range displayed in Figure S 8, suggesting deconsolidation of acrophase differences among the CKD/T2DM patients, in particular.

### Altered circadian organization of biometric variables in CKD patients

In an effort to define ways to condense the data streams from the BioPatch and the Kubios HRV analysis, we applied the following strategy. First, we constructed a variance correlation matrix to capture how the observed variability is shared among pairs of biometric variables. This measures the proportion of variance observed in one variable that can be explained by the variance observed in another variable. This, in a way, examines circadian organization among biometric variables in that two variables with similar temporal patterns should be highly correlated over time. Thus, we used this metric to confirm expectations within domains, such as the high correlations between peak acceleration and activity (R^2^=0.95, Bonferroni corrected p<0.001, observed in all cohorts). While this increases our confidence in the validity of these biometric assessments, it also allows us to explore how the variance explained shifts between participants and cohorts. This is shown, for example, in Figure S 4 for the BioPatch-derived data streams on the individual and cohort levels. Across domain, we observed the expected correlations between activity and heart rate (R^2^=0.22, Bonferroni corrected p<0.001), as well as activity and breathing rate (R^2^=0.15, Bonferroni corrected p<0.001) in controls. These correlations were lower in both CRIC patient cohorts for activity and heart rate (R^2^=0.10, Bonferroni corrected p<0.001), and for activity and breathing rate (R^2^=0.03, Bonferroni corrected p<0.001). Heart rate and body core temperature correlated well in patients (R^2^=0.77, Bonferroni corrected p<0.001 in CKD/T2DM and R^2^=0.78, Bonferroni corrected p<0.001) compared to controls (R^2^=0.51, Bonferroni corrected p<0.001), though the computational derivation of body core temperature from the heart rate signal may confound this. The high correlation between posture and sagittal acceleration likely reflects body movements in the sagittal plane from sitting down (R^2^=0.71, Bonferroni corrected p<0.001 in CKD/T2DM and R^2^=0.79, Bonferroni corrected p<0.001) compared to controls (R^2^=0.54, Bonferroni corrected p<0.001). The only correlation emerging for the BioPatch-SDNN is with heart rate (R^2^=0.14, Bonferroni corrected p<0.001 in CKD/T2DM and R^2^=0.12, Bonferroni corrected p<0.001 in CKD, compared to controls R^2^=0.11, Bonferroni corrected p<0.001).

Above, we calculated variance correlations from data we aggregated by cohort. We observe more drastic differences in these correlation patterns when we examine data from individual subjects. Comparing the two patients displayed in Figure 1, the CKD patient’s SDNN and heart rate correlate at R^2^=0.12 (Bonferroni corrected p<0.001) while this is lost in the CKD/T2DM patient (R^2^=0.01, Bonferroni corrected p<0.001). Similarly, correlations are weaker in the CKD/T2DM patient for heart rate and activity (R^2^=0.11, Bonferroni corrected p<0.001), and for breathing rate and activity (R^2^=0.09, Bonferroni corrected p<0.001) compared to the CDK patient (R^2^=0.25, Bonferroni corrected p<0.001 and R^2^=0.30, Bonferroni corrected p<0.001, respectively). These findings suggest that this metric may be a feasible representation of biometric phenotypes.

Next, we used these R^2^ values derived from the variance explained metric to generate a hierarchical cluster analysis of all subjects, exploring similarities between study participants in their biometric variables. While these clusters did not provide complete separation between the cohorts, we did observe clusters consisting predominantly of controls or a mix of CKD/T2DM and CKD patients (Figure S 5, top). These patterns naturally suggest a hypothesis for future studies to test: do the features captured by these clusters associate with disease risk or trajectory. For example, do the similarities shared by the single healthy volunteer (highlighted by the red arrow in Figure S 5, bottom) with the seven CKD/T2DM (green) and five CKD (orange) patients suggest a comparable risk profile? Or inversely, does the similarity between the single CKD (highlighted by the blue arrow in Figure S 5, bottom) patient and the five controls (blue) point towards a low risk for disease progression?

Many sources contribute to the variability observed in a data set, particularly for data gathered outside of a controlled clinic. As a result, we sought to quantify how much of the overall variability in our data is contributed by temporal differences in biometric measurement versus inter-subject differences. Consequently, a biometric variable with a strong temporal pattern in a homogeneous cohort will have a much higher contribution of time to variance than inter-subject differences. This approach offers an opportunity to gauge, between cohorts, the disconnect of temporal relationships between biometric variables. This is visualized in Figure S 6 where the contribution of time to the observed variability in activity is consistently larger across all cohorts than the contribution to variance by inter-subject differences (green cluster below the diagonal line of identity in Figure S 6). This underscores the strong temporal signal in activity. This is different for heart rate where only the controls show this pattern (blue data point below line of identity in Figure S 6). For breathing rate and BioPatch-SDNN, the contribution to variability is mostly driven by between-patient differences. Notably, the largest separation between cohorts occurred for SDNN where 49.5%, 36.1%, and 19.2% of the variances were contributed by inter-subject differences among CKD/T2DM patients, CKD patients, and controls, respectively. Complete time-versus-subject contribution to variance outputs are provided in Table S 9. Overall, this suggests that the behavioral (activity) phenotype, entrained by the light-dark cycle, is intact among all cohorts. However, rhythms in the cardiovascular (heart rate) phenotype appear to dampen in CKD/T2DM and CKD patients compared to controls.

Taken together, these data reduction strategies suggest the hypothesis that the circadian organization of biometric variables is altered in CKD patients and that this disruption is in part associated with disease state in CKD/T2DM versus CKD patients. Furthermore, these data demonstrate our ability to use wearable devices outside the clinical setting to detect significant alterations in physiological patterns.

## Discussion

We undertook this study to test the feasibility of deriving actionable insight into HRV measures in patients with CKD under outpatient conditions. The range of cardiovascular responsiveness to the challenges of daily life is larger than in the clinic, where conventional EKG and HRV assessments are made. Clearly, the challenge is to discern signal from noise in the natural setting. This approach enables more robust inferences than if there were only a small number of observations per patient. In this pilot study, we observed that i) the wearable device collects interpretable data in freely moving participants, and that ii) behavioral and cardiovascular parameters differed between day and night hours according to expectations. Among the BioPatch data streams and Kubios HRV metrics, only a subset of features passed the correction for multiple testing, highlighting the challenge that the number of parameters exceeded the sample size of participants in this study. The post-hoc tests indicated that phenotypic divergence may be largest between CKD/T2DM patients compared to controls and much less between CKD patients compared to controls. While this may represent the additive burden of risk-increasing co-morbidities, this may also be driven by the age difference between cases and controls in the present study. Notably in this context, a nonlinear parameter, i.e. α2-DFA, is significantly different between CKD and CKD/T2DM patients, suggesting this as a potential biomarker. Focusing on the diurnal patterns in the behavioral and cardiovascular phenotypes, our findings suggest that biorhythms are less robust in CRIC participants compared to controls and that phase relationships between HRV, heart/breathing rate, and physical activity may be deconsolidated among the CKD/T2DM patients, in particular. This latter finding is reminiscent of the abnormal acrophase differences between physical activity and the cardiac autonomous nervous system observed in rotating shift workers ^36^. This may translate into an increased risk of mortality. The MrOS Sleep Study ^37^ found a u-shaped association of mortality with rest-activity rhythms among older men with cardiovascular disease. Patients with the most pronounced phase advance in peak activity (lowest quintile, 08:44-13:22 HH:mm) showed a substantially elevated mortality (a hazard ratio of 2.84 with a 95% confidence interval of 1.29-6.24) compared to the reference patients (middle quintile, 13:59-14:32 HH:mm) ^37^. In contrast, patients with a phase delay (highest quintile, 15:09-23:30 HH:mm) seem to have a similar risk of mortality (a hazard ratio of 1.55 with a 95% confidence interval of 0.67-3.6) compared to the reference patients of the middle quintile ^37^.

We examined variability and temporal relationships by applying variance correlation, time-versus-subject-contribution-to-variance, and hierarchical clustering to transform the data from the present study guided by prior experience ^38^. The value of these preliminary data is to gain confidence in parsing mechanisms associated with disease expression in cohorts of patients and age- and sex-matched controls. Here, using the variances of the time-domain, (BioPatch-SDNN) as a marker, we estimate that sample sizes of n=5 per cohort are required to detect a difference of 50 ms with 90% power at a significance threshold of 0.05. We confirmed these low sample sizes by using the Kubios-SDNN, which resulted in n=8 required per cohort to detect the same difference. This is inspired by the proposed thresholds associated with healthy conditions (SDNN>100ms), compromised health (SDNN=50-100ms), unhealthy (SDNN<50ms), and cumulative survival after myocardial infarction ^39,40^. To account for differences in patients and technology, we recommend a more stringent power calculation. For instance, a sample size of n∼44 subjects per cohort would be required to detect a difference of 0.5 standard deviations in SDNN with 90% power and a significance threshold of 0.05. Turning to α2 DFA, a sample size of n=12 per cohort would detect a difference of 0.14 in α2 DFA with 90% power at a significance threshold of 0.05. This difference of 0.14 is reflective of the decreased parasympathetic modulation noted in diabetic patients compared to healthy controls ^29^. Based on the variance in our study, 44 subjects per cohort would be necessary to detect a difference of 0.5 standard deviations in DFA α2. Notably, the range of α2 DFA values in our data (0.45-0.6) is much lower than the range listed in Roy & Ghatak ^29^ (0.88-1.02), possibly owing to the different collection periods, 10 min in their study ^29^ versus 2 days in the present study.

Biometric data provide insights into dynamic changes in vital signs, activity, and many other aspects of health. Recent advances in technology for remote capture of biometric data offer the opportunity to understand the effects that co-morbidities like CKD have on lifestyle in the places where patients live and move. The growing capacity to incorporate continuous electrocardiographic data, for example, can offer information on the prevalence of cardiovascular risk in this population that is not recorded in the confines of a brief office or research study visit. Since CKD is plagued by a higher age-related death rate, clear impairment of quality of life, and physical frailty, remotely captured biometric data will be useful to aid our understanding of how things like physical activity, short sleep duration, and vagaries in the circadian rhythm affect outcomes like death and further disability in the CKD population. Given the rise of cardiovascular complications and kidney injury among patients with COVID-19, telemedicine efforts should be paired with wearable biometric approaches to improve the clinical care space.

## Methods

### Study population

This two-center study enrolled a total of 39 patients without a diagnosis of atrial fibrillation, n=20 at University Hospitals of Cleveland Medical Center and n=19 at the Hospital of the University of Pennsylvania, HUP. Ethics approval for the addendum to the CRIC clinical study protocol (Penn IRB#707819) was granted by the Institutional Review Board of the University of Pennsylvania (Federalwide Assurance FWA00004028; IRB Registration: IORG0000029) and by the UH Cleveland Medical Center (Federalwide Assurance 00003937; IRB Registration: 02-03-04) in compliance with the guidance issued by the International Conference on Harmonization (ICH) harmonized tripartite guidelines: E6 Guideline for Good Clinical Practices. For the control group, n=10 healthy volunteers were recruited under Penn IRB#828728 at the Institute for Translational Medicine and Therapeutics, University of Pennsylvania. Informed consent was obtained from all participating patients prior to study activities. Study-related time commitment was compensated with a modest stipend. This cohort represents a sample of “convenience” without pre-defined selection bias by study personnel or investigators (CONSORT statement in Figure S 1 and supplemental methods).

### Wearable Device

This study used the Zephyr BioPatch (Zephyr Technology, Annapolis, MD), distributed by Medtronic Corporation (Minneapolis, MN). This research-grade wearable device records cardiovascular, respiratory, and behavioral data consistent with its FDA Class II clearance as a “physiological monitoring telemetry device intended for monitoring adults in the home, workplace and alternate care settings” (510(k) # K113045). This device has been deployed in the field, for example, to collect physiological monitoring of Chilean miners during the San Jose Mine rescue operation ^41^. In the present study, the target observational time for each patient was set to 48 hours to cover two consecutive 24h circadian cycles in order to enhance detection of biological rhythms ^42^. Patients received a plug-in charging cradle and were instructed to remove the BioPatch device (white “puck”) from its chest-mounted holder after approximately 24 hours of use to recharge the device for approximately two hours followed by inserting it back into the BioPatch holder on the chest for continued recordings. Positioning of the two EKG snap electrodes to hold the BioPatch holder in place followed standard EKG guidelines for V1 and V2, i.e. V1 corresponds to the right 4^th^ intercostal space; and V2 corresponds to the left 4^th^ intercostal space. Prior to application of the snap electrodes skin was cleaned with rubbing alcohol and shaved if necessary. Healthy controls received two BioPatch devices and were trained and instructed to exchange the first by the second device after 24 hours.

### Biomedical Informatics

EKG recordings were analyzed in 1-hour increments in Kubios HRV Premium (ver. 3.0, Kubios Team, Kuopio, Finland) to obtain time-of-day-dependent measures of heart rate variability. Custom perl and R code (GitHub, “WearablePhenotypingCRIC”) formatted these data and integrated with BioPatch data streams for heart rate, breathing rate, breathing waveform, posture, accelerometry, and peak/minimum acceleration. In addition to summary statistics for cohort and time-of-day (wake versus sleep hours), data were parsed i) by cosinor analysis to obtain the rhythmic parameters amplitude and phase, ii) by two-way ANOVA analysis and post-hoc Tukey test corrected for multiple testing with the Benjamini-Hochberg method to discover significant associations, iii) by methods reducing data dimensionality to uncover meaningful relationships. Detailed descriptions are provided in Supplemental Methods.

## Data Availability

All data produced in the present work are contained in the manuscript and its supplemental materials

## CRIC Consortium Members

Lawrence J. Appel, MD, MPH^9^, Alan S. Go, MD^10^, Jiang He, MD, PhD^11^, James P. Lash, MD^12^, Robert G. Nelson, MD, PhD, MS^13^, Panduranga S. Rao, MD^14^, Vallabh O Shah, PhD, MS^15^, Mark L. Unruh, MD, MS^16^

^9^ Department of Medicine, Johns Hopkins Bloomberg School of Public Health, Baltimore, MD;

^10^ Kaiser Permanente Division of Research, Oakland, CA

^11^ Department of Epidemiology, Tulane University School of Medicine, New Orleans, LA;

^12^ Department of Medicine, University of Illinois Chicago, Chicago, IL;

^13^ National Institute of Diabetes and Digestive and Kidney Diseases, Chronic Kidney Disease Section, Phoenix Epidemiology and Clinical Research Branch, National Institute of Health, Bethesda, MD;

^14^ Department of Medicine & Nephrology, University of Michigan, Ann Arbor, MI;

^15^ Department of Biochemistry and Molecular Biology, Division of Nephrology, University of New Mexico, Albuquerque, NM;

^16^ Department of Internal Medicine, University of New Mexico, Albuquerque, NM.

## Acknowledgements

We are indebted to our volunteers who consented to participate in this pilot study as well as to Angela Sheridan. Parts of this study were presented by C.S. at the Gordon Research Conference on Chronobiology, June 23-28, 2020, Spain.

## Authors’ Contributions

N.F.L. created visualizations and worked on statistical analysis, biomedical informatics, data interpretation, writing, manuscript revision, visualizations, figures & tables. M.R. executed literature search, study design, regulatory science, data collection, and manuscript revision. J.B.C., D.L.C., J.C., M.R.W. and G.R.G. worked on data analysis, data interpretation, and manuscript revisions. R.R.T. conducted literature search and worked on study design, regulatory science, data collection, data interpretation, as well as manuscript revisions. C.S. conducted literature search and worked on study design, regulatory science, study conduct, data collection, data analysis, data integration, data interpretation, writing, visualizations as well as figures & tables.

