## Supplements for "Time-Specific Associations of Wearable, Sensor-Based Cardiovascular and Behavioral Readouts with Disease Phenotypes in the Outpatient Setting of the Chronic Renal Insufficiency Cohort (CRIC)"

**and the CRIC Study Investigators***

[1] Institute for Translational Medicine and Therapeutics (ITMAT), University of Pennsylvania Perelman School of Medicine, Philadelphia, PA;

[2] Department of Systems Pharmacology and Translational Therapeutics, University of Pennsylvania Perelman School of Medicine, Philadelphia, PA;

[3] University Hospitals Cleveland Medical Center, Case Western Reserve University, Cleveland, OH;

[4] Department of Medicine, University of Pennsylvania Perelman School of Medicine, Philadelphia, PA;

[5] Department of Medicine, Tulane University School of Medicine, New Orleans, LA;

[6] Division of Nephrology, Department of Medicine, University of Maryland School of Medicine, Baltimore, MD;

[7] Department of Biostatistics and Epidemiology, University of Pennsylvania Perelman School of Medicine, Philadelphia, PA;

[8] Department of Genetics, University of Pennsylvania Perelman School of Medicine, Philadelphia, PA.

* Lawrence J. Appel, MD, MPH, Alan S. Go, MD, Jiang He, MD, PhD, James P. Lash, MD, Robert G. Nelson, MD, PhD, MS, Panduranga S. Rao, MD, Vallabh O Shah, PhD, MS, Mark L. Unruh, MD, MS

**Authors for correspondence:**

Nicholas F. Lahens, Ph.D. and Carsten Skarke, M.D.

**Supplemental Methods**

#### In-/Exclusion criteria & recruitment

This clinical research study was carried out in accordance with relevant guidelines and regulations. Inclusion criteria were defined as the person being a current CRIC participant at either of the two study sites and able to give informed consent. Exclusion criteria were defined as i) resident in a nursing home; ii) lost to follow up; iii) non-ambulatory; iv) presence of a heart pacemaker or automatic defibrillator; v) presence of skin rash or irritation at the thoracic site where the monitor patches would be placed; vi) most recent score on Modified Mini-Mental State Examination lower than 80; vii) ESRD; and viii) participants judged by study personnel to be incapable of completing the procedures. The sampling strategy at each center was set to enroll consecutive participants who were scheduled for regular, annual in-clinic CRIC visits between late January and August 31, 2017 (HUP, n=194) and between April 25 and September 22, 2017 (Cleveland, n=56). 29 CRIC participants were approached at Penn of whom 19 consented for study participation, 10 declined for various reasons. At Cleveland, 23 CRIC participants were approached, 20 consented, 3 declined due to scheduling issues. None of the consented participants dropped out. Manual chart review confirmed the absence of a diagnosis of atrial fibrillation in patients from the Penn cohort; however, it was noted that in one case the electronic health record did not hold much information while in another case no diagnostic code was listed, suggesting that the primary care for these two patients occurred mostly outside of the Penn health care system. The data-driven assessment of the outlierness for BioPatch-SDNN resulted in 122±15.1 (Subject Of Interest SOI#1) and 64.7±20.1 (SOI#2) which compares to 47.3 (Median 40.3; Q1 24.5; Q3 63.1), and for Kubios-SDNN in 178±19.9 (SOI#1) and 90.5±25.7 (SOI#2) which compares to 46.2 (Median 31.5; Q1 18.4; Q3 58.2), as visualized in Figure S 11. This suggests that SOI#2 is closer to the main distribution, while SOI#1 seems to be more of an outlier; however, none of these two patients was excluded from analysis. This strategy underscores the sample collection under real-world conditions. Also, Figure S 11 illuminates that the two patients' central tendency in SDNN is higher than the CKD and CKD/T2DM cohorts, thus biasing against detecting a difference between patients and controls.

For the Cleveland cohort, manual chart review produced one patient with a history of atrial fibrillations. Unrelated to this finding, this patient was excluded from analysis due to unreliable heart rate readings with HR confidence <20% (Figure S 1).

Visual inspection for episodes of atrial fibrillation is not feasible considering that the EKG print out at 25 mm/sec for the present study cohort would amount to ≈113,000 pages. Machine learning approaches to identify episodes of atrial fibrillation are in development for standardized conditions, for example, 10-second, 12-lead EKG reads in supine position [1]; however, reliable artificial intelligence (AI)-driven solutions to interpret long-term wearable device EKG data are expected to require a higher level of sophistication.

#### Wearable Device Outputs

Using the proprietary software, OmniSense (Zephyr Technology, Annapolis, MD), BioPatch devices were initialized (Zephyr Config Tool, Firmware Version v1.0.24.0; BioPatch Boot Software Version v1.3.1.0) using the log format "Summary and Waveform" to record timestamped data on a single-lead EKG waveform [bits; 2048 bits = 0 mV; 1 bit = 0.00625mV] sampled at 250Hz, 3-axis accelerometer [bits; 1 g = 83 bits] sampled at 100Hz, breathing waveform [bits] at 25Hz, and heart rate [bpm], breathing rate [Breaths per minute], posture [degrees from vertical, where 0° denotes that the person is vertical, 90° that the subject is prone (face down), and -90° that the subject is supine (face up)], activity [vector magnitude units, VMU, measured in g], peak acceleration [g], EKG amplitude [Volts], EKG noise [Volts], HR confidence [%] (derived from the signal-to-noise ratio of the EKG signal values >20% indicate reliable heart rate readings, 0% indicates not worn or extremely noisy EKG signal), SDNN HRV [standard deviation of normal-to-normal RR intervals, ms] as a rolling 300 heartbeat HRV value updated once per second, vertical minimum acceleration [g], vertical peak acceleration [g], lateral minimum acceleration [g], lateral peak acceleration [g], sagittal minimum acceleration [g], sagittal peak acceleration [g], and body core temperature [degree C] (estimated from heart rate data) as physiological metrics at a sample rate of 1Hz. Zephyr device related recordings at 1Hz included battery voltage [Volts], battery level [% charge] (to monitor battery health), ROG (device status indication; 0=Invalid ROG, 1=Green, 2=Orange, 3=Red), ROG Time [sec] (time duration in unchanged status), device temperature [degree C], Aux ADC1/2/3 [bits] (three channels to measure hardware circuit noise). Metrics used for Zephyr internal development (BR Amplitude [bits]) or related to the deactivated Bluetooth function (status info, link quality, RSSI [dB], Tx power [dBm]) were not submitted for further analysis. Additional device settings included i) disabled the “discoverable Bluetooth” option to save battery life of the BioPatch device; ii) enabled "Log Enable", "Visual Feedback Enable", and "Event Mode Enable"; and iii) disabled "EKG Polarity Invert", Bluetooth Enable", and "ECHO Enable". Data recordings were started by pressing the button at the center of the BioPatch; data logging was visually confirmed on the device by a blue light flashing at regular intervals. Upon return of the BioPatch after approximately 48 hours, data were downloaded using the OmniSense software, Zephyr Downloader module, producing a suite of .csv files (named “Accel”, “BB”, “Breathing”, “EKG”, “Event_Data”, “RR”, “SessionInfo”, and “Summary”) suitable for data analysis.

#### Heart Rate Variability (HRV) Analyses with Kubios

The 250Hz EKG waveform data contained in the “ECG.csv” file was submitted to Kubios HRV Premium (ver. 3.0, Kubios Team, Kuopio, Finland). Given the size of these files, up to 1GB for a 24-hour recording, only naïve files were submitted to Kubios, meaning that these files were not opened by Microsoft Excel 2010, for example, to quality check data, as the number of data rows exceeds the capability of Microsoft Excel 2010, thus corrupting the “ECG.csv” and making it irreparable. HRV analysis used the following settings: i) automatic artifact correction, ii) Smoothn priors, iii) Lambda=500, and iv) single sample analysis type. To allow analysis of diurnal variability, data analysis was conducted in 1-hour intervals which were defined to start on the full hour, e.g. 14:00 + 00:59:59. Data bins smaller than 00:59:59, at the beginning and end of each continuous recording, were discarded.

These data were curated per “Artifacts Corrected %” filter where 1-hr bins with ≥5% were excluded from further analysis in order to prevent a distortion of the signal. Raw Kubios results for each patient were formatted for use in downstream analysis scripts and software using a custom perl script (gathers all metrics from a Kubios result file into tabular format saves them to a csv file). These csv files were read into R for all remaining analyses and visualizations (described below). Kubios data points with an "Artifacts Corrected %" greater than 5 were excluded from all downstream analyses. The perl and R code used for all analyses will be made available on GitHub (WearablePhenotypingCRIC).

##### Processing Zephyr BioPatch Data

The Zephyr BioPatch includes additional data streams that are not processed or included in the Kubios results. These data include heart rate, breathing rate, breathing waveform, posture, accelerometry, and peak/minimum acceleration. Again, these data were formatted, merged, and analyzed using custom perl and R code available on GitHub. Note, data from the healthy volunteers required an additional merge step to combine data collected by two different BioPatch devices. Five of the healthy volunteers had overlapping data from the two BioPatch devices (i.e. both devices had data entries for the same timepoints). This conflict was resolved through manual inspection of the overlapping data from each device. In all cases one device clearly had invalid data values for the overlapping time spans.

##### Accelerometry

Accelerometer data sampled at 25Hz were sourced from the BioPatch file output named “Accel”. The raw data output is count for each accelerometer. This is transformed into vector magnitude to generate a composite measure following $\sqrt{V^2 + L^2 + S^2}$ where $V$ is the vertical axis count, $L$ is the lateral axis count, and $S$ is the perpendicular axis count. The derived vector magnitudes were then binned into 1-minute intervals, 60 second epochs, carrying [g • $min^{-1}$] as unit.

#### Cosinor Analysis

The cosinor method (reviewed in [2,3]) was used to fit cosine curves to HRV and accelerometer data for each patient. These fits yielded MESOR, amplitude, and phase values for each combination of subject and data type.

#### Two-Way ANOVA analysis

For each patient, we calculated the mean measurement during the day (07:00-21:59) and the night (22:00 - 06:59) for each Zephyr Biopatch variable. We used R's aov function to calculate the two-way ANOVA test for each Zephyr Biopatch variable between patient cohort (Healthy control, CKD, CKD/T2DM) and time of day (day/night), including an interaction term. The aov function generated three p-values: one for each factor, and one for the interaction term. Next, we corrected the entire list of p-values from all Zephyr Biopatch measurements for multiple testing with the Benjamini-Hochberg method, implemented in R's p.adjust function. Lastly, we identified pairs of significant factors within the Zephyr measurements with a post-hoc Tukey test, implemented in R's TukeyHSD function.

#### Variance Correlation Analysis

As piloted in [4], we used this metric as an integrative approach to explore the relationships between the various datasets we collected. Similar to the analysis of HRV in 1-hour bins, all data were aggregated at a one-hour resolution. This was achieved by calculating the mean of each measure for a given patient for each one-hour interval. This produced an hourly summary which was then submitted for further analysis. Per Zephyr data descriptors, invalid values, “4095” for example for the accelerometer raw data outputs or “0” and “16777216” for the breathing waveform raw data stream, were removed prior to data handling. After such data processing, each row in the final integrated dataset included the hour-aggregated measures of all variables for a given patient at a given hourly time index, as defined by the hours after the first epoch “00:00” of commencing data collection. This dataset included every hour/patient combination present in the biosensor and Kubios HRV datasets. As we intended to perform linear regression, we plotted the distribution of each variable. To improve normality as a condition of linear regression, we transformed the biosensor and Kubios HRV datasets including the derived circadian signal variables using a log(x+1) transformation.

To evaluate the linear relationships between every pairwise combination of variables in the integrated dataset prepared above, we calculated the linear regression coefficient of determination ($R^2$), for each pair of variables, using the *lm* function in the R statistical language. We then constructed a heat map of the proportion of variance in each variable (e.g. HRV SDNN, accelerometer vector magnitude, breathing waveform) explained by every other variable. In addition to the $R^2$ values, we also examined the p-values produced by the linear regression analysis. We produced heatmaps for the p-values as well, but only displayed data for those p-values that remained significant following a Bonferroni correction.

#### Hierarchical Cluster Analysis

For each subject, we generated lists of $R^2$ values between every combination of variables, as described in the the Variance Correlation Analysis section above. Next, using these lists of $R^2$ values we calculated the pairwise Euclidean distances between each subject and performed hierarchical clustering with the *dist*() and *hclust*() functions in R, respectively. Note, Euclidean distance is a valid distance metric since the $R^2$ values for all Zephyr and Kubios metrics are mapped to the same range [0,1]. Also we excluded all MSE variables since they have near-perfect correlation with each other and almost no correlation with any other variables. When included, the large number of MSE variables drove clustering and overshadowed the effects of all other variables. Lastly, following clustering we identified five subjects (three diabetic and two normoglycemic) that were substantially separated from all others. Closer inspection revealed several of these subjects had Kubios data with a high number of artifacts, so we exclude these subjects from the clustering analysis.

#### Time-versus-Subject Contribution to Variance

For the Zephyr-derived metrics activity (vector magnitude), breathing rate, heart rate, and heart rate variability, we used the method described in Skarke et al. [4] to calculate the contributions of time and subject to the total variability within each metric. Briefly, these calculations are analogous performing an $R^2$ calculation in linear modeling. First, we calculate the sum of the squared differences between each data point and the mean of all data ($SS_{total}$). Next, we group the data by either hour or subject, and calculate the sum of the squared differences between the means of each group-level and the mean of all data ($SS_{group}$). The contribution of time/subject to total variability is the ratio of $SS_{group}$ and $SS_{total}$. We performed these calculations separately for the normoglycemic and diabetic CKD patients, and health control subjects.

**Figure S 1. CONSORT Statement**

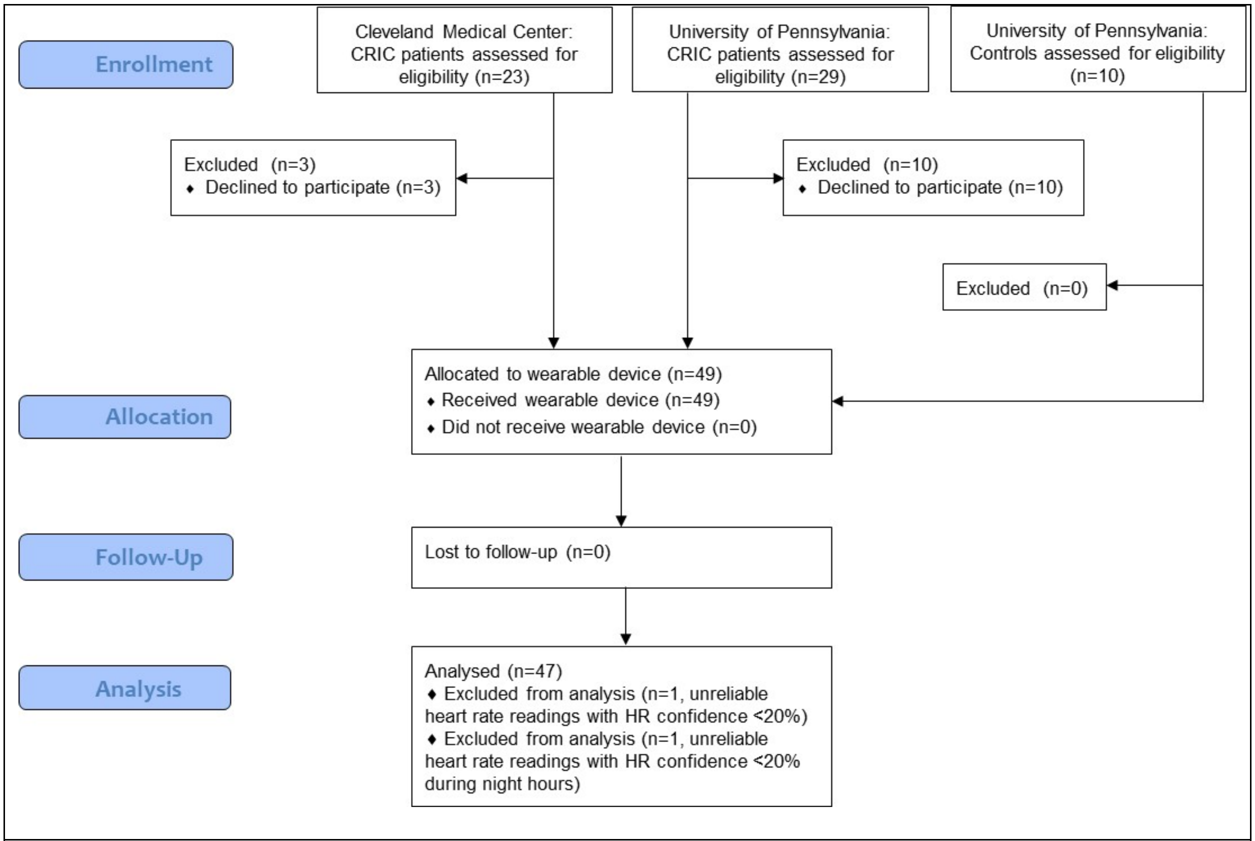


**Figure S 1.** The CONSORT Statement provides an overview of subject enrollment and retention for this observational study of a convenience sample of patients and healthy volunteers.

**Figure S 2. BioPatch Wear Times**

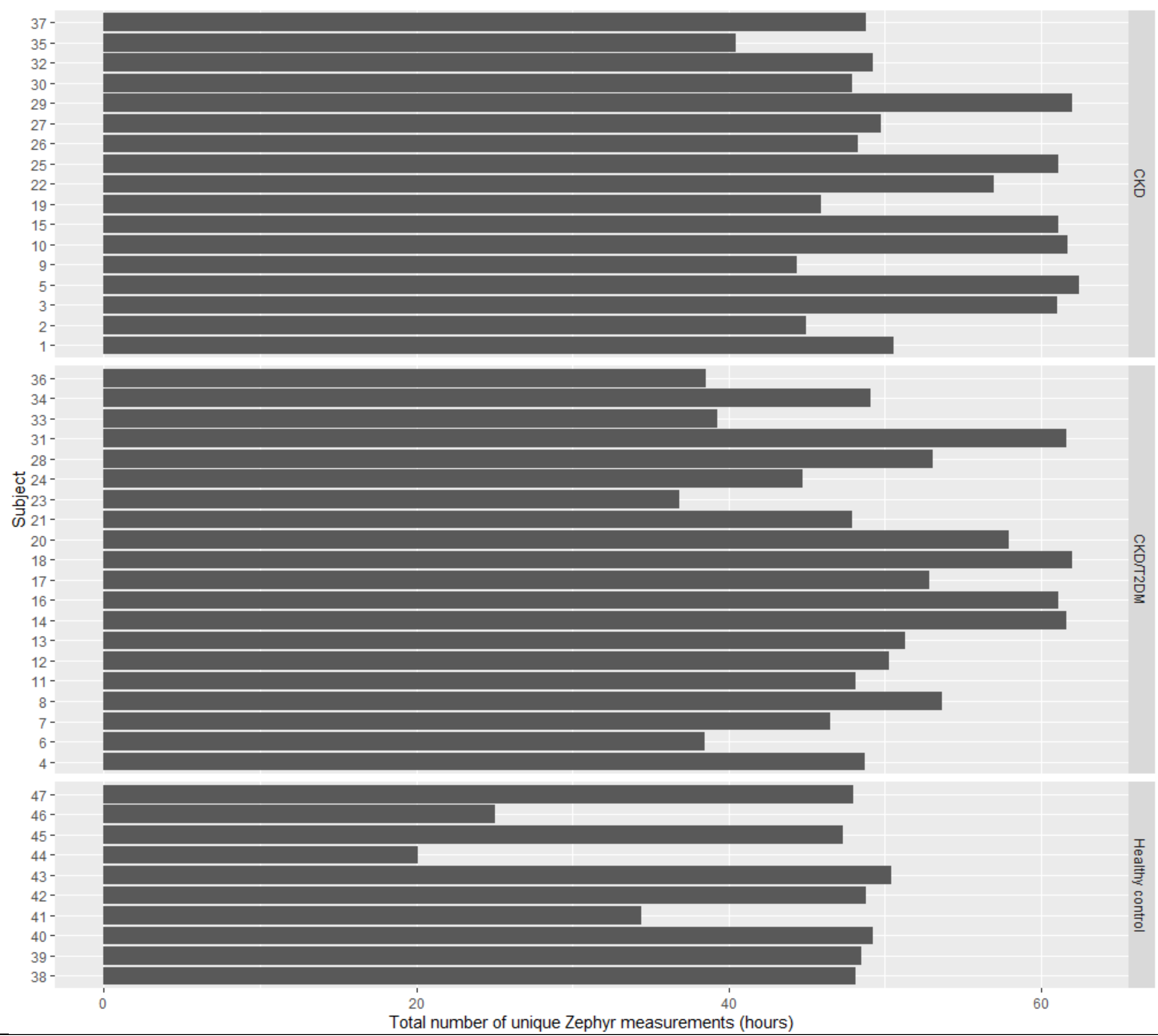


Figure S 2a. Individual BioPatch wear times generated by counting the total number of Zephyr BioPatch measurements. Participant IDs are replaced with arbitrary integers.

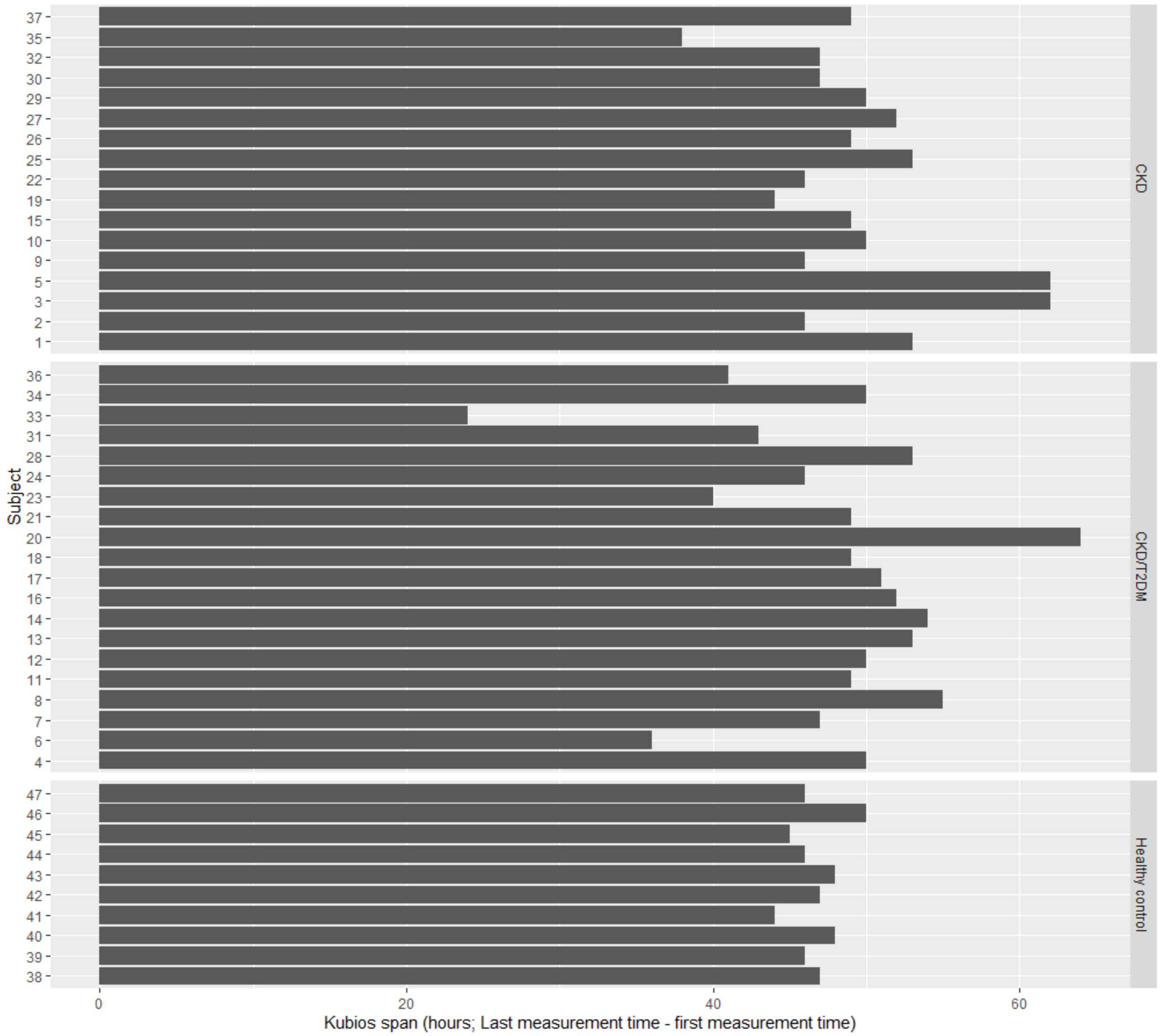


Figure S 2b. Individual BioPatch wear times generated from the Kubios HRV data file by subtracting the timestamps from the last and first measurement periods. Participant IDs are replaced with arbitrary integers.

**Figure S 3. Clinical & Laboratory Parameters**

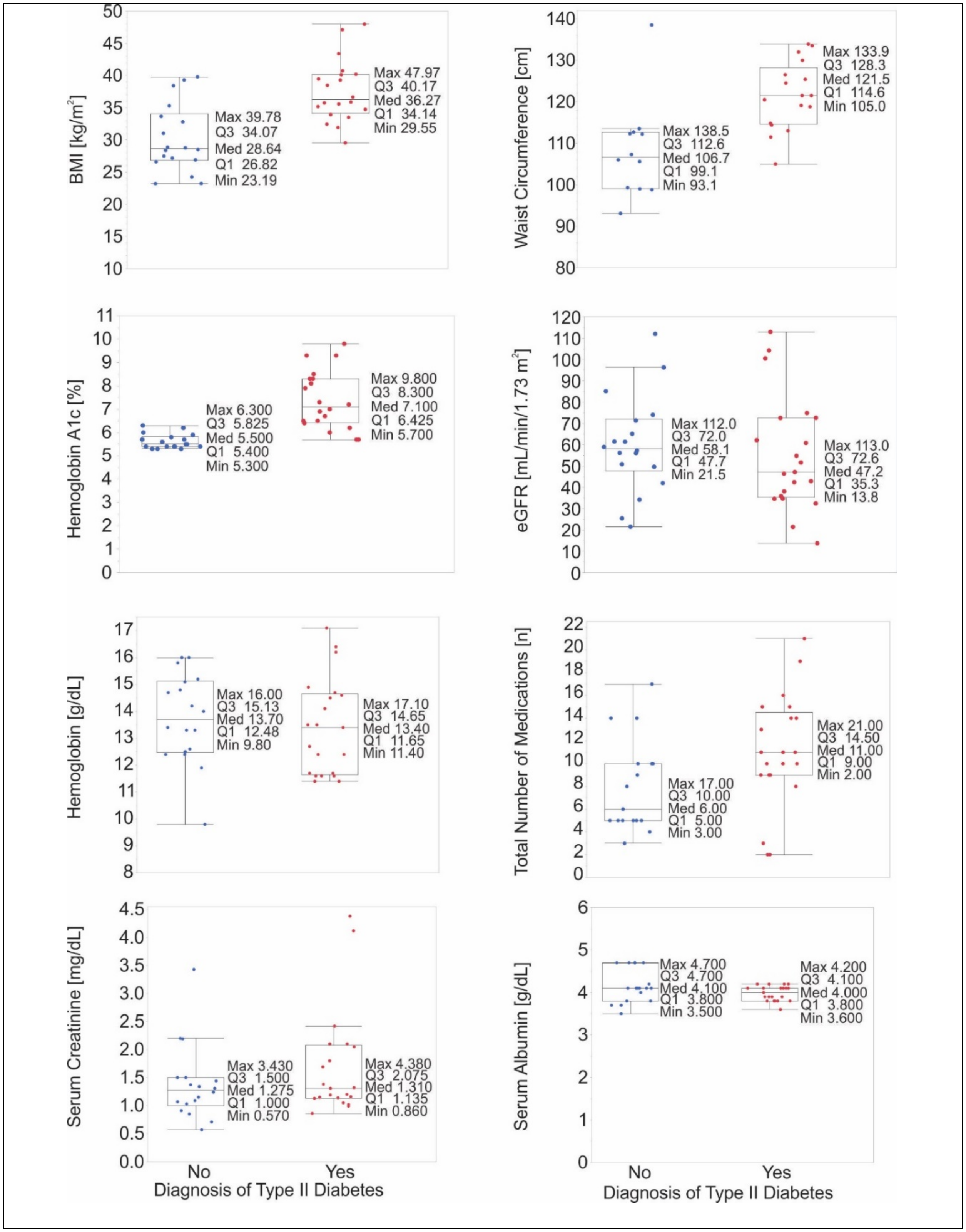

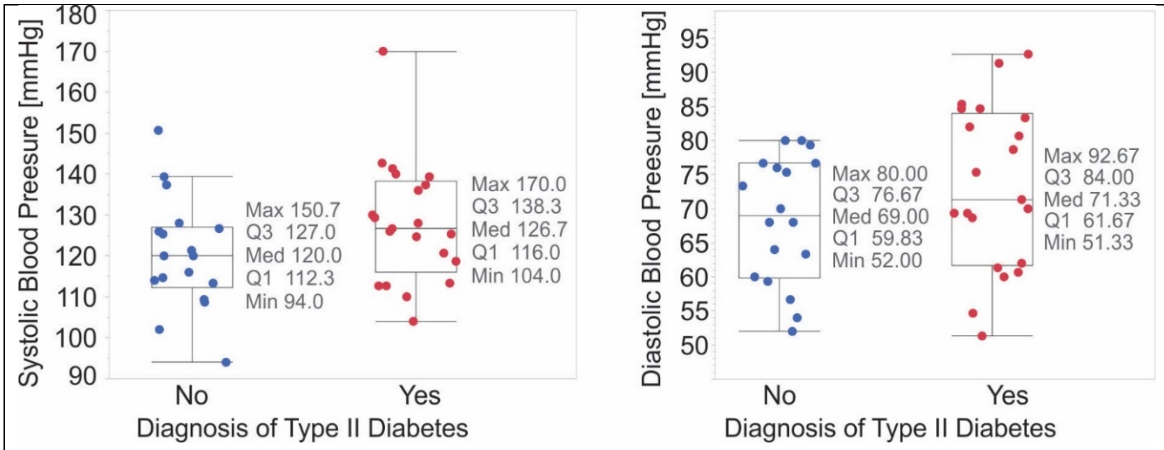


Figure S 2. Clinical & Laboratory Parameters

Figure S 3**.** BMI, waist circumference, hemoglobin A1c, eGFR, hemoglobin, total number of medications taken, serum creatinine, serum albumin and systolic/diastolic blood pressure in patients with (red) and without (blue) a known diagnosis of type 2 diabetes.

**Figure S 4. Variance correlation matrix of BioPatch readouts**

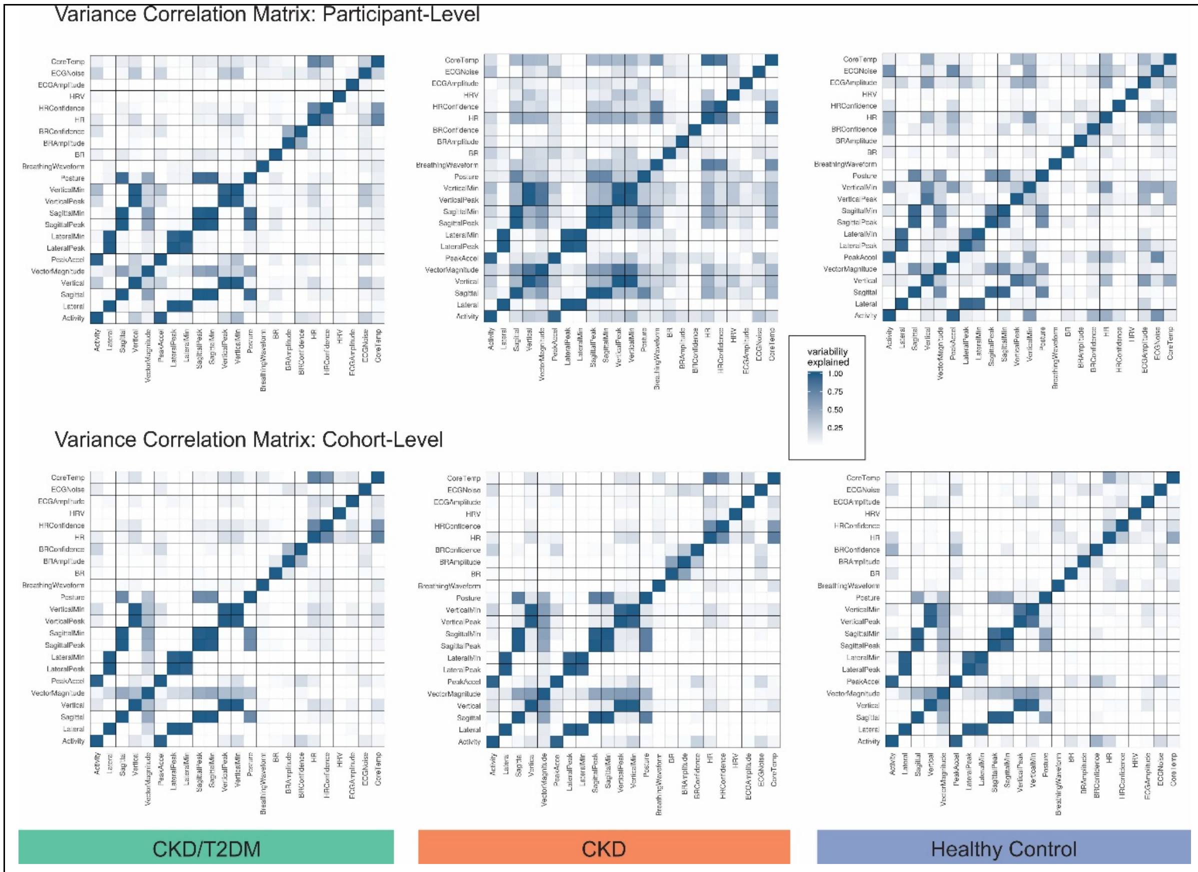


**Figure S 4.** The variance correlation matrices capture how the observed variability is shared among pairs of biometric variables obtained from the BioPatch on the participant-level (top) and the cohort-level (bottom) among CKD/T2DM patients (left), CKD patients (center) and healthy volunteers (right). This allows the examination of circadian organization among biometric variables based on the concept that two variables with a similar temporal pattern share a high degree of variance.

**Figure S 5. Hierarchical cluster analysis**

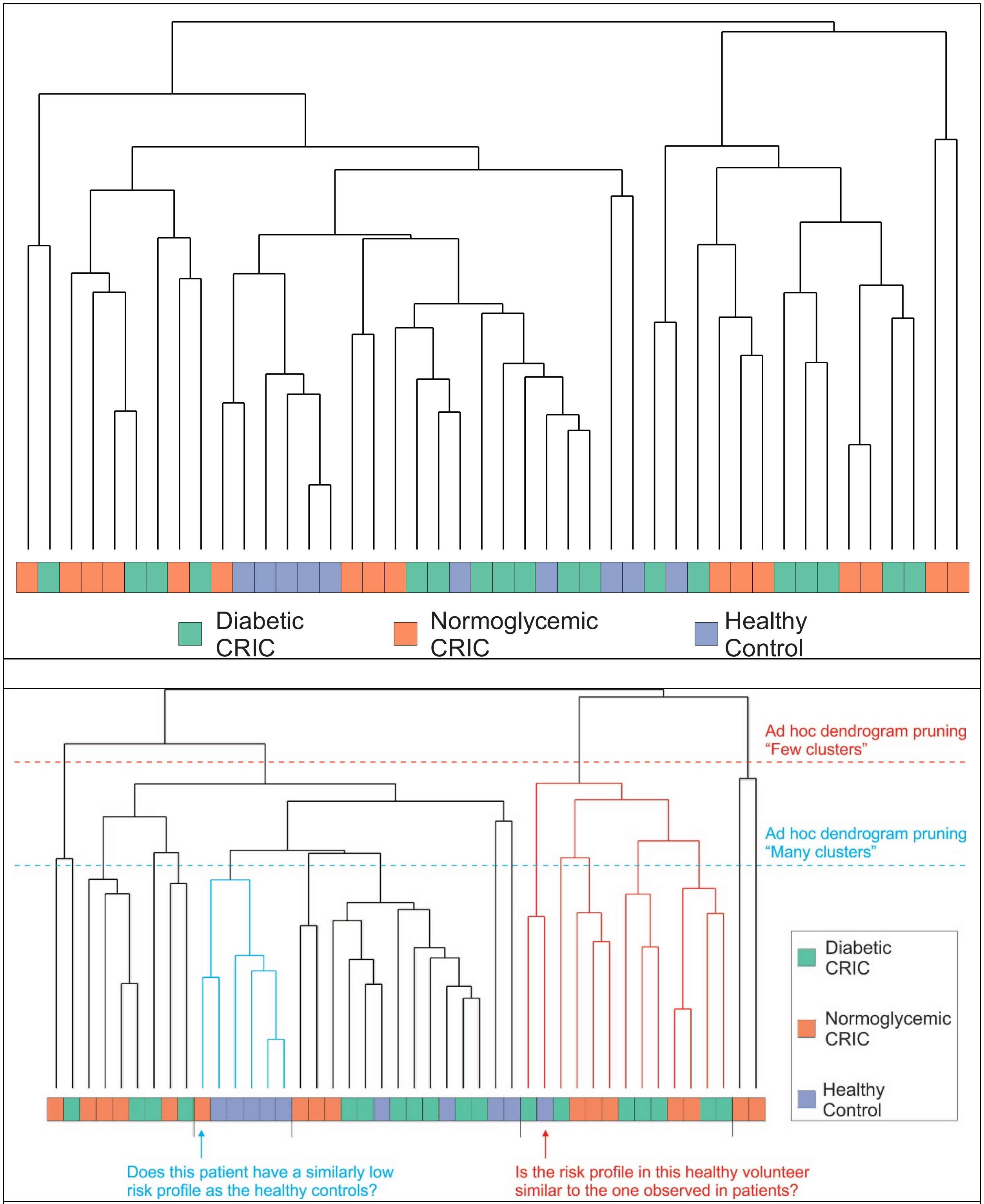


**Figure S 5.** Hierarchical cluster analysis, based on the $R^2$ values derived from the variance correlation metric, produce clusters where members are predominantly controls or a mix of

CKD/T2DM and CKD patients (top). Dendrogram pruning (bottom) to result in few (orange dashed line) and many clusters (blue dashed line) leverages these data to develop hypotheses.

**Figure S 6. Time-versus-subject contribution to variance analysis**

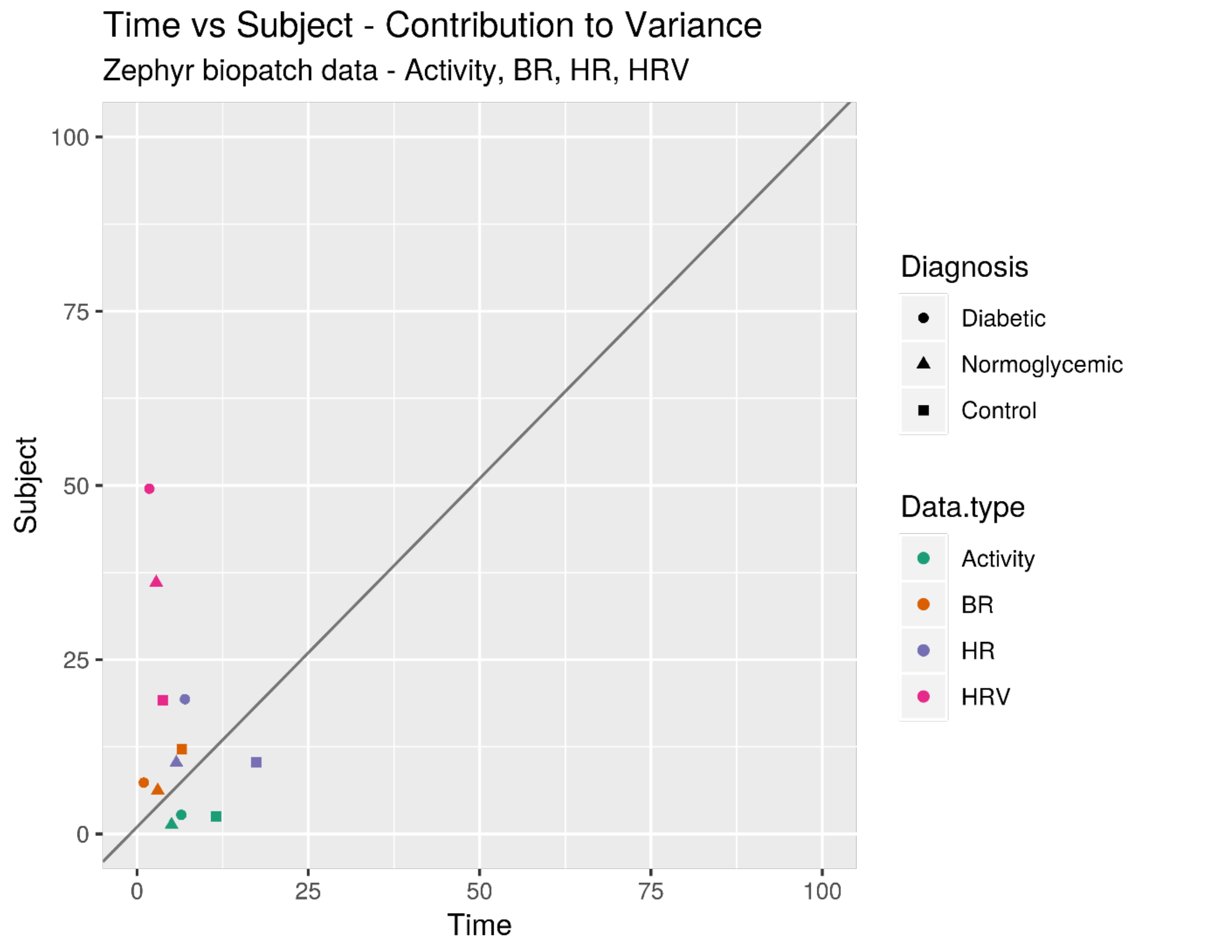


**Figure S 6.** This time-versus-subject contribution to variance analysis visualizes how much of the overall variability is contributed by variability observed in time of biometric measurement versus variability attributable to the inter-subject differences. Units are percent.

**Figure S 7. Distribution of acrophases for activity, breath and heart rate and HRV-SDNN**

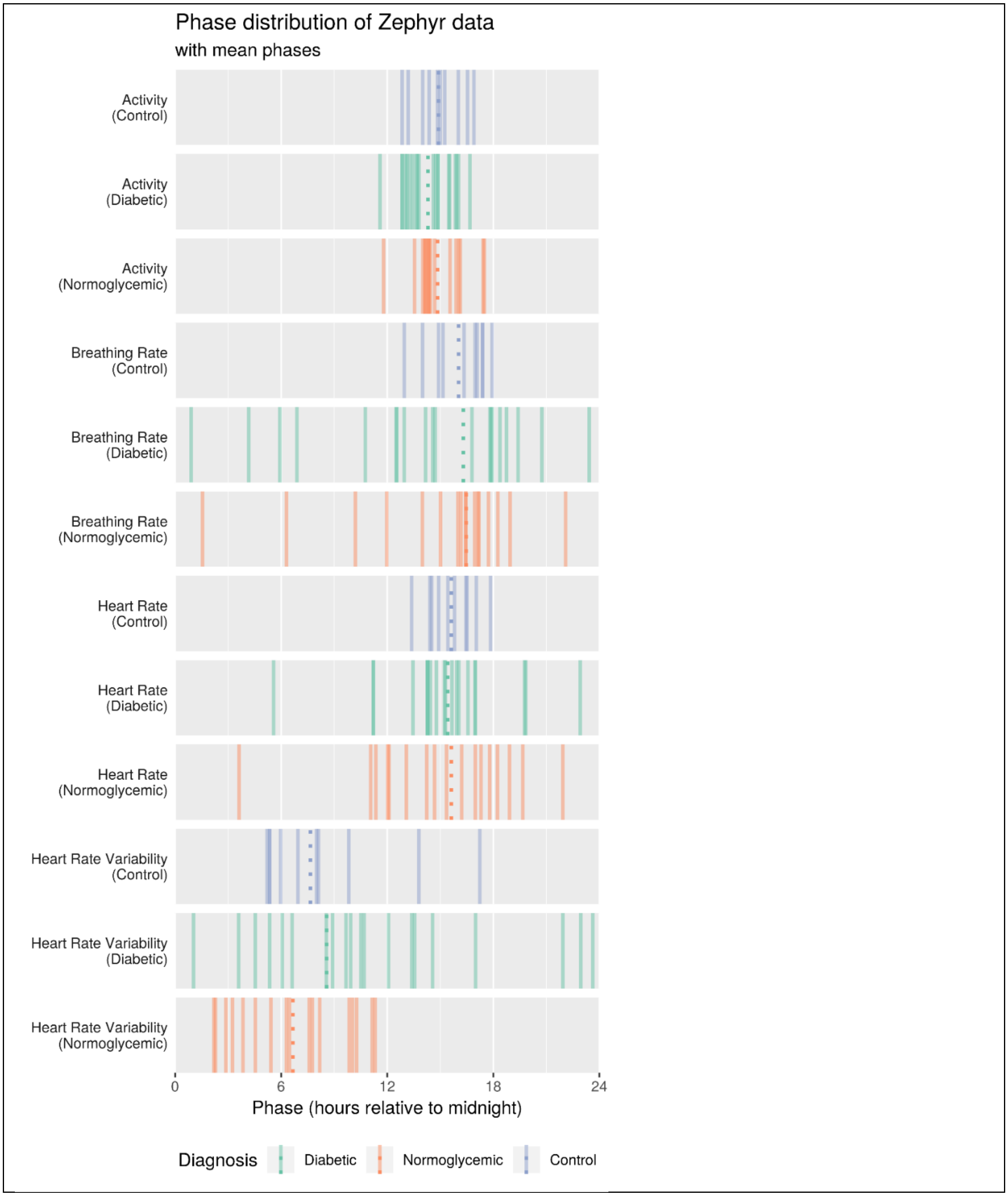

**Figure S 7.** The distribution of acrophases for activity, breath and heart rate and HRV-SDNN indicates the time of day (HH) when physiological readout peaks on the participant level (lines) and cohort-level (dashed line).

#### Figure S 8. Phase relationships between activity versus breath and heart rate and HRV-SDNN

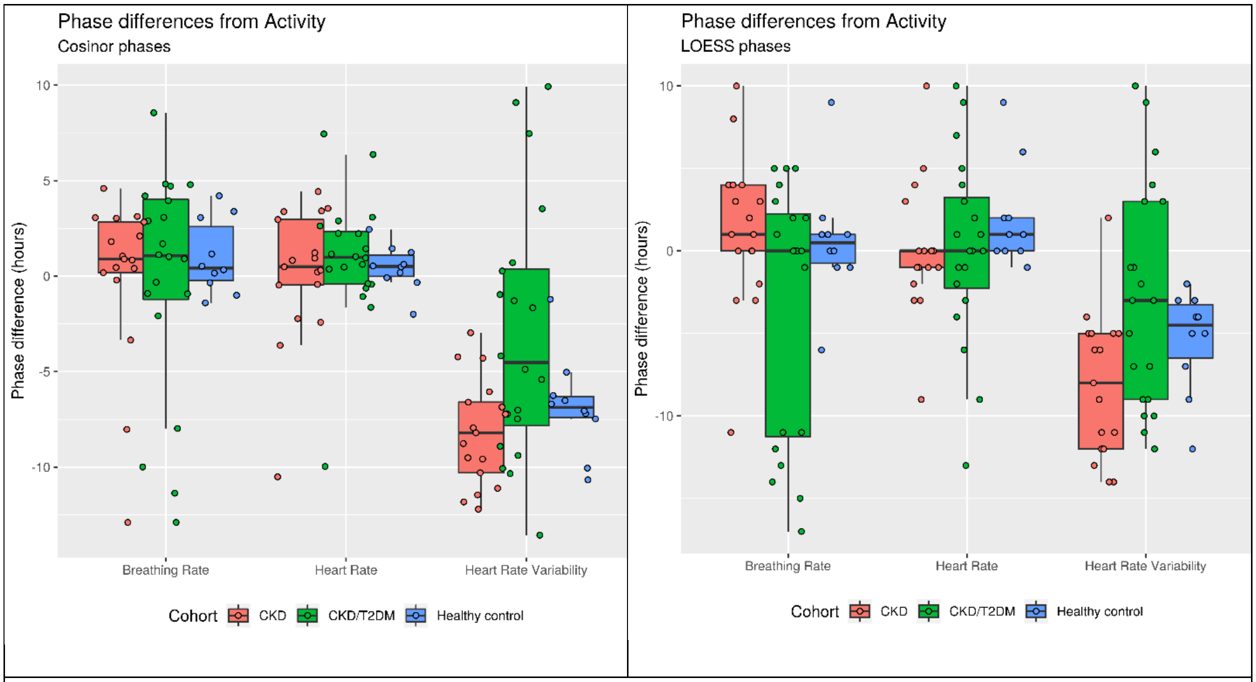


**Figure S 8.** Using the physical activity phenotype as a reference, the phase relationships between activity versus breath and heart rate and HRV-SDNN is shown as the phase difference in hours for the three study cohorts. Cosinor analysis (left) and the more stringent weighted smoothing method, LOESS, (right) were used to determine acrophases.

**Figure S 9. Variance correlation matrix of Kubios readouts: Day versus night**

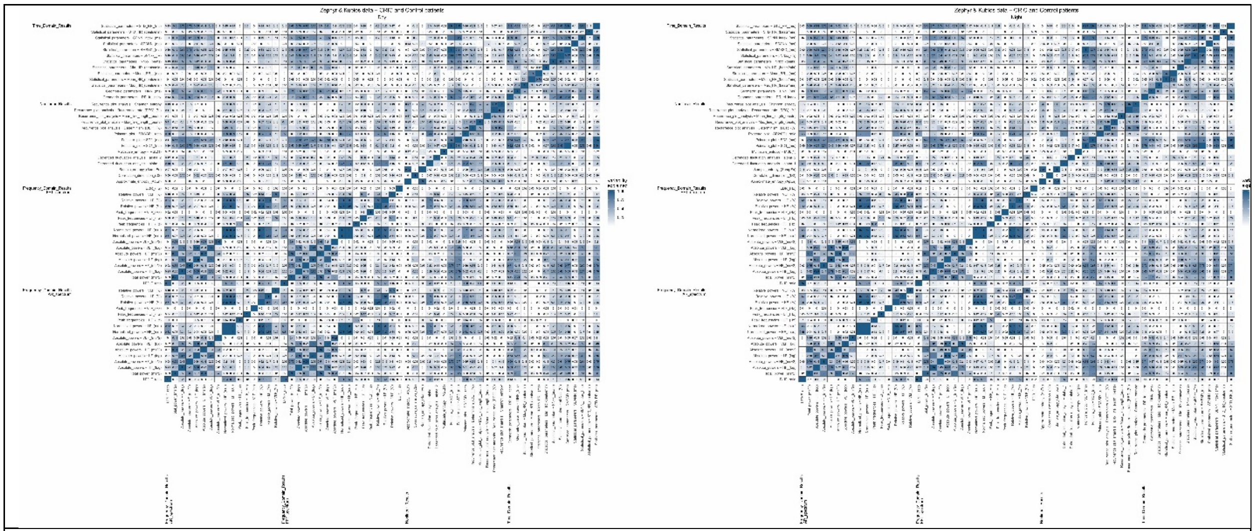

**Figure S 9.** The variance correlation matrices capture how the observed variability is shared among pairs of biometric variables from the Kubios heart rate variability analysis among all participants (n=47) for day (left) versus night (right). This allows the examination of circadian organization among biometric variables based on the concept that two variables with a similar temporal pattern share a high degree of variance.

**Figure S 10. Variance correlation matrix of Kubios readouts: Cohort-level, day versus night**

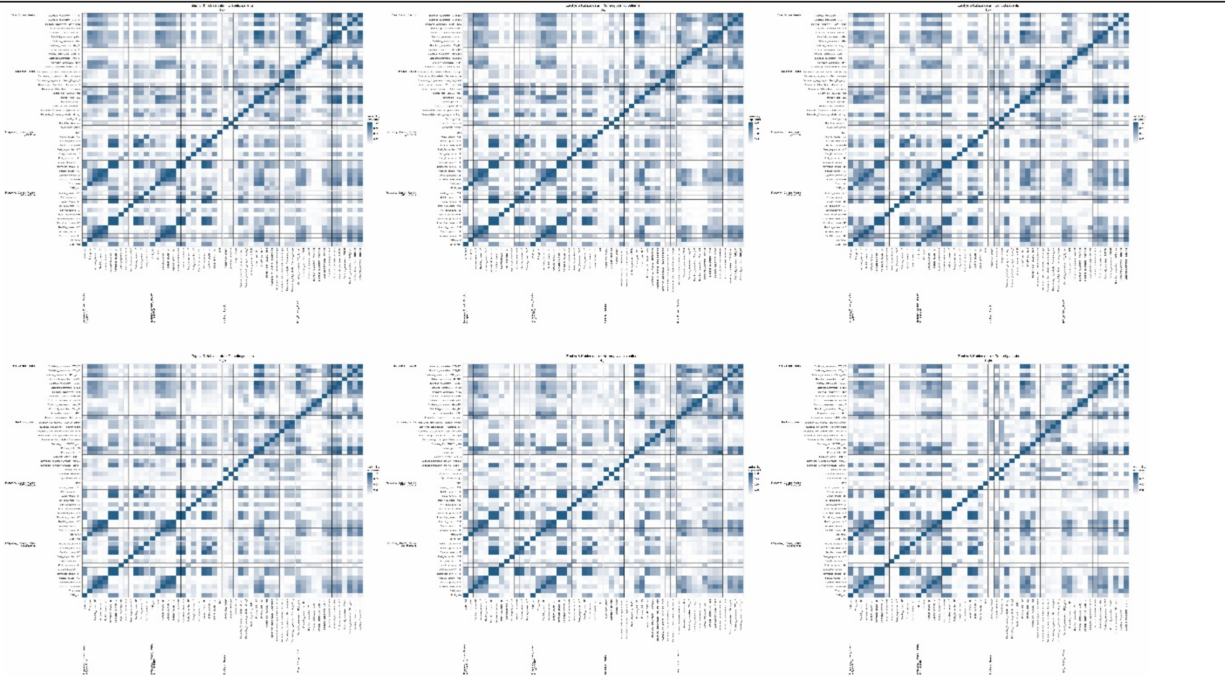

**Figure** S 10**.** The variance correlation matrices capture how the observed variability is shared among pairs of biometric variables from the Kubios heart rate variability analysis on the cohort-level among CKD/T2DM patients (left), CKD patients (center) and healthy volunteers (right) for day (top) versus night (bottom). This allows the examination of circadian organization among biometric variables based on the concept that two variables with a similar temporal pattern share a high degree of variance.

**Figure S 11. Assessment of the outlierness for SDNN**

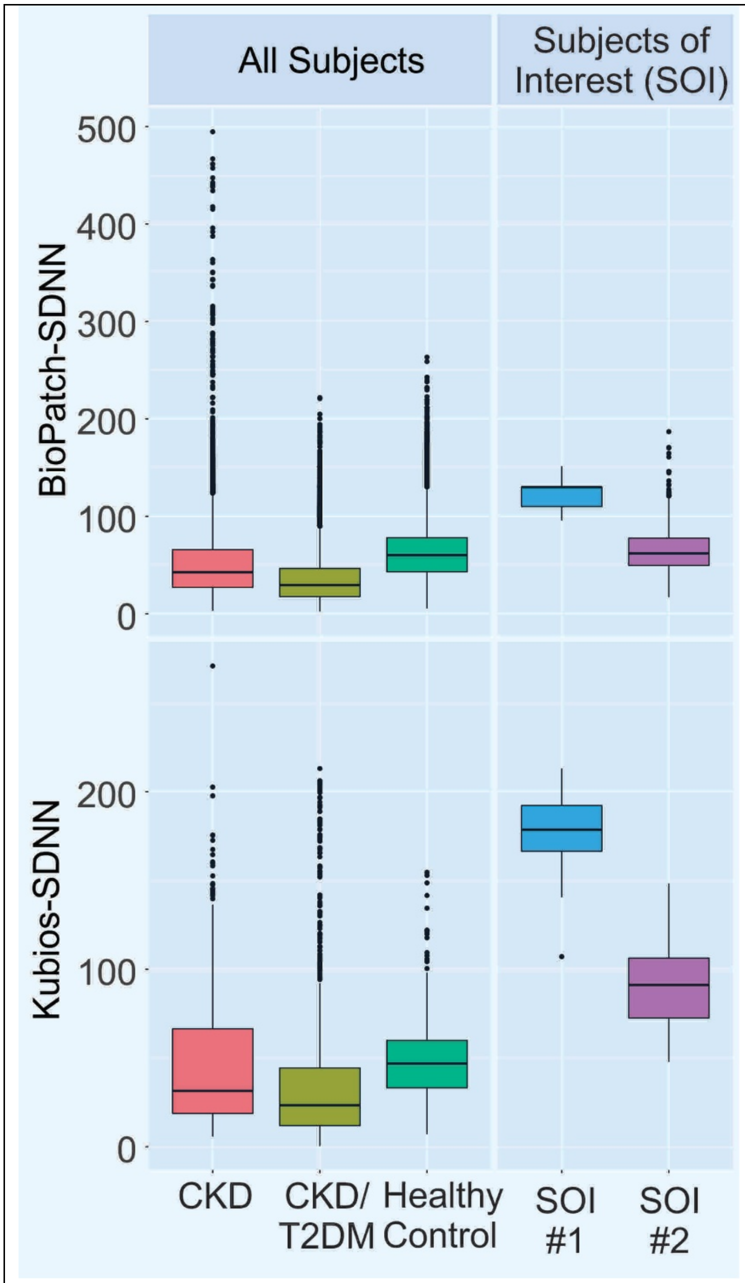


Figure S 11. Comparison of the BioPatch-SDNN (top) and Kubios-SDNN (bottom) for central tendencies in CKD (red), CKD/T2DM (mustard) and controls (green) versus two patients (blue, magenta) from the Penn cohort.

##### Table S 1. Demographics

| Cohort | n | Age [mean] | Age [SD] | Sex (Female) | Ethnicity-Cau | Ethnicity-AA | Ethnicity-Asian |
|---|---|---|---|---|---|---|---|
| **CKD** | 18 | 65.4 | 7.60 | 7 | 13 | 5 | 0 |
| **CKD/T2DM** | 21 | 62.7 | 7.81 | 7 | 9 | 12 | 0 |
| **Control** | 10 | 30.4 | 10.5 | 7 | 8 | 0 | 2 |

##### Table S 2. Clinical & Laboratory Parameters

| Type 2 Diabetes | No | | Yes | |
|---|---|---|---|---|
| | n | Mean±SD | n | Mean±SD |
| **BMI [kg/cm$^2$]** | 18 | 30.2±5.3 | 20 | 37.6±4.8 |
| **Waist Circumference [cm]** | 12 | 108.2±11.6 | 17 | 121.5±8.2 |
| **eGFR [mL/min/1.73 m$^2$]** | 18 | 59.9±22.9 | 21 | 55.1±26.7 |
| **HbA1c [%]** | 18 | 5.6±0.3 | 20 | 7.4±1.2 |
| **Hb [mg/dL]** | 18 | 13.7±1.7 | 21 | 13.4±1.8 |
| **Number of total medications [n]** | 17 | 7.9±4.1 | 21 | 11±5 |
| **Serum Creatinine [mg/dL]** | 18 | 1.4±0.7 | 21 | 1.7±1 |
| **Serum Albumin [g/dL]** | 18 | 4.2±0.4 | 21 | 4±0.2 |
| **Systolic Blood Pressure [mmHg]** | 18 | 120.4±13.6 | 21 | 128±14.7 |
| **Diastolic Blood Pressure [mmHg]** | 18 | 68.5±9.3 | 21 | 73.2±12 |

##### Table S 3. Patient heterogeneity

| Metric | Group 1 (n=3) | Group 2 (n=5) | Group 3 (n=18) | Group 4 (n=7) | Group 5 (n=6) | Incidence |
|---|---|---|---|---|---|---|
| **hypertension** | 3 | 5 | 14 | 7 | 4 | 33 |
| **ALCOH_USE** | 3 | 4 | 15 | 5 | 4 | 31 |
| **lipid** | 2 | 4 | 12 | 5 | 4 | 27 |
| **statins** | 2 | 4 | 12 | 5 | 4 | 27 |
| **acearb** | 2 | 3 | 12 | 6 | 3 | 26 |
| **nsaid** | 1 | 2 | 15 | 4 | 4 | 26 |
| **antiplatelet** | 0 | 1 | 12 | 5 | 3 | 21 |
| **diabetes** | 0 | 4 | 8 | 5 | 4 | 21 |
| **ANTIPLATE_COX1** | 0 | 1 | 12 | 4 | 3 | 20 |
| **anyillicit** | 1 | 3 | 8 | 6 | 2 | 20 |
| **aspirin** | 0 | 1 | 12 | 4 | 3 | 20 |
| **angioblk** | 1 | 3 | 7 | 4 | 2 | 17 |
| **calciferols** | 1 | 4 | 9 | 0 | 3 | 17 |
| **betablk** | 1 | 2 | 6 | 4 | 2 | 15 |
| **cablk** | 1 | 1 | 8 | 4 | 0 | 14 |
| **diuretic** | 1 | 3 | 7 | 3 | 0 | 14 |

| | | | | | | |
|---|---|---|---|---|---|---|
| **anycvd** | 0 | 2 | 8 | 2 | 1 | 13 |
| **insulin** | 0 | 2 | 3 | 4 | 2 | 11 |
| **ACE_INH** | 1 | 0 | 5 | 3 | 1 | 10 |
| **antidiabetes** | 0 | 1 | 5 | 3 | 1 | 10 |
| **mirevasc** | 0 | 2 | 6 | 1 | 1 | 10 |
| **alphablk** | 1 | 2 | 3 | 1 | 2 | 9 |
| **antidepressant** | 0 | 1 | 3 | 2 | 3 | 9 |
| **asthma** | 1 | 1 | 5 | 1 | 0 | 8 |
| **LOOP_DIURETIC** | 0 | 3 | 2 | 3 | 0 | 8 |
| **ssri** | 0 | 1 | 3 | 1 | 3 | 8 |
| **sulfonylurea** | 0 | 1 | 4 | 3 | 0 | 8 |
| **allopurinol** | 1 | 2 | 2 | 1 | 1 | 7 |
| **afib** | 1 | 1 | 2 | 2 | 0 | 6 |
| **biguanide** | 0 | 0 | 4 | 1 | 1 | 6 |
| **copd** | 0 | 3 | 2 | 1 | 0 | 6 |
| **steroids** | 0 | 0 | 4 | 1 | 1 | 6 |
| **THIAZ_DIURETIC** | 0 | 0 | 6 | 0 | 0 | 6 |
| **ALDO_ANTAGONIST** | 1 | 1 | 2 | 1 | 0 | 5 |
| **KSPARE_DIURETIC** | 1 | 1 | 2 | 1 | 0 | 5 |
| **antiepileptics** | 0 | 0 | 2 | 2 | 0 | 4 |
| **ORAL_KCL** | 0 | 1 | 3 | 0 | 0 | 4 |
| **OTHER_LIPID** | 0 | 1 | 1 | 2 | 0 | 4 |
| **ANTI_ACIDOSIS** | 0 | 2 | 1 | 0 | 0 | 3 |
| **arthritis** | 0 | 1 | 0 | 0 | 2 | 3 |
| **stroke** | 0 | 0 | 2 | 0 | 1 | 3 |
| **ANTIPLATE_CAMP_CA** | 0 | 0 | 0 | 2 | 0 | 2 |
| **chf** | 0 | 0 | 0 | 2 | 0 | 2 |
| **CMED_PHOSPHATE** | 0 | 0 | 1 | 0 | 1 | 2 |
| **prednisone** | 0 | 0 | 2 | 0 | 0 | 2 |
| **smokenow** | 0 | 1 | 1 | 0 | 0 | 2 |
| **vasodil** | 0 | 1 | 0 | 1 | 0 | 2 |
| **ACTIVE_VITD** | 0 | 1 | 0 | 0 | 0 | 1 |
| **ANTICOAG_THROMBIN** | 0 | 1 | 0 | 0 | 0 | 1 |
| **ANTICOAG_VITAMINK** | 0 | 0 | 1 | 0 | 0 | 1 |
| **CALC_PHOSBINDERS1** | 0 | 0 | 0 | 0 | 1 | 1 |
| **coronary** | 0 | 0 | 0 | 1 | 0 | 1 |
| **NA_K_CITRATE** | 0 | 0 | 0 | 0 | 1 | 1 |
| **PHOSBINDER_BINARY** | 0 | 0 | 0 | 0 | 1 | 1 |
| **serms** | 0 | 1 | 0 | 0 | 0 | 1 |
| **ALPHA_GLUCO_INH** | 0 | 0 | 0 | 0 | 0 | 0 |
| **ALPHA2AG** | 0 | 0 | 0 | 0 | 0 | 0 |
| **ANTICOAG_HEPARIN** | 0 | 0 | 0 | 0 | 0 | 0 |
| **ANTICOAG_OTHERS** | 0 | 0 | 0 | 0 | 0 | 0 |

| | | | | | | |
|---|---|---|---|---|---|---|
| **ANTICOAG_XA** | 0 | 0 | 0 | 0 | 0 | 0 |
| **antidementia** | 0 | 0 | 0 | 0 | 0 | 0 |
| **ANTIPLATE_EPA** | 0 | 0 | 0 | 0 | 0 | 0 |
| **ANTIPLATE_OTHERS** | 0 | 0 | 0 | 0 | 0 | 0 |
| **anxiolytics** | 0 | 0 | 0 | 0 | 0 | 0 |
| **bilesequestrants** | 0 | 0 | 0 | 0 | 0 | 0 |
| **bisphosphonates** | 0 | 0 | 0 | 0 | 0 | 0 |
| **boneri** | 0 | 0 | 0 | 0 | 0 | 0 |
| **CALC_PHOSBINDERS2** | 0 | 0 | 0 | 0 | 0 | 0 |
| **calcimimetic** | 0 | 0 | 0 | 0 | 0 | 0 |
| **digoxin** | 0 | 0 | 0 | 0 | 0 | 0 |
| **DPP4** | 0 | 0 | 0 | 0 | 0 | 0 |
| **erythrostim** | 0 | 0 | 0 | 0 | 0 | 0 |
| **estrogen** | 0 | 0 | 0 | 0 | 0 | 0 |
| **heparin** | 0 | 0 | 0 | 0 | 0 | 0 |
| **meglitinide** | 0 | 0 | 0 | 0 | 0 | 0 |
| **NON_CALC_PHOSBINDERS** | 0 | 0 | 0 | 0 | 0 | 0 |
| **NON_SEL_SRI** | 0 | 0 | 0 | 0 | 0 | 0 |
| **probenecid** | 0 | 0 | 0 | 0 | 0 | 0 |
| **pvd** | 0 | 0 | 0 | 0 | 0 | 0 |
| **tzd** | 0 | 0 | 0 | 0 | 0 | 0 |

##### Table S 4. BioPatch Summary Outputs: Day versus Night

| Measurement | Time of day | Mean | Std Dev |
|---|---|---|---|
| **Activity** | Day | 0.06 | 0.02 |
| **Activity** | Night | 0.02 | 0.01 |
| **BR** | Day | 14.44 | 1.92 |
| **BR** | Night | 12.83 | 2.04 |
| **BRAmplitude** | Day | 3793 | 3381 |
| **BRAmplitude** | Night | 2282 | 1933 |
| **BRConfidence** | Day | 69.33 | 8.19 |
| **BRConfidence** | Night | 76.01 | 6.46 |
| **BreathingWaveform** | Day | 5198013 | 288839 |
| **BreathingWaveform** | Night | 5256492 | 316557 |
| **CoreTemp** | Day | 37.02 | 0.34 |
| **CoreTemp** | Night | 36.70 | 0.48 |
| **EKGAmplitude** | Day | 0.00 | 0.00 |
| **EKGAmplitude** | Night | 0.00 | 0.00 |
| **EKGNoise** | Day | 0.00 | 0.00 |
| **EKGNoise** | Night | 0.00 | 0.00 |
| **HR** | Day | 68.70 | 13.04 |
| **HR** | Night | 55.09 | 16.41 |

| | | | |
|---|---|---|---|
| **HRConfidence** | Day | 80.54 | 16.04 |
| **HRConfidence** | Night | 73.67 | 20.72 |
| **HRV** | Day | 47.77 | 23.17 |
| **HRV** | Night | 50.23 | 23.75 |
| **Lateral** | Day | 24.66 | 0.07 |
| **Lateral** | Night | 24.70 | 0.17 |
| **LateralMin** | Day | -0.09 | 0.07 |
| **LateralMin** | Night | -0.02 | 0.17 |
| **LateralPeak** | Day | 0.05 | 0.07 |
| **LateralPeak** | Night | 0.06 | 0.17 |
| **PeakAccel** | Day | 0.13 | 0.03 |
| **PeakAccel** | Night | 0.06 | 0.02 |
| **Posture** | Day | 38.64 | 16.25 |
| **Posture** | Night | 64.91 | 17.33 |
| **Posture.Original** | Day | -26.45 | 22.02 |
| **Posture.Original** | Night | -35.80 | 37.01 |
| **Sagittal** | Day | 25.02 | 0.23 |
| **Sagittal** | Night | 25.07 | 0.30 |
| **SagittalMin** | Day | 0.26 | 0.24 |
| **SagittalMin** | Night | 0.34 | 0.30 |
| **SagittalPeak** | Day | 0.42 | 0.23 |
| **SagittalPeak** | Night | 0.44 | 0.30 |
| **VectorMagnitude** | Day | 42.61 | 0.21 |
| **VectorMagnitude** | Night | 42.91 | 0.24 |
| **Vertical** | Day | 24.11 | 0.27 |
| **Vertical** | Night | 24.54 | 0.19 |
| **VerticalMin** | Day | -0.65 | 0.28 |
| **VerticalMin** | Night | -0.18 | 0.19 |
| **VerticalPeak** | Day | -0.49 | 0.26 |
| **VerticalPeak** | Night | -0.10 | 0.18 |

##### Table S 5. HRV Kubios Summary Outputs: Day versus Night

| Measurement | Day.Mean | Night.Mean | Day.SD | Night.SD | Day.Min | Night.Min | Day.Max | Night.Max |
|---|---|---|---|---|---|---|---|---|
| **alpha_1...Nonlinear_Results...Detrended_fluctuation_analysis** | 1.16 | 1.03 | 0.20 | 0.22 | 0.59 | 0.42 | 1.66 | 1.68 |
| **alpha_2...Nonlinear_Results...Detrended_fluctuation_analysis** | 0.49 | 0.45 | 0.09 | 0.11 | 0.24 | 0.22 | 0.78 | 0.92 |
| **Approximate_entropy_(ApEn)...Nonlinear_Results** | 1.40 | 1.43 | 0.12 | 0.10 | 0.89 | 0.92 | 1.65 | 1.58 |
| **Correlation_dimension_(D2)...Nonlinear_Results** | 2.50 | 2.48 | 1.60 | 1.54 | 0.01 | 0.02 | 4.92 | 5.03 |
| **Determinism_(DET)_(%)...Nonlinear_Results...Recurrence_plot_analysis** | 97.68 | 97.65 | 1.10 | 0.82 | 93.20 | 94.78 | 99.92 | 99.77 |
| **EDR_(Hz)...Frequency_Domain_Results...FFT_spectrum** | 0.30 | 0.28 | 0.05 | 0.06 | 0.18 | 0.18 | 0.47 | 0.56 |
| **HF_(%)...Frequency_Domain_Results...Relative_powers...AR_spectrum** | 31.91 | 36.83 | 12.12 | 14.45 | 6.74 | 9.28 | 70.87 | 76.05 |

| | | | | | | | | |
|---|---|---|---|---|---|---|---|---|
| **HF_(%)...Frequency_Domain_Results...Relative_powers...FFT_spectrum** | 31.90 | 36.68 | 12.27 | 14.13 | 6.88 | 9.15 | 70.83 | 75.78 |
| **HF_(Hz)...Frequency_Domain_Results...Peak_frequencies...AR_spectrum** | 0.22 | 0.24 | 0.07 | 0.06 | 0.15 | 0.15 | 0.39 | 0.36 |
| **HF_(Hz)...Frequency_Domain_Results...Peak_frequencies...FFT_spectrum** | 0.20 | 0.22 | 0.07 | 0.06 | 0.15 | 0.15 | 0.35 | 0.35 |
| **HF_(log)...Frequency_Domain_Results...Absolute_powers...AR_spectrum** | 6.29 | 6.44 | 1.13 | 1.22 | 2.16 | 2.56 | 9.35 | 9.86 |
| **HF_(log)...Frequency_Domain_Results...Absolute_powers...FFT_spectrum** | 6.31 | 6.45 | 1.15 | 1.23 | 2.09 | 2.45 | 9.29 | 9.84 |
| **HF_(ms^2)...Frequency_Domain_Results...Absolute_powers...AR_spectrum** | 964.33 | 1182.59 | 1358.83 | 1839.21 | 8.68 | 12.96 | 11484.59 | 19154.87 |
| **HF_(ms^2)...Frequency_Domain_Results...Absolute_powers...FFT_spectrum** | 1003.14 | 1180.95 | 1435.42 | 1793.69 | 8.08 | 11.61 | 10803.11 | 18681.74 |
| **HF_(n.u.)...Frequency_Domain_Results...Normalized_powers...AR_spectrum** | 35.39 | 41.00 | 12.36 | 15.07 | 9.38 | 11.83 | 75.20 | 81.26 |
| **HF_(n.u.)...Frequency_Domain_Results...Normalized_powers...FFT_spectrum** | 36.32 | 41.95 | 13.08 | 15.36 | 9.31 | 11.52 | 76.13 | 81.33 |
| **LF_(%)...Frequency_Domain_Results...Relative_powers...AR_spectrum** | 57.11 | 51.97 | 10.05 | 12.12 | 23.29 | 17.52 | 79.47 | 76.73 |
| **LF_(%)...Frequency_Domain_Results...Relative_powers...FFT_spectrum** | 55.01 | 49.99 | 10.48 | 12.48 | 22.10 | 17.37 | 76.19 | 73.62 |
| **LF_(Hz)...Frequency_Domain_Results...Peak_frequencies...AR_spectrum** | 0.07 | 0.06 | 0.02 | 0.02 | 0.04 | 0.04 | 0.13 | 0.13 |
| **LF_(Hz)...Frequency_Domain_Results...Peak_frequencies...FFT_spectrum** | 0.09 | 0.08 | 0.02 | 0.02 | 0.04 | 0.04 | 0.15 | 0.15 |
| **LF_(log)...Frequency_Domain_Results...Absolute_powers...AR_spectrum** | 6.93 | 6.85 | 0.75 | 0.84 | 3.48 | 4.49 | 9.10 | 9.74 |
| **LF_(log)...Frequency_Domain_Results...Absolute_powers...FFT_spectrum** | 6.91 | 6.81 | 0.75 | 0.82 | 3.50 | 4.43 | 9.06 | 9.63 |
| **LF_(ms^2)...Frequency_Domain_Results...Absolute_powers...AR_spectrum** | 1359.97 | 1362.64 | 1217.35 | 1696.20 | 32.35 | 89.11 | 8915.59 | 17066.74 |
| **LF_(ms^2)...Frequency_Domain_Results...Absolute_powers...FFT_spectrum** | 1331.47 | 1287.99 | 1212.62 | 1552.83 | 33.03 | 83.67 | 8638.79 | 15288.44 |
| **LF_(n.u.)...Frequency_Domain_Results...Normalized_powers...AR_spectrum** | 64.50 | 58.91 | 12.42 | 15.09 | 24.72 | 18.72 | 90.51 | 88.10 |
| **LF_(n.u.)...Frequency_Domain_Results...Normalized_powers...FFT_spectrum** | 63.57 | 57.96 | 13.14 | 15.39 | 23.76 | 18.64 | 90.60 | 88.41 |
| **LF/HF_ratio...Frequency_Domain_Results...AR_spectrum** | 2.26 | 1.95 | 1.43 | 1.60 | 0.33 | 0.23 | 9.65 | 7.45 |
| **LF/HF_ratio...Frequency_Domain_Results...FFT_spectrum** | 2.20 | 1.89 | 1.47 | 1.60 | 0.31 | 0.23 | 9.74 | 7.67 |
| **Max_HR_(beats/min)...Time_Domain_Results...Statistical_parameters** | 147.45 | 139.28 | 53.02 | 56.85 | 62.11 | 68.80 | 240.00 | 240.00 |
| **Max_line_length_(beats)...Nonlinear_Results...Recurrence_plot_analysis** | 255.77 | 213.75 | 122.83 | 107.79 | 44.00 | 66.00 | 869.00 | 782.00 |
| **Mean_HR_(beats/min)...Time_Domain_Results...Statistical_parameters** | 79.32 | 69.95 | 18.84 | 23.68 | 41.48 | 44.73 | 182.85 | 188.78 |
| **Mean_line_length_(beats)...Nonlinear_Results...Recurrence_plot_analysis** | 12.72 | 13.46 | 5.96 | 5.39 | 5.90 | 7.55 | 69.39 | 42.28 |
| **Mean_RR__(ms)...Time_Domain_Results...Statistical_parameters** | 800.30 | 915.17 | 202.40 | 191.94 | 328.13 | 317.83 | 1446.62 | 1341.50 |
| **Min_HR_(beats/min)...Time_Domain_Results...Statistical_parameters** | 56.69 | 50.77 | 11.24 | 9.33 | 29.53 | 29.29 | 99.25 | 97.10 |
| **MSE(1)...Nonlinear_Results...Multiscale_entropy** | 1.39 | 1.33 | 0.41 | 0.46 | 0.26 | 0.26 | 2.04 | 1.82 |
| **MSE(10)...Nonlinear_Results...Multiscale_entropy** | 0.94 | 0.88 | 0.26 | 0.27 | 0.27 | 0.27 | 1.52 | 1.52 |
| **MSE(11)...Nonlinear_Results...Multiscale_entropy** | 0.88 | 0.82 | 0.25 | 0.26 | 0.28 | 0.28 | 1.50 | 1.50 |
| **MSE(12)...Nonlinear_Results...Multiscale_entropy** | 0.81 | 0.76 | 0.23 | 0.24 | 0.25 | 0.25 | 1.43 | 1.43 |
| **MSE(13)...Nonlinear_Results...Multiscale_entropy** | 0.76 | 0.71 | 0.20 | 0.19 | 0.28 | 0.28 | 1.44 | 1.44 |
| **MSE(14)...Nonlinear_Results...Multiscale_entropy** | 0.72 | 0.68 | 0.19 | 0.20 | 0.26 | 0.26 | 1.36 | 1.36 |
| **MSE(15)...Nonlinear_Results...Multiscale_entropy** | 0.68 | 0.65 | 0.16 | 0.17 | 0.25 | 0.25 | 1.48 | 1.48 |

| | | | | | | | | |
|---|---|---|---|---|---|---|---|---|
| **MSE(16)...Nonlinear_Results...Multiscale_entropy** | 0.64 | 0.61 | 0.15 | 0.16 | 0.23 | 0.23 | 1.23 | 1.23 |
| **MSE(17)...Nonlinear_Results...Multiscale_entropy** | 0.61 | 0.57 | 0.16 | 0.16 | 0.25 | 0.25 | 1.19 | 1.19 |
| **MSE(18)...Nonlinear_Results...Multiscale_entropy** | 0.57 | 0.55 | 0.15 | 0.16 | 0.24 | 0.24 | 1.09 | 1.09 |
| **MSE(19)...Nonlinear_Results...Multiscale_entropy** | 0.55 | 0.54 | 0.13 | 0.14 | 0.28 | 0.28 | 1.20 | 1.20 |
| **MSE(2)...Nonlinear_Results...Multiscale_entropy** | 1.46 | 1.39 | 0.42 | 0.47 | 0.27 | 0.27 | 1.87 | 1.87 |
| **MSE(20)...Nonlinear_Results...Multiscale_entropy** | 0.52 | 0.50 | 0.13 | 0.12 | 0.27 | 0.27 | 1.08 | 1.08 |
| **MSE(3)...Nonlinear_Results...Multiscale_entropy** | 1.38 | 1.30 | 0.40 | 0.45 | 0.22 | 0.22 | 1.75 | 1.75 |
| **MSE(4)...Nonlinear_Results...Multiscale_entropy** | 1.36 | 1.29 | 0.39 | 0.43 | 0.25 | 0.25 | 1.88 | 1.85 |
| **MSE(5)...Nonlinear_Results...Multiscale_entropy** | 1.32 | 1.25 | 0.37 | 0.40 | 0.28 | 0.28 | 1.72 | 1.72 |
| **MSE(6)...Nonlinear_Results...Multiscale_entropy** | 1.24 | 1.18 | 0.35 | 0.38 | 0.30 | 0.30 | 1.68 | 1.68 |
| **MSE(7)...Nonlinear_Results...Multiscale_entropy** | 1.18 | 1.12 | 0.32 | 0.36 | 0.29 | 0.29 | 1.63 | 1.63 |
| **MSE(8)...Nonlinear_Results...Multiscale_entropy** | 1.08 | 1.02 | 0.31 | 0.32 | 0.29 | 0.29 | 1.49 | 1.47 |
| **MSE(9)...Nonlinear_Results...Multiscale_entropy** | 1.00 | 0.93 | 0.27 | 0.28 | 0.27 | 0.27 | 1.54 | 1.54 |
| **NNxx_(beats)...Time_Domain_Results...Statistical_parameters** | 905.05 | 999.60 | 928.79 | 1070.91 | 0.00 | 2.00 | 5622.00 | 6175.00 |
| **pNNxx_(%)...Time_Domain_Results...Statistical_parameters** | 19.25 | 24.01 | 16.90 | 20.39 | 0.00 | 0.03 | 75.32 | 72.85 |
| **Recurrence_rate_(REC)_(%)...Nonlinear_Results...Recurrence_plot_analysis** | 33.14 | 33.74 | 8.27 | 7.33 | 15.17 | 19.67 | 85.64 | 55.97 |
| **RMSSD_(ms)...Time_Domain_Results...Statistical_parameters** | 47.80 | 53.70 | 32.54 | 32.77 | 4.06 | 3.91 | 210.77 | 177.53 |
| **RR_tri_index...Time_Domain_Results...Geometric_parameters** | 11.86 | 11.87 | 5.01 | 4.66 | 2.10 | 2.75 | 35.78 | 23.77 |
| **Sample_entropy_(SampEn)...Nonlinear_Results** | 1.52 | 1.59 | 0.22 | 0.18 | 0.73 | 0.81 | 2.04 | 1.91 |
| **SD1_(ms)...Nonlinear_Results...Poincare_plot** | 33.80 | 37.98 | 23.01 | 23.17 | 2.87 | 2.77 | 149.05 | 125.55 |
| **SD2_(ms)...Nonlinear_Results...Poincare_plot** | 62.11 | 61.36 | 26.24 | 26.58 | 9.58 | 15.59 | 174.82 | 176.47 |
| **SD2/SD1_ratio...Nonlinear_Results...Poincare_plot** | 2.11 | 1.86 | 0.55 | 0.58 | 0.96 | 0.74 | 4.00 | 5.63 |
| **SDANN_(ms)...Time_Domain_Results...Statistical_parameters** | 73.20 | 69.87 | 58.61 | 73.60 | 7.34 | 5.53 | 513.08 | 407.98 |
| **SDNN_index_(ms)...Time_Domain_Results...Statistical_parameters** | 76.14 | 76.12 | 27.48 | 28.69 | 16.54 | 21.35 | 189.93 | 165.80 |
| **Shannon_entropy...Nonlinear_Results...Recurrence_plot_analysis** | 3.20 | 3.28 | 0.28 | 0.23 | 2.47 | 2.70 | 4.94 | 4.11 |
| **STD_HR_(beats/min)...Time_Domain_Results...Statistical_parameters** | 20.90 | 61.47 | 77.61 | 534.99 | 0.90 | 1.19 | 636.18 | 6633.83 |
| **STD_RR_(ms)...Time_Domain_Results...Statistical_parameters** | 50.35 | 51.42 | 23.97 | 24.11 | 7.07 | 11.19 | 154.62 | 148.69 |
| **TINN_(ms)...Time_Domain_Results...Geometric_parameters** | 430.14 | 457.72 | 276.63 | 286.26 | 72.00 | 122.00 | 1534.00 | 1797.00 |
| **Total_power_(ms^2)...Frequency_Domain_Results...AR_spectrum** | 2568.78 | 2831.36 | 2615.94 | 3801.41 | 47.14 | 123.09 | 19907.08 | 39922.83 |
| **Total_power_(ms^2)...Frequency_Domain_Results...FFT_spectrum** | 2656.92 | 2847.56 | 2788.33 | 3811.94 | 48.49 | 120.65 | 19979.16 | 39680.25 |
| **VLF_(%)...Frequency_Domain_Results...Relative_powers...AR_spectrum** | 10.89 | 11.12 | 6.30 | 5.01 | 2.89 | 3.09 | 38.58 | 24.53 |
| **VLF_(%)...Frequency_Domain_Results...Relative_powers...FFT_spectrum** | 13.00 | 13.25 | 4.91 | 4.56 | 6.22 | 5.76 | 31.62 | 26.62 |
| **VLF_(Hz)...Frequency_Domain_Results...Peak_frequencies...AR_spectrum** | 0.04 | 0.04 | 0.00 | 0.00 | 0.03 | 0.03 | 0.04 | 0.04 |
| **VLF_(Hz)...Frequency_Domain_Results...Peak_frequencies...FFT_spectrum** | 0.04 | 0.04 | 0.00 | 0.01 | 0.00 | 0.00 | 0.04 | 0.04 |
| **VLF_(log)...Frequency_Domain_Results...Absolute_powers...AR_spectrum** | 5.18 | 5.24 | 0.78 | 0.81 | 1.80 | 3.04 | 7.46 | 8.24 |
| **VLF_(log)...Frequency_Domain_Results...Absolute_powers...FFT_spectrum** | 5.43 | 5.46 | 0.80 | 0.88 | 1.99 | 3.23 | 7.91 | 8.64 |

| | | | | | | | | |
|---|---|---|---|---|---|---|---|---|
| **VLF_(ms^2)...Frequency_Domain_Results...Absolute_powers...AR_spectrum** | 240.89 | 282.78 | 240.42 | 437.50 | 6.08 | 21.01 | 1728.52 | 3790.77 |
| **VLF_(ms^2)...Frequency_Domain_Results...Absolute_powers...FFT_spectrum** | 318.59 | 375.35 | 334.96 | 604.08 | 7.35 | 25.35 | 2726.74 | 5639.96 |

### Table S 6. BioPatch Summary Outputs: Day versus Night by Cohort

| Condition | | CKD/T2DM | CKD/T2DM | CKD | CKD | Control | Control |
|---|---|---|---|---|---|---|---|
| Measurement | **Time_of_day** | **Mean** | **Std Dev** | **Mean** | **Std Dev** | **Mean** | **Std Dev** |
| **Activity** | Day | 0.050 | 0.010 | 0.058 | 0.012 | 0.080 | 0.020 |
| **Activity** | Night | 0.021 | 0.007 | 0.022 | 0.006 | 0.026 | 0.009 |
| **BR** | Day | 13.7 | 1.5 | 14.9 | 2.3 | 15.0 | 1.6 |
| **BR** | Night | 13.1 | 1.9 | 12.7 | 2.3 | 12.4 | 2.1 |
| **BRAmplitude** | Day | 4061.9 | 2614.9 | 4109.5 | 4804.0 | 2785.4 | 1123.1 |
| **BRAmplitude** | Night | 2924.3 | 2411.3 | 1764.8 | 1640.2 | 1981.7 | 741.7 |
| **BRConfidence** | Day | 68.6 | 7.8 | 71.1 | 9.8 | 67.2 | 6.6 |
| **BRConfidence** | Night | 74.9 | 7.1 | 77.6 | 5.2 | 75.9 | 6.3 |
| **BreathingWaveform** | Day | 5287673.4 | 295854.3 | 5277585.4 | 206550.7 | 4889735.6 | 166313.0 |
| **BreathingWaveform** | Night | 5335222.1 | 284366.6 | 5306605.0 | 301096.4 | 4998843.8 | 288185.3 |
| **CoreTemp** | Day | 37.0 | 0.4 | 36.9 | 0.3 | 37.2 | 0.2 |
| **CoreTemp** | Night | 36.8 | 0.6 | 36.7 | 0.4 | 36.7 | 0.3 |
| **EKGAmplitude** | Day | 0.00158 | 0.00074 | 0.00162 | 0.00066 | 0.00191 | 0.00072 |
| **EKGAmplitude** | Night | 0.00134 | 0.00064 | 0.00138 | 0.00049 | 0.00162 | 0.00060 |
| **EKGNoise** | Day | 0.00009 | 0.00004 | 0.00010 | 0.00005 | 0.00015 | 0.00005 |
| **EKGNoise** | Night | 0.00007 | 0.00004 | 0.00006 | 0.00002 | 0.00009 | 0.00005 |
| **HR** | Day | 71.4 | 14.5 | 61.8 | 9.8 | 74.4 | 8.2 |
| **HR** | Night | 58.1 | 20.7 | 51.7 | 11.8 | 55.6 | 10.2 |
| **HRConfidence** | Day | 80.9 | 15.1 | 75.0 | 18.2 | 89.3 | 7.8 |
| **HRConfidence** | Night | 72.8 | 18.0 | 73.7 | 19.6 | 84.2 | 10.2 |
| **HRV** | Day | 38.6 | 24.4 | 49.7 | 22.0 | 63.3 | 13.1 |
| **HRV** | Night | 39.1 | 23.8 | 53.9 | 22.5 | 65.5 | 15.5 |
| **Lateral** | Day | 24.6 | 0.1 | 24.7 | 0.0 | 24.7 | 0.0 |
| **Lateral** | Night | 24.7 | 0.2 | 24.7 | 0.1 | 24.7 | 0.3 |
| **LateralMin** | Day | -0.09 | 0.10 | -0.08 | 0.05 | -0.07 | 0.07 |
| **LateralMin** | Night | -0.05 | 0.15 | -0.01 | 0.13 | 0.00 | 0.28 |
| **LateralPeak** | Day | 0.04 | 0.10 | 0.06 | 0.04 | 0.09 | 0.06 |
| **LateralPeak** | Night | 0.03 | 0.16 | 0.07 | 0.13 | 0.08 | 0.28 |
| **PeakAccel** | Day | 0.11 | 0.02 | 0.13 | 0.03 | 0.17 | 0.04 |
| **PeakAccel** | Night | 0.06 | 0.01 | 0.06 | 0.01 | 0.06 | 0.02 |
| **Posture** | Day | -29.4 | 26.1 | -26.7 | 16.5 | -20.5 | 23.3 |
| **Posture** | Night | -32.4 | 43.8 | -41.9 | 35.7 | -35.9 | 24.8 |
| **Sagittal** | Day | 25.1 | 0.2 | 25.0 | 0.2 | 24.9 | 0.2 |
| **Sagittal** | Night | 25.0 | 0.3 | 25.1 | 0.3 | 25.0 | 0.2 |
| **SagittalMin** | Day | 0.34 | 0.24 | 0.28 | 0.19 | 0.10 | 0.24 |

| | | | | | | | |
|---|---|---|---|---|---|---|---|
| **SagittalMin** | Night | 0.33 | 0.33 | 0.42 | 0.31 | 0.29 | 0.19 |
| **SagittalPeak** | Day | 0.48 | 0.25 | 0.43 | 0.19 | 0.29 | 0.25 |
| **SagittalPeak** | Night | 0.41 | 0.33 | 0.51 | 0.32 | 0.39 | 0.19 |
| **VectorMagnitude** | Day | 42.7 | 0.2 | 42.6 | 0.2 | 42.6 | 0.3 |
| **VectorMagnitude** | Night | 42.9 | 0.3 | 42.9 | 0.2 | 42.9 | 0.2 |
| **Vertical** | Day | 24.2 | 0.2 | 24.0 | 0.1 | 24.2 | 0.4 |
| **Vertical** | Night | 24.6 | 0.2 | 24.5 | 0.1 | 24.6 | 0.2 |
| **VerticalMin** | Day | -0.56 | 0.24 | -0.76 | 0.15 | -0.59 | 0.42 |
| **VerticalMin** | Night | -0.16 | 0.24 | -0.21 | 0.12 | -0.15 | 0.15 |
| **VerticalPeak** | Day | -0.43 | 0.22 | -0.60 | 0.13 | -0.37 | 0.40 |
| **VerticalPeak** | Night | -0.08 | 0.23 | -0.13 | 0.12 | -0.06 | 0.15 |

#### Table S 7. HRV Kubios Summary Outputs: Day versus Night by Cohort

| Measurement | Control_Day.Mean | Control_Night.Mean | CKD/T2DM_Day.Mean | CKD/T2DM_Night.Mean | CKD_Day.Mean | CKD_Night.Mean | Control_Day.SD | Control_Night.SD | CKD/T2DM_Day.SD | CKD/T2DM_Night.SD | CKD_Day.SD | CKD_Night.SD |
|---|---|---|---|---|---|---|---|---|---|---|---|---|
| **alpha_1...Nonlinear_Results...Detrended_fluctuation_analysis** | 1.16 | 1.03 | 1.08 | 1.04 | 1.09 | 1.01 | 0.20 | 0.22 | 0.29 | 0.30 | 0.31 | 0.32 |
| alpha_2...Nonlinear_Results...Detrended_fluctuation_analysis | **0.49** | **0.45** | **0.58** | **0.58** | **0.50** | **0.45** | **0.09** | **0.11** | **0.17** | **0.20** | **0.13** | **0.12** |
| **Approximate_entropy_(ApEn)...Nonlinear_Results** | 1.40 | 1.43 | 1.41 | 1.43 | 1.40 | 1.41 | 0.12 | 0.10 | 0.18 | 0.17 | 0.17 | 0.17 |
| Correlation_dimension_(D2)...Nonlinear_Results | **2.50** | **2.48** | **0.58** | **0.94** | **1.05** | **1.33** | **1.60** | **1.54** | **0.93** | **1.41** | **1.27** | **1.29** |
| **Determinism_(DET)_(%)...Nonlinear_Results...Recurrence_plot_analysis** | 97.7 | 97.7 | 97.4 | 97.2 | 97.4 | 97.2 | 1.1 | 0.8 | 1.8 | 1.8 | 1.7 | 1.7 |
| EDR_(Hz)...Frequency_Domain_Results...FFT_spectrum | **0.30** | **0.28** | **0.22** | **0.22** | **0.23** | **0.23** | **0.05** | **0.06** | **0.06** | **0.05** | **0.04** | **0.05** |
| **HF_(%)...Frequency_Domain_Results...Relative_powers...AR_spectrum** | 31.9 | 36.8 | 30.3 | 35.1 | 32.0 | 35.6 | 12.1 | 14.5 | 16.7 | 19.5 | 19.0 | 20.1 |
| **HF_(%)...Frequency_Domain_Results...Relative_powers...FFT_spectrum** | 31.9 | 36.7 | 29.8 | 34.8 | 31.9 | 35.7 | 12.3 | 14.1 | 16.7 | 19.6 | 19.1 | 20.2 |
| **HF_(Hz)...Frequency_Domain_Results...Peak_frequencies...AR_spectrum** | 0.22 | 0.24 | 0.23 | 0.24 | 0.22 | 0.23 | 0.07 | 0.06 | 0.08 | 0.07 | 0.08 | 0.07 |
| **HF_(Hz)...Frequency_Domain_Results...Peak_frequencies...FFT_spectrum** | 0.20 | 0.22 | 0.20 | 0.22 | 0.21 | 0.22 | 0.07 | 0.06 | 0.08 | 0.07 | 0.08 | 0.08 |
| **HF_(log)...Frequency_Domain_Results...Absolute_powers...AR_spectrum** | 6.29 | 6.44 | 4.78 | 5.23 | 5.39 | 6.01 | 1.13 | 1.22 | 2.39 | 2.44 | 2.02 | 1.75 |
| **HF_(log)...Frequency_Domain_Results...Absolute_powers...FFT_spectrum** | 6.31 | 6.45 | 4.76 | 5.22 | 5.38 | 6.03 | 1.15 | 1.23 | 2.39 | 2.45 | 2.02 | 1.73 |
| **HF_(ms^2)...Frequency_Domain_Results...Absolute_powers...AR_spectrum** | 964.3 | 1182.6 | 1815.9 | 1894.7 | 1377.7 | 1572.1 | 1358.8 | 1839.2 | 4767.6 | 4301.5 | 2906.7 | 2577.6 |
| **HF_(ms^2)...Frequency_Domain_Results...Absolute_powers...FFT_spectrum** | 1003.1 | 1180.9 | 1811.0 | 1881.6 | 1336.0 | 1554.0 | 1435.4 | 1793.7 | 4763.8 | 4240.9 | 2684.3 | 2520.3 |
| **HF_(n.u.)...Frequency_Domain_Results...Normalized_powers...AR_spectrum** | 35.39 | 41.00 | 34.78 | 40.47 | 35.84 | 40.00 | 12.36 | 15.07 | 16.66 | 18.94 | 19.05 | 20.35 |
| **HF_(n.u.)...Frequency_Domain_Results...Normalized_powers...FFT_spectrum** | 36.32 | 41.95 | 35.52 | 41.34 | 36.90 | 41.40 | 13.08 | 15.36 | 17.19 | 19.49 | 19.86 | 21.10 |
| **LF_(%)...Frequency_Domain_Results...Relative_powers...AR_spectrum** | 57.11 | 51.97 | 54.47 | 48.47 | 54.59 | 51.17 | 10.05 | 12.12 | 13.07 | 14.02 | 14.82 | 16.66 |
| **LF_(%)...Frequency_Domain_Results...Relative_powers...FFT_spectrum** | 55.01 | 49.99 | 51.79 | 46.16 | 51.98 | 48.28 | 10.48 | 12.48 | 12.88 | 13.71 | 15.16 | 16.59 |
| **LF_(Hz)...Frequency_Domain_Results...Peak_frequencies...AR_spectrum** | 0.07 | 0.06 | 0.06 | 0.06 | 0.06 | 0.06 | 0.02 | 0.02 | 0.02 | 0.02 | 0.02 | 0.02 |
| **LF_(Hz)...Frequency_Domain_Results...Peak_frequencies...FFT_spectrum** | 0.09 | 0.08 | 0.08 | 0.07 | 0.08 | 0.08 | 0.02 | 0.02 | 0.03 | 0.03 | 0.03 | 0.03 |

| | | | | | | | | | | | | |
|---|---|---|---|---|---|---|---|---|---|---|---|---|
| LF_(log)...Frequency_Domain_Results...Absolute_powers...AR_spectrum | 6.93 | 6.85 | 5.48 | 5.66 | 6.06 | 6.50 | 0.75 | 0.84 | 2.10 | 2.27 | 1.62 | 1.63 |
| LF_(log)...Frequency_Domain_Results...Absolute_powers...FFT_spectrum | 6.91 | 6.81 | 5.43 | 5.61 | 6.01 | 6.45 | 0.75 | 0.82 | 2.08 | 2.24 | 1.59 | 1.59 |
| LF_(ms^2)...Frequency_Domain_Results...Absolute_powers...AR_spectrum | 1360.0 | 1362.6 | 1598.2 | 1704.5 | 1884.2 | 1908.2 | 1217.3 | 1696.2 | 3780.5 | 3869.5 | 5597.6 | 3164.8 |
| LF_(ms^2)...Frequency_Domain_Results...Absolute_powers...FFT_spectrum | 1331.5 | 1288.0 | 1444.9 | 1552.1 | 1630.9 | 1743.4 | 1212.6 | 1552.8 | 3398.0 | 3439.9 | 4320.6 | 2964.1 |
| LF_(n.u.)...Frequency_Domain_Results...Normalized_powers...AR_spectrum | 64.50 | 58.91 | 65.04 | 59.38 | 63.98 | 59.80 | 12.42 | 15.09 | 16.77 | 19.02 | 19.19 | 20.49 |
| LF_(n.u.)...Frequency_Domain_Results...Normalized_powers...FFT_spectrum | 63.57 | 57.96 | 64.30 | 58.50 | 62.91 | 58.40 | 13.14 | 15.39 | 17.29 | 19.57 | 19.99 | 21.24 |
| LF/HF_ratio...Frequency_Domain_Results...AR_spectrum | 2.26 | 1.95 | 2.67 | 2.17 | 2.94 | 2.73 | 1.43 | 1.60 | 2.00 | 1.82 | 2.84 | 3.32 |
| LF/HF_ratio...Frequency_Domain_Results...FFT_spectrum | 2.20 | 1.89 | 2.63 | 2.15 | 2.84 | 2.56 | 1.47 | 1.60 | 2.16 | 1.94 | 2.74 | 2.95 |
| Max_HR_(beats/min)...Time_Domain_Results...Statistical_parameters | 147.4 | 139.3 | 132.4 | 133.0 | 126.2 | 121.2 | 53.0 | 56.8 | 59.7 | 66.2 | 59.0 | 64.0 |
| Max_line_length_(beats)...Nonlinear_Results...Recurrence_plot_analysis | 255.8 | 213.7 | 272.6 | 271.4 | 274.5 | 241.7 | 122.8 | 107.8 | 153.1 | 173.6 | 168.1 | 165.8 |
| Mean_HR_(beats/min)...Time_Domain_Results...Statistical_parameters | 79.3 | 69.9 | 87.1 | 83.5 | 80.2 | 77.1 | 18.8 | 23.7 | 23.3 | 27.3 | 22.3 | 29.0 |
| Mean_line_length_(beats)...Nonlinear_Results...Recurrence_plot_analysis | 12.7 | 13.5 | 15.5 | 15.5 | 14.3 | 14.3 | 6.0 | 5.4 | 12.4 | 13.9 | 12.1 | 11.9 |
| Mean_RR__(ms)...Time_Domain_Results...Statistical_parameters | 800.3 | 915.2 | 732.6 | 775.4 | 790.6 | 852.6 | 202.4 | 191.9 | 187.6 | 201.3 | 170.7 | 216.6 |
| Min_HR_(beats/min)...Time_Domain_Results...Statistical_parameters | 56.7 | 50.8 | 63.4 | 60.5 | 57.8 | 52.2 | 11.2 | 9.3 | 17.0 | 16.9 | 13.2 | 12.0 |
| MSE(1)...Nonlinear_Results...Multiscale_entropy | 1.39 | 1.33 | 1.51 | 1.48 | 1.45 | 1.40 | 0.41 | 0.46 | 0.42 | 0.44 | 0.36 | 0.46 |
| MSE(10)...Nonlinear_Results...Multiscale_entropy | 0.94 | 0.88 | 0.98 | 0.95 | 0.99 | 0.93 | 0.26 | 0.27 | 0.39 | 0.42 | 0.27 | 0.31 |
| MSE(11)...Nonlinear_Results...Multiscale_entropy | 0.88 | 0.82 | 0.93 | 0.91 | 0.95 | 0.88 | 0.25 | 0.26 | 0.38 | 0.40 | 0.29 | 0.31 |
| MSE(12)...Nonlinear_Results...Multiscale_entropy | 0.81 | 0.76 | 0.87 | 0.85 | 0.89 | 0.82 | 0.23 | 0.24 | 0.38 | 0.39 | 0.29 | 0.31 |
| MSE(13)...Nonlinear_Results...Multiscale_entropy | 0.76 | 0.71 | 0.82 | 0.80 | 0.85 | 0.78 | 0.20 | 0.19 | 0.37 | 0.38 | 0.28 | 0.30 |
| MSE(14)...Nonlinear_Results...Multiscale_entropy | 0.72 | 0.68 | 0.77 | 0.75 | 0.79 | 0.74 | 0.19 | 0.20 | 0.34 | 0.35 | 0.23 | 0.26 |
| MSE(15)...Nonlinear_Results...Multiscale_entropy | 0.68 | 0.65 | 0.73 | 0.71 | 0.73 | 0.68 | 0.16 | 0.17 | 0.31 | 0.32 | 0.22 | 0.25 |
| MSE(16)...Nonlinear_Results...Multiscale_entropy | 0.64 | 0.61 | 0.70 | 0.68 | 0.69 | 0.64 | 0.15 | 0.16 | 0.32 | 0.33 | 0.21 | 0.23 |
| MSE(17)...Nonlinear_Results...Multiscale_entropy | 0.61 | 0.57 | 0.66 | 0.64 | 0.66 | 0.61 | 0.16 | 0.16 | 0.31 | 0.31 | 0.19 | 0.21 |
| MSE(18)...Nonlinear_Results...Multiscale_entropy | 0.57 | 0.55 | 0.61 | 0.60 | 0.61 | 0.57 | 0.15 | 0.16 | 0.28 | 0.29 | 0.18 | 0.20 |
| MSE(19)...Nonlinear_Results...Multiscale_entropy | 0.55 | 0.54 | 0.60 | 0.58 | 0.57 | 0.54 | 0.13 | 0.14 | 0.27 | 0.27 | 0.18 | 0.20 |
| MSE(2)...Nonlinear_Results...Multiscale_entropy | 1.46 | 1.39 | 1.42 | 1.38 | 1.45 | 1.38 | 0.42 | 0.47 | 0.41 | 0.46 | 0.31 | 0.42 |

| | | | | | | | | | | | | |
|---|---|---|---|---|---|---|---|---|---|---|---|---|
| **MSE(20)...Nonlinear_Results...Multiscale_entropy** | 0.52 | 0.50 | 0.56 | 0.55 | 0.57 | 0.53 | 0.13 | 0.12 | 0.26 | 0.26 | 0.19 | 0.20 |
| **MSE(3)...Nonlinear_Results...Multiscale_entropy** | 1.38 | 1.30 | 1.34 | 1.31 | 1.39 | 1.32 | 0.40 | 0.45 | 0.36 | 0.40 | 0.30 | 0.38 |
| **MSE(4)...Nonlinear_Results...Multiscale_entropy** | 1.36 | 1.29 | 1.28 | 1.24 | 1.37 | 1.29 | 0.39 | 0.43 | 0.38 | 0.42 | 0.32 | 0.39 |
| **MSE(5)...Nonlinear_Results...Multiscale_entropy** | 1.32 | 1.25 | 1.27 | 1.25 | 1.33 | 1.25 | 0.37 | 0.40 | 0.37 | 0.41 | 0.32 | 0.38 |
| **MSE(6)...Nonlinear_Results...Multiscale_entropy** | 1.24 | 1.18 | 1.21 | 1.18 | 1.27 | 1.19 | 0.35 | 0.38 | 0.43 | 0.46 | 0.34 | 0.39 |
| **MSE(7)...Nonlinear_Results...Multiscale_entropy** | 1.18 | 1.12 | 1.16 | 1.13 | 1.23 | 1.15 | 0.32 | 0.36 | 0.41 | 0.45 | 0.33 | 0.37 |
| **MSE(8)...Nonlinear_Results...Multiscale_entropy** | 1.08 | 1.02 | 1.11 | 1.08 | 1.15 | 1.06 | 0.31 | 0.32 | 0.42 | 0.46 | 0.32 | 0.36 |
| **MSE(9)...Nonlinear_Results...Multiscale_entropy** | 1.00 | 0.93 | 1.04 | 1.01 | 1.08 | 1.01 | 0.27 | 0.28 | 0.40 | 0.42 | 0.33 | 0.36 |
| **NNxx_(beats)...Time_Domain_Results...Statistical_parameters** | 905.0 | 999.6 | 752.2 | 907.7 | 892.1 | 1217.0 | 928.8 | 1070.9 | 1440.6 | 1562.0 | 1506.7 | 1764.0 |
| **pNNxx_(%)...Time_Domain_Results...Statistical_parameters** | 19.25 | 24.01 | 13.67 | 16.89 | 15.91 | 21.07 | 16.90 | 20.39 | 24.28 | 25.58 | 22.56 | 24.42 |
| **Recurrence_rate_(REC)_(%)...Nonlinear_Results...Recurrence_plot_analysis** | 33.14 | 33.74 | 33.71 | 32.81 | 32.77 | 32.02 | 8.27 | 7.33 | 9.68 | 10.31 | 10.05 | 10.58 |
| **RMSSD_(ms)...Time_Domain_Results...Statistical_parameters** | 47.80 | 53.70 | 45.33 | 50.22 | 49.25 | 59.90 | 32.54 | 32.77 | 68.85 | 64.97 | 56.03 | 60.62 |
| **RR_tri_index...Time_Domain_Results...Geometric_parameters** | 11.86 | 11.87 | 8.71 | 9.87 | 9.06 | 11.62 | 5.01 | 4.66 | 11.47 | 10.82 | 7.26 | 8.28 |
| **Sample_entropy_(SampEn)...Nonlinear_Results** | 1.52 | 1.59 | 1.57 | 1.60 | 1.53 | 1.57 | 0.22 | 0.18 | 0.31 | 0.29 | 0.28 | 0.28 |
| **SD1_(ms)...Nonlinear_Results...Poincare_plot** | 33.80 | 37.98 | 32.05 | 35.52 | 34.83 | 42.36 | 23.01 | 23.17 | 48.69 | 45.95 | 39.62 | 42.87 |
| **SD2_(ms)...Nonlinear_Results...Poincare_plot** | 62.11 | 61.36 | 44.57 | 50.84 | 51.64 | 62.23 | 26.24 | 26.58 | 48.96 | 48.55 | 44.66 | 42.68 |
| **SD2/SD1_ratio...Nonlinear_Results...Poincare_plot** | 2.11 | 1.86 | 2.06 | 2.03 | 2.03 | 1.88 | 0.55 | 0.58 | 0.78 | 0.80 | 0.81 | 0.76 |
| **SDANN_(ms)...Time_Domain_Results...Statistical_parameters** | 73.20 | 69.87 | 62.70 | 55.58 | 65.73 | 63.57 | 58.61 | 73.60 | 63.79 | 81.93 | 70.62 | 81.48 |
| **SDNN_index_(ms)...Time_Domain_Results...Statistical_parameters** | 76.14 | 76.12 | 56.39 | 57.51 | 65.06 | 72.00 | 27.48 | 28.69 | 49.73 | 47.23 | 46.63 | 44.37 |
| **Shannon_entropy...Nonlinear_Results...Recurrence_plot_analysis** | 3.20 | 3.28 | 3.21 | 3.20 | 3.20 | 3.20 | 0.28 | 0.23 | 0.37 | 0.37 | 0.37 | 0.38 |
| **STD_HR_(beats/min)...Time_Domain_Results...Statistical_parameters** | 20.90 | 61.47 | 7.66 | 9.14 | 8.70 | 10.50 | 77.61 | 534.99 | 11.66 | 13.95 | 12.50 | 14.39 |
| **STD_RR_(ms)...Time_Domain_Results...Statistical_parameters** | 50.35 | 51.42 | 39.45 | 44.52 | 44.91 | 54.30 | 23.97 | 24.11 | 48.32 | 46.75 | 41.30 | 41.40 |
| **TINN_(ms)...Time_Domain_Results...Geometric_parameters** | 430.14 | 457.72 | 359.45 | 438.55 | 409.26 | 455.07 | 276.63 | 286.26 | 413.21 | 569.13 | 505.02 | 437.26 |
| **Total_power_(ms^2)...Frequency_Domain_Results...AR_spectrum** | 2568.78 | 2831.36 | 3732.91 | 4012.48 | 3875.99 | 3923.71 | 2615.94 | 3801.41 | 8654.61 | 8404.53 | 10426.46 | 6242.91 |
| **Total_power_(ms^2)...Frequency_Domain_Results...FFT_spectrum** | 2656.92 | 2847.56 | 3684.63 | 4083.90 | 3644.74 | 3896.62 | 2788.33 | 3811.94 | 8508.11 | 8718.96 | 8979.99 | 6242.36 |

| | | | | | | | | | | | |
|---|---|---|---|---|---|---|---|---|---|---|---|
| **VLF_(%)...Frequency_Domain_Results...Relative_powers...AR_spectrum** | 10.89 | 11.12 | 15.10 | 16.26 | 13.23 | 13.03 | 6.30 | 5.01 | 8.24 | 10.50 | 7.46 | 8.10 |
| VLF_(%)...Frequency_Domain_Results...Relative_powers...FFT_spectrum | **13.00** | **13.25** | **18.28** | **18.86** | **16.01** | **15.85** | **4.91** | **4.56** | **7.37** | **9.70** | **6.49** | **7.10** |
| **VLF_(Hz)...Frequency_Domain_Results...Peak_frequencies...AR_spectrum** | 0.04 | 0.04 | 0.04 | 0.04 | 0.04 | 0.04 | 0.00 | 0.00 | 0.00 | 0.00 | 0.00 | 0.00 |
| **VLF_(Hz)...Frequency_Domain_Results...Peak_frequencies...FFT_spectrum** | 0.04 | 0.04 | 0.04 | 0.04 | 0.04 | 0.04 | 0.00 | 0.01 | 0.01 | 0.01 | 0.01 | 0.01 |
| **VLF_(log)...Frequency_Domain_Results...Absolute_powers...AR_spectrum** | 5.18 | 5.24 | 4.01 | 4.31 | 4.48 | 4.97 | 0.78 | 0.81 | 2.02 | 2.35 | 1.52 | 1.42 |
| **VLF_(log)...Frequency_Domain_Results...Absolute_powers...FFT_spectrum** | 5.43 | 5.46 | 4.31 | 4.59 | 4.79 | 5.29 | 0.80 | 0.88 | 2.02 | 2.28 | 1.55 | 1.46 |
| **VLF_(ms^2)...Frequency_Domain_Results...Absolute_powers...AR_spectrum** | 240.89 | 282.78 | 308.30 | 403.68 | 607.67 | 435.03 | 240.42 | 437.50 | 1129.47 | 1354.03 | 3337.89 | 1447.12 |
| **VLF_(ms^2)...Frequency_Domain_Results...Absolute_powers...FFT_spectrum** | 318.59 | 375.35 | 419.24 | 641.08 | 671.64 | 591.08 | 334.96 | 604.08 | 1164.12 | 2472.64 | 2877.00 | 1682.85 |

#### Table S 8. Two-Way ANOVA BioPatch & Kubios

| Measurement | Data_Source | Term | ANOVA.p_value | ANOVA.q_value |
|---|---|---|---|---|
| **Activity** | Zephyr | Time_of_day | 4.13766E-20 | 1.26612E-17 |
| **Activity** | Zephyr | Cohort | 4.08527E-06 | 9.61611E-05 |
| **Activity** | Zephyr | Cohort:Time_of_day | 0.000124081 | 0.001808041 |
| **BR** | Zephyr | Time_of_day | 5.17426E-06 | 0.000113095 |
| **BR** | Zephyr | Cohort | 0.705114773 | 0.910114258 |
| **BR** | Zephyr | Cohort:Time_of_day | 0.017537612 | 0.107275792 |
| **BRAmplitude** | Zephyr | Time_of_day | 0.000628855 | 0.006635504 |
| **BRAmplitude** | Zephyr | Cohort | 0.581077645 | 0.827022136 |
| **BRAmplitude** | Zephyr | Cohort:Time_of_day | 0.199296411 | 0.413918475 |
| **BRConfidence** | Zephyr | Time_of_day | 8.14697E-08 | 3.11621E-06 |
| **BRConfidence** | Zephyr | Cohort | 0.391770689 | 0.647379849 |
| **BRConfidence** | Zephyr | Cohort:Time_of_day | 0.538760945 | 0.788807891 |
| **BreathingWaveform** | Zephyr | Time_of_day | 0.0150805 | 0.094883828 |
| **BreathingWaveform** | Zephyr | Cohort | 0.001112878 | 0.01031941 |
| **BreathingWaveform** | Zephyr | Cohort:Time_of_day | 0.455462994 | 0.707255193 |
| **CoreTemp** | Zephyr | Time_of_day | 1.02094E-06 | 3.12408E-05 |
| **CoreTemp** | Zephyr | Cohort | 0.493702139 | 0.744201255 |
| **CoreTemp** | Zephyr | Cohort:Time_of_day | 0.143177796 | 0.334445843 |
| **EKGAmplitude** | Zephyr | Time_of_day | 2.21831E-06 | 5.65669E-05 |
| **EKGAmplitude** | Zephyr | Cohort | 0.441671877 | 0.701529546 |
| **EKGAmplitude** | Zephyr | Cohort:Time_of_day | 0.816301619 | 0.910114258 |
| **EKGNoise** | Zephyr | Time_of_day | 1.16347E-07 | 3.95581E-06 |
| **EKGNoise** | Zephyr | Cohort | 0.034573878 | 0.165306353 |
| **EKGNoise** | Zephyr | Cohort:Time_of_day | 0.005286663 | 0.040442969 |
| **HR** | Zephyr | Time_of_day | 1.44747E-10 | 6.32749E-09 |
| **HR** | Zephyr | Cohort | 0.1616742 | 0.360377253 |
| **HR** | Zephyr | Cohort:Time_of_day | 0.184321948 | 0.39442319 |
| **HRConfidence** | Zephyr | Time_of_day | 0.001082055 | 0.01031941 |
| **HRConfidence** | Zephyr | Cohort | 0.150364919 | 0.343370636 |
| **HRConfidence** | Zephyr | Cohort:Time_of_day | 0.194533698 | 0.410533184 |
| **HRV** | Zephyr | Time_of_day | 0.092410831 | 0.248561834 |
| **HRV** | Zephyr | Cohort | 0.01129857 | 0.078576422 |
| **HRV** | Zephyr | Cohort:Time_of_day | 0.278502322 | 0.516495216 |
| **Lateral** | Zephyr | Time_of_day | 0.209899309 | 0.425358865 |
| **Lateral** | Zephyr | Cohort | 0.60032351 | 0.838808193 |
| **Lateral** | Zephyr | Cohort:Time_of_day | 0.848790672 | 0.910133522 |
| **LateralMin** | Zephyr | Time_of_day | 0.021928251 | 0.12426009 |
| **LateralMin** | Zephyr | Cohort | 0.698210106 | 0.910114258 |
| **LateralMin** | Zephyr | Cohort:Time_of_day | 0.794117759 | 0.910114258 |

| | | | | |
|---|---|---|---|---|
| **LateralPeak** | Zephyr | Time_of_day | 0.814567333 | 0.910114258 |
| **LateralPeak** | Zephyr | Cohort | 0.465122709 | 0.715213813 |
| **LateralPeak** | Zephyr | Cohort:Time_of_day | 0.853250795 | 0.910133522 |
| **PeakAccel** | Zephyr | Time_of_day | 1.90115E-19 | 2.90877E-17 |
| **PeakAccel** | Zephyr | Cohort | 1.68824E-05 | 0.000322876 |
| **PeakAccel** | Zephyr | Cohort:Time_of_day | 0.00015606 | 0.002170649 |
| **Posture** | Zephyr | Time_of_day | 0.041129724 | 0.17979565 |
| **Posture** | Zephyr | Cohort | 0.816324196 | 0.910114258 |
| **Posture** | Zephyr | Cohort:Time_of_day | 0.41199882 | 0.667045709 |
| **Sagittal** | Zephyr | Time_of_day | 0.217733647 | 0.432639583 |
| **Sagittal** | Zephyr | Cohort | 0.259110911 | 0.486429072 |
| **Sagittal** | Zephyr | Cohort:Time_of_day | 0.163700778 | 0.360377253 |
| **SagittalMin** | Zephyr | Time_of_day | 0.055258677 | 0.204637691 |
| **SagittalMin** | Zephyr | Cohort | 0.200195864 | 0.413918475 |
| **SagittalMin** | Zephyr | Cohort:Time_of_day | 0.115108008 | 0.284056859 |
| **SagittalPeak** | Zephyr | Time_of_day | 0.554092294 | 0.799774726 |
| **SagittalPeak** | Zephyr | Cohort | 0.324821553 | 0.562845771 |
| **SagittalPeak** | Zephyr | Cohort:Time_of_day | 0.205551586 | 0.420441695 |
| **VectorMagnitude** | Zephyr | Time_of_day | 2.61566E-11 | 1.33399E-09 |
| **VectorMagnitude** | Zephyr | Cohort | 0.78095116 | 0.910114258 |
| **VectorMagnitude** | Zephyr | Cohort:Time_of_day | 0.061810396 | 0.204637691 |
| **Vertical** | Zephyr | Time_of_day | 7.29673E-14 | 5.582E-12 |
| **Vertical** | Zephyr | Cohort | 0.101082397 | 0.259926164 |
| **Vertical** | Zephyr | Cohort:Time_of_day | 0.301410139 | 0.545748536 |
| **VerticalMin** | Zephyr | Time_of_day | 1.25386E-14 | 1.27893E-12 |
| **VerticalMin** | Zephyr | Cohort | 0.122832262 | 0.295958048 |
| **VerticalMin** | Zephyr | Cohort:Time_of_day | 0.321617208 | 0.562845771 |
| **VerticalPeak** | Zephyr | Time_of_day | 4.70513E-13 | 2.87954E-11 |
| **VerticalPeak** | Zephyr | Cohort | 0.07759584 | 0.219854879 |
| **VerticalPeak** | Zephyr | Cohort:Time_of_day | 0.255687525 | 0.482965326 |
| **alpha_1...Nonlinear_Results...Detrended_fluctuation_analysis** | Kubios | Time_of_day | 0.003083316 | 0.024192171 |
| **alpha_1...Nonlinear_Results...Detrended_fluctuation_analysis** | Kubios | Cohort | 0.809025659 | 0.910114258 |
| **alpha_1...Nonlinear_Results...Detrended_fluctuation_analysis** | Kubios | Cohort:Time_of_day | 0.532078839 | 0.786551327 |
| **alpha_2...Nonlinear_Results...Detrended_fluctuation_analysis** | Kubios | Time_of_day | 0.001152097 | 0.01036887 |
| **alpha_2...Nonlinear_Results...Detrended_fluctuation_analysis** | Kubios | Cohort | 0.002591817 | 0.02087095 |
| **alpha_2...Nonlinear_Results...Detrended_fluctuation_analysis** | Kubios | Cohort:Time_of_day | 0.404287362 | 0.661561138 |
| **Approximate_entropy_(ApEn)...Nonlinear_Results** | Kubios | Time_of_day | 0.060334935 | 0.204637691 |
| **Approximate_entropy_(ApEn)...Nonlinear_Results** | Kubios | Cohort | 0.753463652 | 0.910114258 |
| **Approximate_entropy_(ApEn)...Nonlinear_Results** | Kubios | Cohort:Time_of_day | 0.802754915 | 0.910114258 |
| **Correlation_dimension_(D2)...Nonlinear_Results** | Kubios | Time_of_day | 0.052274651 | 0.204637691 |
| **Correlation_dimension_(D2)...Nonlinear_Results** | Kubios | Cohort | 6.65448E-05 | 0.001082735 |
| **Correlation_dimension_(D2)...Nonlinear_Results** | Kubios | Cohort:Time_of_day | 0.393505398 | 0.647379849 |
| **Determinism_(DET)_(%)...Nonlinear_Results...Recurrence_plot_analysis** | Kubios | Time_of_day | 0.090397782 | 0.246979654 |

| | | | | |
|---|---|---|---|---|
| **Determinism_(DET)_(%)...Nonlinear_Results...Recurrence_plot_analysis** | Kubios | Cohort | 0.758865743 | 0.910114258 |
| **Determinism_(DET)_(%)...Nonlinear_Results...Recurrence_plot_analysis** | Kubios | Cohort:Time_of_day | 0.736307228 | 0.910114258 |
| **EDR_(Hz)...Frequency_Domain_Results...FFT_spectrum** | Kubios | Time_of_day | 0.537098289 | 0.788807891 |
| **EDR_(Hz)...Frequency_Domain_Results...FFT_spectrum** | Kubios | Cohort | 1.78989E-06 | 4.97915E-05 |
| **EDR_(Hz)...Frequency_Domain_Results...FFT_spectrum** | Kubios | Cohort:Time_of_day | 0.240402174 | 0.465589021 |
| **HF_(%)...Frequency_Domain_Results...Relative_powers...AR_spectrum** | Kubios | Time_of_day | 0.001648346 | 0.013632267 |
| **HF_(%)...Frequency_Domain_Results...Relative_powers...AR_spectrum** | Kubios | Cohort | 0.805455251 | 0.910114258 |
| **HF_(%)...Frequency_Domain_Results...Relative_powers...AR_spectrum** | Kubios | Cohort:Time_of_day | 0.766002229 | 0.910114258 |
| **HF_(%)...Frequency_Domain_Results...Relative_powers...FFT_spectrum** | Kubios | Time_of_day | 0.000913595 | 0.00931867 |
| **HF_(%)...Frequency_Domain_Results...Relative_powers...FFT_spectrum** | Kubios | Cohort | 0.761484966 | 0.910114258 |
| **HF_(%)...Frequency_Domain_Results...Relative_powers...FFT_spectrum** | Kubios | Cohort:Time_of_day | 0.771206545 | 0.910114258 |
| **HF_(Hz)...Frequency_Domain_Results...Peak_frequencies...AR_spectrum** | Kubios | Time_of_day | 0.071990473 | 0.218109751 |
| **HF_(Hz)...Frequency_Domain_Results...Peak_frequencies...AR_spectrum** | Kubios | Cohort | 0.623547097 | 0.857663783 |
| **HF_(Hz)...Frequency_Domain_Results...Peak_frequencies...AR_spectrum** | Kubios | Cohort:Time_of_day | 0.547428019 | 0.797680827 |
| **HF_(Hz)...Frequency_Domain_Results...Peak_frequencies...FFT_spectrum** | Kubios | Time_of_day | 0.001384212 | 0.012101969 |
| **HF_(Hz)...Frequency_Domain_Results...Peak_frequencies...FFT_spectrum** | Kubios | Cohort | 0.979383118 | 0.992200057 |
| **HF_(Hz)...Frequency_Domain_Results...Peak_frequencies...FFT_spectrum** | Kubios | Cohort:Time_of_day | 0.522755641 | 0.776520515 |
| **HF_(log)...Frequency_Domain_Results...Absolute_powers...AR_spectrum** | Kubios | Time_of_day | 0.00048898 | 0.005602714 |
| **HF_(log)...Frequency_Domain_Results...Absolute_powers...AR_spectrum** | Kubios | Cohort | 0.042494692 | 0.183146135 |
| **HF_(log)...Frequency_Domain_Results...Absolute_powers...AR_spectrum** | Kubios | Cohort:Time_of_day | 0.262803543 | 0.490352951 |
| **HF_(log)...Frequency_Domain_Results...Absolute_powers...FFT_spectrum** | Kubios | Time_of_day | 0.000382855 | 0.004881404 |
| **HF_(log)...Frequency_Domain_Results...Absolute_powers...FFT_spectrum** | Kubios | Cohort | 0.038869281 | 0.174911763 |
| **HF_(log)...Frequency_Domain_Results...Absolute_powers...FFT_spectrum** | Kubios | Cohort:Time_of_day | 0.236527448 | 0.461002542 |
| **HF_(ms^2)...Frequency_Domain_Results...Absolute_powers...AR_spectrum** | Kubios | Time_of_day | 0.086167691 | 0.239083916 |
| **HF_(ms^2)...Frequency_Domain_Results...Absolute_powers...AR_spectrum** | Kubios | Cohort | 0.84053362 | 0.910133522 |
| **HF_(ms^2)...Frequency_Domain_Results...Absolute_powers...AR_spectrum** | Kubios | Cohort:Time_of_day | 0.852027277 | 0.910133522 |
| **HF_(ms^2)...Frequency_Domain_Results...Absolute_powers...FFT_spectrum** | Kubios | Time_of_day | 0.079934768 | 0.224404028 |
| **HF_(ms^2)...Frequency_Domain_Results...Absolute_powers...FFT_spectrum** | Kubios | Cohort | 0.855278154 | 0.910133522 |
| **HF_(ms^2)...Frequency_Domain_Results...Absolute_powers...FFT_spectrum** | Kubios | Cohort:Time_of_day | 0.863841226 | 0.911501431 |
| **HF_(n.u.)...Frequency_Domain_Results...Normalized_powers...AR_spectrum** | Kubios | Time_of_day | 0.000494357 | 0.005602714 |
| **HF_(n.u.)...Frequency_Domain_Results...Normalized_powers...AR_spectrum** | Kubios | Cohort | 0.937998479 | 0.965255479 |
| **HF_(n.u.)...Frequency_Domain_Results...Normalized_powers...AR_spectrum** | Kubios | Cohort:Time_of_day | 0.758755184 | 0.910114258 |
| **HF_(n.u.)...Frequency_Domain_Results...Normalized_powers...FFT_spectrum** | Kubios | Time_of_day | 0.000376207 | 0.004881404 |
| **HF_(n.u.)...Frequency_Domain_Results...Normalized_powers...FFT_spectrum** | Kubios | Cohort | 0.909206804 | 0.945168969 |
| **HF_(n.u.)...Frequency_Domain_Results...Normalized_powers...FFT_spectrum** | Kubios | Cohort:Time_of_day | 0.799072996 | 0.910114258 |
| **LF/HF_ratio...Frequency_Domain_Results...AR_spectrum** | Kubios | Time_of_day | 0.062349828 | 0.204637691 |
| **LF/HF_ratio...Frequency_Domain_Results...AR_spectrum** | Kubios | Cohort | 0.375255786 | 0.627476888 |
| **LF/HF_ratio...Frequency_Domain_Results...AR_spectrum** | Kubios | Cohort:Time_of_day | 0.829290136 | 0.910114258 |
| **LF/HF_ratio...Frequency_Domain_Results...FFT_spectrum** | Kubios | Time_of_day | 0.039444798 | 0.174929104 |
| **LF/HF_ratio...Frequency_Domain_Results...FFT_spectrum** | Kubios | Cohort | 0.433072835 | 0.693823495 |
| **LF/HF_ratio...Frequency_Domain_Results...FFT_spectrum** | Kubios | Cohort:Time_of_day | 0.856596256 | 0.910133522 |
| **LF_(%)...Frequency_Domain_Results...Relative_powers...AR_spectrum** | Kubios | Time_of_day | 6.72287E-05 | 0.001082735 |
| **LF_(%)...Frequency_Domain_Results...Relative_powers...AR_spectrum** | Kubios | Cohort | 0.768485564 | 0.910114258 |

| | | | | |
|---|---|---|---|---|
| **LF_(%)...Frequency_Domain_Results...Relative_powers...AR_spectrum** | Kubios | Cohort:Time_of_day | 0.558266868 | 0.80201719 |
| **LF_(%)...Frequency_Domain_Results...Relative_powers...FFT_spectrum** | Kubios | Time_of_day | 5.68826E-05 | 0.001023888 |
| **LF_(%)...Frequency_Domain_Results...Relative_powers...FFT_spectrum** | Kubios | Cohort | 0.708395771 | 0.910114258 |
| **LF_(%)...Frequency_Domain_Results...Relative_powers...FFT_spectrum** | Kubios | Cohort:Time_of_day | 0.6591689 | 0.892503024 |
| **LF_(Hz)...Frequency_Domain_Results...Peak_frequencies...AR_spectrum** | Kubios | Time_of_day | 0.030611609 | 0.153559876 |
| **LF_(Hz)...Frequency_Domain_Results...Peak_frequencies...AR_spectrum** | Kubios | Cohort | 0.179978475 | 0.38784094 |
| **LF_(Hz)...Frequency_Domain_Results...Peak_frequencies...AR_spectrum** | Kubios | Cohort:Time_of_day | 0.109028945 | 0.27124274 |
| **LF_(Hz)...Frequency_Domain_Results...Peak_frequencies...FFT_spectrum** | Kubios | Time_of_day | 0.015193816 | 0.094883828 |
| **LF_(Hz)...Frequency_Domain_Results...Peak_frequencies...FFT_spectrum** | Kubios | Cohort | 0.316920535 | 0.560564645 |
| **LF_(Hz)...Frequency_Domain_Results...Peak_frequencies...FFT_spectrum** | Kubios | Cohort:Time_of_day | 0.107692905 | 0.270114991 |
| **LF_(log)...Frequency_Domain_Results...Absolute_powers...AR_spectrum** | Kubios | Time_of_day | 0.013002264 | 0.084653037 |
| **LF_(log)...Frequency_Domain_Results...Absolute_powers...AR_spectrum** | Kubios | Cohort | 0.025221546 | 0.135399878 |
| **LF_(log)...Frequency_Domain_Results...Absolute_powers...AR_spectrum** | Kubios | Cohort:Time_of_day | 0.154076932 | 0.349241047 |
| **LF_(log)...Frequency_Domain_Results...Absolute_powers...FFT_spectrum** | Kubios | Time_of_day | 0.012648693 | 0.084653037 |
| **LF_(log)...Frequency_Domain_Results...Absolute_powers...FFT_spectrum** | Kubios | Cohort | 0.02266194 | 0.126082796 |
| **LF_(log)...Frequency_Domain_Results...Absolute_powers...FFT_spectrum** | Kubios | Cohort:Time_of_day | 0.136488079 | 0.326291814 |
| **LF_(ms^2)...Frequency_Domain_Results...Absolute_powers...AR_spectrum** | Kubios | Time_of_day | 0.457635713 | 0.707255193 |
| **LF_(ms^2)...Frequency_Domain_Results...Absolute_powers...AR_spectrum** | Kubios | Cohort | 0.688951155 | 0.910114258 |
| **LF_(ms^2)...Frequency_Domain_Results...Absolute_powers...AR_spectrum** | Kubios | Cohort:Time_of_day | 0.704434562 | 0.910114258 |
| **LF_(ms^2)...Frequency_Domain_Results...Absolute_powers...FFT_spectrum** | Kubios | Time_of_day | 0.373256379 | 0.627476888 |
| **LF_(ms^2)...Frequency_Domain_Results...Absolute_powers...FFT_spectrum** | Kubios | Cohort | 0.783355307 | 0.910114258 |
| **LF_(ms^2)...Frequency_Domain_Results...Absolute_powers...FFT_spectrum** | Kubios | Cohort:Time_of_day | 0.676353311 | 0.907737339 |
| **LF_(n.u.)...Frequency_Domain_Results...Normalized_powers...AR_spectrum** | Kubios | Time_of_day | 0.000540635 | 0.005908366 |
| **LF_(n.u.)...Frequency_Domain_Results...Normalized_powers...AR_spectrum** | Kubios | Cohort | 0.940020042 | 0.965255479 |
| **LF_(n.u.)...Frequency_Domain_Results...Normalized_powers...AR_spectrum** | Kubios | Cohort:Time_of_day | 0.764877053 | 0.910114258 |
| **LF_(n.u.)...Frequency_Domain_Results...Normalized_powers...FFT_spectrum** | Kubios | Time_of_day | 0.000407449 | 0.004987178 |
| **LF_(n.u.)...Frequency_Domain_Results...Normalized_powers...FFT_spectrum** | Kubios | Cohort | 0.910631849 | 0.945168969 |
| **LF_(n.u.)...Frequency_Domain_Results...Normalized_powers...FFT_spectrum** | Kubios | Cohort:Time_of_day | 0.803408786 | 0.910114258 |
| **Max_HR_(beats/min)...Time_Domain_Results...Statistical_parameters** | Kubios | Time_of_day | 0.355486481 | 0.607703147 |
| **Max_HR_(beats/min)...Time_Domain_Results...Statistical_parameters** | Kubios | Cohort | 0.066077621 | 0.206324001 |
| **Max_HR_(beats/min)...Time_Domain_Results...Statistical_parameters** | Kubios | Cohort:Time_of_day | 0.587497228 | 0.83228774 |
| **Max_line_length_(beats)...Nonlinear_Results...Recurrence_plot_analysis** | Kubios | Time_of_day | 0.062862559 | 0.204637691 |
| **Max_line_length_(beats)...Nonlinear_Results...Recurrence_plot_analysis** | Kubios | Cohort | 0.57491243 | 0.822071044 |
| **Max_line_length_(beats)...Nonlinear_Results...Recurrence_plot_analysis** | Kubios | Cohort:Time_of_day | 0.442467982 | 0.701529546 |
| **Mean_HR_(beats/min)...Time_Domain_Results...Statistical_parameters** | Kubios | Time_of_day | 0.025137375 | 0.135399878 |
| **Mean_HR_(beats/min)...Time_Domain_Results...Statistical_parameters** | Kubios | Cohort | 0.028041141 | 0.143009819 |
| **Mean_HR_(beats/min)...Time_Domain_Results...Statistical_parameters** | Kubios | Cohort:Time_of_day | 0.550906306 | 0.79894469 |
| **Mean_line_length_(beats)...Nonlinear_Results...Recurrence_plot_analysis** | Kubios | Time_of_day | 0.835758518 | 0.910114258 |
| **Mean_line_length_(beats)...Nonlinear_Results...Recurrence_plot_analysis** | Kubios | Cohort | 0.163598602 | 0.360377253 |
| **Mean_line_length_(beats)...Nonlinear_Results...Recurrence_plot_analysis** | Kubios | Cohort:Time_of_day | 0.835392348 | 0.910114258 |
| **Mean_RR__(ms)...Time_Domain_Results...Statistical_parameters** | Kubios | Time_of_day | 8.39555E-05 | 0.00128452 |
| **Mean_RR__(ms)...Time_Domain_Results...Statistical_parameters** | Kubios | Cohort | 0.034259044 | 0.165306353 |
| **Mean_RR__(ms)...Time_Domain_Results...Statistical_parameters** | Kubios | Cohort:Time_of_day | 0.246830139 | 0.475031589 |

| | | | | |
|---|---|---|---|---|
| **Min_HR_(beats/min)...Time_Domain_Results...Statistical_parameters** | Kubios | Time_of_day | 1.3153E-05 | 0.000268321 |
| **Min_HR_(beats/min)...Time_Domain_Results...Statistical_parameters** | Kubios | Cohort | 0.02742446 | 0.142235333 |
| **Min_HR_(beats/min)...Time_Domain_Results...Statistical_parameters** | Kubios | Cohort:Time_of_day | 0.721849302 | 0.910114258 |
| **MSE(1)...Nonlinear_Results...Multiscale_entropy** | Kubios | Time_of_day | 0.47418307 | 0.725500097 |
| **MSE(1)...Nonlinear_Results...Multiscale_entropy** | Kubios | Cohort | 0.48951147 | 0.741537177 |
| **MSE(1)...Nonlinear_Results...Multiscale_entropy** | Kubios | Cohort:Time_of_day | 0.36911766 | 0.627476888 |
| **MSE(10)...Nonlinear_Results...Multiscale_entropy** | Kubios | Time_of_day | 0.061235277 | 0.204637691 |
| **MSE(10)...Nonlinear_Results...Multiscale_entropy** | Kubios | Cohort | 0.83215701 | 0.910114258 |
| **MSE(10)...Nonlinear_Results...Multiscale_entropy** | Kubios | Cohort:Time_of_day | 0.074480067 | 0.219854879 |
| **MSE(11)...Nonlinear_Results...Multiscale_entropy** | Kubios | Time_of_day | 0.038009349 | 0.174852571 |
| **MSE(11)...Nonlinear_Results...Multiscale_entropy** | Kubios | Cohort | 0.741097123 | 0.910114258 |
| **MSE(11)...Nonlinear_Results...Multiscale_entropy** | Kubios | Cohort:Time_of_day | 0.059554637 | 0.204637691 |
| **MSE(12)...Nonlinear_Results...Multiscale_entropy** | Kubios | Time_of_day | 0.057276324 | 0.204637691 |
| **MSE(12)...Nonlinear_Results...Multiscale_entropy** | Kubios | Cohort | 0.716976542 | 0.910114258 |
| **MSE(12)...Nonlinear_Results...Multiscale_entropy** | Kubios | Cohort:Time_of_day | 0.061754349 | 0.204637691 |
| **MSE(13)...Nonlinear_Results...Multiscale_entropy** | Kubios | Time_of_day | 0.054633399 | 0.204637691 |
| **MSE(13)...Nonlinear_Results...Multiscale_entropy** | Kubios | Cohort | 0.695716372 | 0.910114258 |
| **MSE(13)...Nonlinear_Results...Multiscale_entropy** | Kubios | Cohort:Time_of_day | 0.053746836 | 0.204637691 |
| **MSE(14)...Nonlinear_Results...Multiscale_entropy** | Kubios | Time_of_day | 0.049784614 | 0.200448576 |
| **MSE(14)...Nonlinear_Results...Multiscale_entropy** | Kubios | Cohort | 0.752307989 | 0.910114258 |
| **MSE(14)...Nonlinear_Results...Multiscale_entropy** | Kubios | Cohort:Time_of_day | 0.073465959 | 0.219854879 |
| **MSE(15)...Nonlinear_Results...Multiscale_entropy** | Kubios | Time_of_day | 0.055840904 | 0.204637691 |
| **MSE(15)...Nonlinear_Results...Multiscale_entropy** | Kubios | Cohort | 0.757812519 | 0.910114258 |
| **MSE(15)...Nonlinear_Results...Multiscale_entropy** | Kubios | Cohort:Time_of_day | 0.077314951 | 0.219854879 |
| **MSE(16)...Nonlinear_Results...Multiscale_entropy** | Kubios | Time_of_day | 0.086726519 | 0.239083916 |
| **MSE(16)...Nonlinear_Results...Multiscale_entropy** | Kubios | Cohort | 0.716224423 | 0.910114258 |
| **MSE(16)...Nonlinear_Results...Multiscale_entropy** | Kubios | Cohort:Time_of_day | 0.118470925 | 0.290016824 |
| **MSE(17)...Nonlinear_Results...Multiscale_entropy** | Kubios | Time_of_day | 0.038284713 | 0.174852571 |
| **MSE(17)...Nonlinear_Results...Multiscale_entropy** | Kubios | Cohort | 0.708953036 | 0.910114258 |
| **MSE(17)...Nonlinear_Results...Multiscale_entropy** | Kubios | Cohort:Time_of_day | 0.094608323 | 0.249570231 |
| **MSE(18)...Nonlinear_Results...Multiscale_entropy** | Kubios | Time_of_day | 0.064733793 | 0.206324001 |
| **MSE(18)...Nonlinear_Results...Multiscale_entropy** | Kubios | Cohort | 0.764642192 | 0.910114258 |
| **MSE(18)...Nonlinear_Results...Multiscale_entropy** | Kubios | Cohort:Time_of_day | 0.121575551 | 0.295254909 |
| **MSE(19)...Nonlinear_Results...Multiscale_entropy** | Kubios | Time_of_day | 0.049220474 | 0.200448576 |
| **MSE(19)...Nonlinear_Results...Multiscale_entropy** | Kubios | Cohort | 0.684612769 | 0.910114258 |
| **MSE(19)...Nonlinear_Results...Multiscale_entropy** | Kubios | Cohort:Time_of_day | 0.145429109 | 0.337131116 |
| **MSE(2)...Nonlinear_Results...Multiscale_entropy** | Kubios | Time_of_day | 0.222687534 | 0.439628294 |
| **MSE(2)...Nonlinear_Results...Multiscale_entropy** | Kubios | Cohort | 0.991407126 | 0.994657641 |
| **MSE(2)...Nonlinear_Results...Multiscale_entropy** | Kubios | Cohort:Time_of_day | 0.250442511 | 0.478971303 |
| **MSE(20)...Nonlinear_Results...Multiscale_entropy** | Kubios | Time_of_day | 0.048476097 | 0.200448576 |
| **MSE(20)...Nonlinear_Results...Multiscale_entropy** | Kubios | Cohort | 0.745013238 | 0.910114258 |
| **MSE(20)...Nonlinear_Results...Multiscale_entropy** | Kubios | Cohort:Time_of_day | 0.066771525 | 0.206384714 |
| **MSE(3)...Nonlinear_Results...Multiscale_entropy** | Kubios | Time_of_day | 0.10671234 | 0.269867571 |

| | | | | |
|---|---|---|---|---|
| **MSE(3)...Nonlinear_Results...Multiscale_entropy** | Kubios | Cohort | 0.986000694 | 0.992487541 |
| **MSE(3)...Nonlinear_Results...Multiscale_entropy** | Kubios | Cohort:Time_of_day | 0.141518259 | 0.33311221 |
| **MSE(4)...Nonlinear_Results...Multiscale_entropy** | Kubios | Time_of_day | 0.075793194 | 0.219854879 |
| **MSE(4)...Nonlinear_Results...Multiscale_entropy** | Kubios | Cohort | 0.911192306 | 0.945168969 |
| **MSE(4)...Nonlinear_Results...Multiscale_entropy** | Kubios | Cohort:Time_of_day | 0.172211177 | 0.373734895 |
| **MSE(5)...Nonlinear_Results...Multiscale_entropy** | Kubios | Time_of_day | 0.061339337 | 0.204637691 |
| **MSE(5)...Nonlinear_Results...Multiscale_entropy** | Kubios | Cohort | 0.982472606 | 0.992200057 |
| **MSE(5)...Nonlinear_Results...Multiscale_entropy** | Kubios | Cohort:Time_of_day | 0.141453814 | 0.33311221 |
| **MSE(6)...Nonlinear_Results...Multiscale_entropy** | Kubios | Time_of_day | 0.062658333 | 0.204637691 |
| **MSE(6)...Nonlinear_Results...Multiscale_entropy** | Kubios | Cohort | 0.973315822 | 0.989483859 |
| **MSE(6)...Nonlinear_Results...Multiscale_entropy** | Kubios | Cohort:Time_of_day | 0.09602015 | 0.251129624 |
| **MSE(7)...Nonlinear_Results...Multiscale_entropy** | Kubios | Time_of_day | 0.056453694 | 0.204637691 |
| **MSE(7)...Nonlinear_Results...Multiscale_entropy** | Kubios | Cohort | 0.929903984 | 0.961319659 |
| **MSE(7)...Nonlinear_Results...Multiscale_entropy** | Kubios | Cohort:Time_of_day | 0.076696134 | 0.219854879 |
| **MSE(8)...Nonlinear_Results...Multiscale_entropy** | Kubios | Time_of_day | 0.044198844 | 0.185271867 |
| **MSE(8)...Nonlinear_Results...Multiscale_entropy** | Kubios | Cohort | 0.889809666 | 0.932471773 |
| **MSE(8)...Nonlinear_Results...Multiscale_entropy** | Kubios | Cohort:Time_of_day | 0.07500979 | 0.219854879 |
| **MSE(9)...Nonlinear_Results...Multiscale_entropy** | Kubios | Time_of_day | 0.032990996 | 0.162826527 |
| **MSE(9)...Nonlinear_Results...Multiscale_entropy** | Kubios | Cohort | 0.757926367 | 0.910114258 |
| **MSE(9)...Nonlinear_Results...Multiscale_entropy** | Kubios | Cohort:Time_of_day | 0.055209917 | 0.204637691 |
| **NNxx_(beats)...Time_Domain_Results...Statistical_parameters** | Kubios | Time_of_day | 0.097009687 | 0.251567493 |
| **NNxx_(beats)...Time_Domain_Results...Statistical_parameters** | Kubios | Cohort | 0.594412167 | 0.834358363 |
| **NNxx_(beats)...Time_Domain_Results...Statistical_parameters** | Kubios | Cohort:Time_of_day | 0.823393999 | 0.910114258 |
| **pNNxx_(%)...Time_Domain_Results...Statistical_parameters** | Kubios | Time_of_day | 0.01956449 | 0.112957242 |
| **pNNxx_(%)...Time_Domain_Results...Statistical_parameters** | Kubios | Cohort | 0.499315536 | 0.745319775 |
| **pNNxx_(%)...Time_Domain_Results...Statistical_parameters** | Kubios | Cohort:Time_of_day | 0.998789488 | 0.998789488 |
| **Recurrence_rate_(REC)_(%)...Nonlinear_Results...Recurrence_plot_analysis** | Kubios | Time_of_day | 0.378713404 | 0.629816857 |
| **Recurrence_rate_(REC)_(%)...Nonlinear_Results...Recurrence_plot_analysis** | Kubios | Cohort | 0.765802974 | 0.910114258 |
| **Recurrence_rate_(REC)_(%)...Nonlinear_Results...Recurrence_plot_analysis** | Kubios | Cohort:Time_of_day | 0.801349103 | 0.910114258 |
| **RMSSD_(ms)...Time_Domain_Results...Statistical_parameters** | Kubios | Time_of_day | 0.065448583 | 0.206324001 |
| **RMSSD_(ms)...Time_Domain_Results...Statistical_parameters** | Kubios | Cohort | 0.625026862 | 0.857663783 |
| **RMSSD_(ms)...Time_Domain_Results...Statistical_parameters** | Kubios | Cohort:Time_of_day | 0.834752354 | 0.910114258 |
| **RR_tri_index...Time_Domain_Results...Geometric_parameters** | Kubios | Time_of_day | 0.005523364 | 0.041223154 |
| **RR_tri_index...Time_Domain_Results...Geometric_parameters** | Kubios | Cohort | 0.457179662 | 0.707255193 |
| **RR_tri_index...Time_Domain_Results...Geometric_parameters** | Kubios | Cohort:Time_of_day | 0.20609887 | 0.420441695 |
| **Sample_entropy_(SampEn)...Nonlinear_Results** | Kubios | Time_of_day | 0.012845344 | 0.084653037 |
| **Sample_entropy_(SampEn)...Nonlinear_Results** | Kubios | Cohort | 0.645926433 | 0.878459949 |
| **Sample_entropy_(SampEn)...Nonlinear_Results** | Kubios | Cohort:Time_of_day | 0.496545758 | 0.744818636 |
| **SD1_(ms)...Nonlinear_Results...Poincare_plot** | Kubios | Time_of_day | 0.065430889 | 0.206324001 |
| **SD1_(ms)...Nonlinear_Results...Poincare_plot** | Kubios | Cohort | 0.625029489 | 0.857663783 |
| **SD1_(ms)...Nonlinear_Results...Poincare_plot** | Kubios | Cohort:Time_of_day | 0.834764461 | 0.910114258 |
| **SD2/SD1_ratio...Nonlinear_Results...Poincare_plot** | Kubios | Time_of_day | 0.036481206 | 0.171742294 |
| **SD2/SD1_ratio...Nonlinear_Results...Poincare_plot** | Kubios | Cohort | 0.854118221 | 0.910133522 |

| | | | | |
|---|---|---|---|---|
| **SD2/SD1_ratio...Nonlinear_Results...Poincare_plot** | Kubios | Cohort:Time_of_day | 0.593277859 | 0.834358363 |
| **SD2_(ms)...Nonlinear_Results...Poincare_plot** | Kubios | Time_of_day | 0.007001814 | 0.04982686 |
| **SD2_(ms)...Nonlinear_Results...Poincare_plot** | Kubios | Cohort | 0.315687405 | 0.560564645 |
| **SD2_(ms)...Nonlinear_Results...Poincare_plot** | Kubios | Cohort:Time_of_day | 0.198475014 | 0.413918475 |
| **SDANN_(ms)...Time_Domain_Results...Statistical_parameters** | Kubios | Time_of_day | 0.414213634 | 0.667101958 |
| **SDANN_(ms)...Time_Domain_Results...Statistical_parameters** | Kubios | Cohort | 0.481988228 | 0.733773124 |
| **SDANN_(ms)...Time_Domain_Results...Statistical_parameters** | Kubios | Cohort:Time_of_day | 0.956723075 | 0.975857536 |
| **SDNN_index_(ms)...Time_Domain_Results...Statistical_parameters** | Kubios | Time_of_day | 0.171529311 | 0.373734895 |
| **SDNN_index_(ms)...Time_Domain_Results...Statistical_parameters** | Kubios | Cohort | 0.162335173 | 0.360377253 |
| **SDNN_index_(ms)...Time_Domain_Results...Statistical_parameters** | Kubios | Cohort:Time_of_day | 0.703639637 | 0.910114258 |
| **Shannon_entropy...Nonlinear_Results...Recurrence_plot_analysis** | Kubios | Time_of_day | 0.785467015 | 0.910114258 |
| **Shannon_entropy...Nonlinear_Results...Recurrence_plot_analysis** | Kubios | Cohort | 0.886486403 | 0.932181579 |
| **Shannon_entropy...Nonlinear_Results...Recurrence_plot_analysis** | Kubios | Cohort:Time_of_day | 0.409615632 | 0.666714805 |
| **STD_HR_(beats/min)...Time_Domain_Results...Statistical_parameters** | Kubios | Time_of_day | 0.298569679 | 0.543823343 |
| **STD_HR_(beats/min)...Time_Domain_Results...Statistical_parameters** | Kubios | Cohort | 0.017879299 | 0.107275792 |
| **STD_HR_(beats/min)...Time_Domain_Results...Statistical_parameters** | Kubios | Cohort:Time_of_day | 0.253710223 | 0.482207007 |
| **STD_RR_(ms)...Time_Domain_Results...Statistical_parameters** | Kubios | Time_of_day | 0.019528123 | 0.112957242 |
| **STD_RR_(ms)...Time_Domain_Results...Statistical_parameters** | Kubios | Cohort | 0.45072229 | 0.707255193 |
| **STD_RR_(ms)...Time_Domain_Results...Statistical_parameters** | Kubios | Cohort:Time_of_day | 0.450602384 | 0.707255193 |
| **TINN_(ms)...Time_Domain_Results...Geometric_parameters** | Kubios | Time_of_day | 0.043777458 | 0.185271867 |
| **TINN_(ms)...Time_Domain_Results...Geometric_parameters** | Kubios | Cohort | 0.809445359 | 0.910114258 |
| **TINN_(ms)...Time_Domain_Results...Geometric_parameters** | Kubios | Cohort:Time_of_day | 0.371574795 | 0.627476888 |
| **Total_power_(ms^2)...Frequency_Domain_Results...AR_spectrum** | Kubios | Time_of_day | 0.289707642 | 0.530841548 |
| **Total_power_(ms^2)...Frequency_Domain_Results...AR_spectrum** | Kubios | Cohort | 0.749435516 | 0.910114258 |
| **Total_power_(ms^2)...Frequency_Domain_Results...AR_spectrum** | Kubios | Cohort:Time_of_day | 0.665714614 | 0.89739503 |
| **Total_power_(ms^2)...Frequency_Domain_Results...FFT_spectrum** | Kubios | Time_of_day | 0.190268569 | 0.404320709 |
| **Total_power_(ms^2)...Frequency_Domain_Results...FFT_spectrum** | Kubios | Cohort | 0.79435772 | 0.910114258 |
| **Total_power_(ms^2)...Frequency_Domain_Results...FFT_spectrum** | Kubios | Cohort:Time_of_day | 0.642068317 | 0.877111183 |
| **VLF_(%)...Frequency_Domain_Results...Relative_powers...AR_spectrum** | Kubios | Time_of_day | 0.325567652 | 0.562845771 |
| **VLF_(%)...Frequency_Domain_Results...Relative_powers...AR_spectrum** | Kubios | Cohort | 0.0269123 | 0.141985585 |
| **VLF_(%)...Frequency_Domain_Results...Relative_powers...AR_spectrum** | Kubios | Cohort:Time_of_day | 0.862160855 | 0.911501431 |
| **VLF_(%)...Frequency_Domain_Results...Relative_powers...FFT_spectrum** | Kubios | Time_of_day | 0.605451592 | 0.842128123 |
| **VLF_(%)...Frequency_Domain_Results...Relative_powers...FFT_spectrum** | Kubios | Cohort | 0.006189736 | 0.045096647 |
| **VLF_(%)...Frequency_Domain_Results...Relative_powers...FFT_spectrum** | Kubios | Cohort:Time_of_day | 0.949793395 | 0.972029361 |
| **VLF_(Hz)...Frequency_Domain_Results...Peak_frequencies...AR_spectrum** | Kubios | Time_of_day | 0.105911029 | 0.269867571 |
| **VLF_(Hz)...Frequency_Domain_Results...Peak_frequencies...AR_spectrum** | Kubios | Cohort | 0.092601467 | 0.248561834 |
| **VLF_(Hz)...Frequency_Domain_Results...Peak_frequencies...AR_spectrum** | Kubios | Cohort:Time_of_day | 0.212983678 | 0.428769773 |
| **VLF_(Hz)...Frequency_Domain_Results...Peak_frequencies...FFT_spectrum** | Kubios | Time_of_day | 0.094445539 | 0.249570231 |
| **VLF_(Hz)...Frequency_Domain_Results...Peak_frequencies...FFT_spectrum** | Kubios | Cohort | 0.217490914 | 0.432639583 |
| **VLF_(Hz)...Frequency_Domain_Results...Peak_frequencies...FFT_spectrum** | Kubios | Cohort:Time_of_day | 0.727596374 | 0.910114258 |
| **VLF_(log)...Frequency_Domain_Results...Absolute_powers...AR_spectrum** | Kubios | Time_of_day | 0.000978751 | 0.009661216 |
| **VLF_(log)...Frequency_Domain_Results...Absolute_powers...AR_spectrum** | Kubios | Cohort | 0.059246386 | 0.204637691 |
| **VLF_(log)...Frequency_Domain_Results...Absolute_powers...AR_spectrum** | Kubios | Cohort:Time_of_day | 0.287765459 | 0.53045922 |

| | | | | |
|---|---|---|---|---|
| **VLF_(log)...Frequency_Domain_Results...Absolute_powers...FFT_spectrum** | Kubios | Time_of_day | 0.001586824 | 0.013488002 |
| **VLF_(log)...Frequency_Domain_Results...Absolute_powers...FFT_spectrum** | Kubios | Cohort | 0.070878689 | 0.216888788 |
| **VLF_(log)...Frequency_Domain_Results...Absolute_powers...FFT_spectrum** | Kubios | Cohort:Time_of_day | 0.22487298 | 0.441096999 |
| **VLF_(ms^2)...Frequency_Domain_Results...Absolute_powers...AR_spectrum** | Kubios | Time_of_day | 0.810538891 | 0.910114258 |
| **VLF_(ms^2)...Frequency_Domain_Results...Absolute_powers...AR_spectrum** | Kubios | Cohort | 0.150107891 | 0.343370636 |
| **VLF_(ms^2)...Frequency_Domain_Results...Absolute_powers...AR_spectrum** | Kubios | Cohort:Time_of_day | 0.323307609 | 0.562845771 |
| **VLF_(ms^2)...Frequency_Domain_Results...Absolute_powers...FFT_spectrum** | Kubios | Time_of_day | 0.316068105 | 0.560564645 |
| **VLF_(ms^2)...Frequency_Domain_Results...Absolute_powers...FFT_spectrum** | Kubios | Cohort | 0.306374432 | 0.551473978 |
| **VLF_(ms^2)...Frequency_Domain_Results...Absolute_powers...FFT_spectrum** | Kubios | Cohort:Time_of_day | 0.35139206 | 0.604078485 |

#### Table S 9. Time-versus-subject contribution to variance analysis

| Diagnosis | Data.type | Time [%] | Subject [%] |
|---|---|---|---|
| **CKD/T2DM** | Activity | 6.5 | 2.7 |
| **CKD/T2DM** | BR | 1.0 | 7.4 |
| **CKD/T2DM** | HR | 7.0 | 19.3 |
| **CKD/T2DM** | HRV | 1.8 | 49.5 |
| **CKD** | Activity | 5.0 | 1.3 |
| **CKD** | BR | 3.0 | 6.2 |
| **CKD** | HR | 5.7 | 10.2 |
| **CKD** | HRV | 2.8 | 36.1 |
| **Control** | Activity | 11.5 | 2.5 |
| **Control** | BR | 6.5 | 12.2 |
| **Control** | HR | 17.4 | 10.3 |
| **Control** | HRV | 3.8 | 19.2 |

**Table S 10. Cosinor versus LOESS derived phase and amplitude of activity, breath/heart rate and HRV-SDNN**

| Measurement | Cohort | Cosinor Phase | LOESS Peak Time | Cosinor Amplitude | LOESS Amplitude |
|---|---|---|---|---|---|
| **Unit** | | Decimal hour | Decimal hour | Decimal hour | Decimal hour |
| **Activity** | **CKD** | 14.8±1.4 | 14.2±3.1 | 0.02±0.01 | 0.05±0.01 |
| **Activity** | **CKD/T2DM** | 14.2±1.3 | 13.8±2.5 | 0.02±0 | 0.04±0.01 |
| **Activity** | **Healthy control** | 14.9±1.5 | 14±3.5 | 0.03±0.01 | 0.09±0.03 |
| **BR** | **CKD** | 14.8±4.9 | 15.4±5.2 | 2.6±2.9 | 5.7±5.9 |
| **BR** | **CKD/T2DM** | 14±5.8 | 10.4±6.7 | 1.3±0.8 | 3.1±1.6 |
| **BR** | **Healthy control** | 15.9±1.6 | 14.6±3.6 | 1.8±0.7 | 4.6±1.9 |
| **HR** | **CKD** | 14.9±4.2 | 14.2±4.3 | 14.5±7.1 | 32.6±12.5 |
| **HR** | **CKD/T2DM** | 15.2±3.5 | 13.9±5 | 15.4±9.6 | 34.3±17.5 |
| **HR** | **Healthy control** | 15.4±1.6 | 16±2.9 | 15.9±5.8 | 37.2±16 |
| **HRV-SDNN** | **CKD** | 6.6±3.1 | 6.1±3.1 | 9.4±7.4 | 22.9±16.2 |
| **HRV-SDNN** | **CKD/T2DM** | 11.2±6.3 | 11±5.9 | 8.4±6.6 | 20.1±14.7 |
| **HRV-SDNN** | **Healthy control** | 8.1±2.9 | 8.6±3.5 | 8.7±3.3 | 20.8±8.1 |

**Table S 11. Cosinor summary statistics**

| Measurement | Stat | Diagnosis | Mean | SD | Min | Max |
|---|---|---|---|---|---|---|
| **Activity** | Amplitude | Control | 0.032933 | 0.010551 | 0.01999 | 0.054076 |
| **Activity** | Amplitude | CKD/T2DM | 0.022108 | 0.007288 | 0.004486 | 0.039199 |
| **Activity** | Amplitude | CKD | 0.023348 | 0.010424 | 0.009618 | 0.048383 |
| **Activity** | MESOR | Control | 0.057242 | 0.012264 | 0.043323 | 0.076702 |
| **Activity** | MESOR | CKD/T2DM | 0.038199 | 0.007767 | 0.02458 | 0.054018 |
| **Activity** | MESOR | CKD | 0.04115 | 0.006114 | 0.035814 | 0.055244 |
| **Activity** | Phase | Control | 14.94188 | 1.485604 | 12.83845 | 18.3043 |
| **Activity** | Phase | CKD/T2DM | 14.29866 | 1.326464 | 11.59713 | 16.68756 |
| **Activity** | Phase | CKD | 14.84831 | 1.379352 | 11.7978 | 17.49499 |
| **BR** | Amplitude | Control | 1.897828 | 0.728783 | 0.307239 | 3.075083 |
| **BR** | Amplitude | CKD/T2DM | 1.307728 | 0.871598 | 0.107151 | 3.206893 |
| **BR** | Amplitude | CKD | 2.620348 | 2.901209 | 0.141466 | 12.85332 |
| **BR** | MESOR | Control | 14.07311 | 1.685208 | 11.30725 | 16.35684 |
| **BR** | MESOR | CKD/T2DM | 13.45193 | 1.484155 | 10.19805 | 15.41471 |
| **BR** | MESOR | CKD | 14.13405 | 1.891871 | 11.01114 | 17.29843 |
| **BR** | Phase | Control | 16.00052 | 1.580133 | 12.96504 | 17.92036 |
| **BR** | Phase | CKD/T2DM | 16.30554 | 5.387609 | 0.905382 | 23.42762 |
| **BR** | Phase | CKD | 16.46192 | 3.893574 | 1.55436 | 22.08823 |
| **HR** | Amplitude | Control | 15.95094 | 5.896111 | 9.281105 | 27.28443 |
| **HR** | Amplitude | CKD/T2DM | 15.49539 | 9.648755 | 3.344289 | 43.26601 |
| **HR** | Amplitude | CKD | 14.56778 | 7.150251 | 2.83839 | 27.56381 |
| **HR** | MESOR | Control | 67.05452 | 10.55413 | 49.00797 | 84.10365 |
| **HR** | MESOR | CKD/T2DM | 66.79328 | 15.23166 | 35.01084 | 88.15576 |
| **HR** | MESOR | CKD | 59.42396 | 10.03368 | 31.36897 | 70.58682 |
| **HR** | Phase | Control | 15.38548 | 1.526589 | 13.37371 | 18.92673 |
| **HR** | Phase | CKD/T2DM | 15.42187 | 3.241405 | 5.571365 | 22.9212 |
| **HR** | Phase | CKD | 15.62654 | 3.862089 | 3.617431 | 21.92535 |
| **HRV** | Amplitude | Control | 8.714579 | 3.389514 | 2.548679 | 13.12954 |
| **HRV** | Amplitude | CKD/T2DM | 8.491701 | 6.694675 | 1.123308 | 24.04616 |
| **HRV** | Amplitude | CKD | 9.46413 | 7.403547 | 1.398102 | 27.14131 |
| **HRV** | MESOR | Control | 62.53539 | 14.07859 | 41.67722 | 85.63802 |
| **HRV** | MESOR | CKD/T2DM | 38.44986 | 23.56721 | 9.572568 | 117.9782 |
| **HRV** | MESOR | CKD | 47.70461 | 19.39366 | 15.70544 | 82.20842 |
| **HRV** | Phase | Control | 7.94997 | 2.860183 | 5.202564 | 13.78725 |
| **HRV** | Phase | CKD/T2DM | 8.572798 | 5.746595 | 1.048827 | 23.62322 |
| **HRV** | Phase | CKD | 6.659687 | 3.196528 | 2.185542 | 11.31282 |

**Table S 12. Cosinor metrics: ANOVA & post-hoc tests**

| Measurement | Metric | term | df | sumsq | meansq | statistic | p.value | Holm.Adj.p_value |
|---|---|---|---|---|---|---|---|---|
| **Activity** | MESOR | Diagnosis | 2 | 0.0025 | 0.0013 | 18.0567 | 1.88E-06 | 1.51E-05 |
| **BR** | MESOR | Diagnosis | 2 | 5.0213 | 2.5107 | 0.8860 | 0.420 | 1 |
| **HR** | MESOR | Diagnosis | 2 | 603.75 | 301.88 | 1.8917 | 0.163 | 0.651 |
| **HRV** | MESOR | Diagnosis | 2 | 3883.73 | 1941.87 | 4.6551 | 0.015 | 0.088 |
| **Activity** | amp | Diagnosis | 2 | 0.0008 | 0.0004 | 4.9400 | 0.012 | 0.081 |
| **BR** | amp | Diagnosis | 2 | 15.8340 | 7.9170 | 2.2637 | 0.116 | 0.580 |
| **HR** | amp | Diagnosis | 2 | 14.0275 | 7.0138 | 0.1064 | 0.899 | 1 |
| **HRV** | amp | Diagnosis | 2 | 9.0842 | 4.5421 | 0.1091 | 0.897 | 1 |

| Measurement | Metric | term | comparison | estimate | conf.low | conf.high | adj.p.value |
|---|---|---|---|---|---|---|---|
| **Activity** | MESOR | Diagnosis | Normoglycemic-Diabetic | 0.003 | -0.004 | 0.010 | 0.540 |
| **Activity** | MESOR | Diagnosis | Control-Diabetic | 0.019 | 0.011 | 0.027 | 1.61E-06 |
| **Activity** | MESOR | Diagnosis | Control-Normoglycemic | 0.016 | 0.008 | 0.024 | 5.24E-05 |
| **BR** | MESOR | Diagnosis | Normoglycemic-Diabetic | 0.682 | -0.665 | 2.029 | 0.443 |
| **BR** | MESOR | Diagnosis | Control-Diabetic | 0.621 | -0.960 | 2.202 | 0.610 |
| **BR** | MESOR | Diagnosis | Control-Normoglycemic | -0.061 | -1.688 | 1.566 | 0.995 |
| **HR** | MESOR | Diagnosis | Normoglycemic-Diabetic | -7.369 | -17.477 | 2.738 | 0.192 |
| **HR** | MESOR | Diagnosis | Control-Diabetic | 0.261 | -11.605 | 12.128 | 0.998 |
| **HR** | MESOR | Diagnosis | Control-Normoglycemic | 7.631 | -4.580 | 19.841 | 0.293 |
| **HRV** | MESOR | Diagnosis | Normoglycemic-Diabetic | 9.255 | -7.087 | 25.597 | 0.363 |
| **HRV** | MESOR | Diagnosis | Control-Diabetic | 24.086 | 4.899 | 43.272 | 0.011 |
| **HRV** | MESOR | Diagnosis | Control-Normoglycemic | 14.831 | -4.912 | 34.573 | 0.174 |
| **Activity** | Amplitude | Diagnosis | Normoglycemic-Diabetic | 0.001 | -0.006 | 0.009 | 0.913 |
| **Activity** | Amplitude | Diagnosis | Control-Diabetic | 0.011 | 0.002 | 0.019 | 0.011 |
| **Activity** | Amplitude | Diagnosis | Control-Normoglycemic | 0.010 | 0.001 | 0.019 | 0.033 |
| **BR** | Amplitude | Diagnosis | Normoglycemic-Diabetic | 1.313 | -0.184 | 2.809 | 0.096 |
| **BR** | Amplitude | Diagnosis | Control-Diabetic | 0.590 | -1.167 | 2.347 | 0.696 |
| **BR** | Amplitude | Diagnosis | Control-Normoglycemic | -0.723 | -2.530 | 1.085 | 0.600 |
| **HR** | Amplitude | Diagnosis | Normoglycemic-Diabetic | -0.928 | -7.423 | 5.568 | 0.936 |
| **HR** | Amplitude | Diagnosis | Control-Diabetic | 0.456 | -7.170 | 8.082 | 0.988 |
| **HR** | Amplitude | Diagnosis | Control-Normoglycemic | 1.383 | -6.464 | 9.230 | 0.904 |
| **HRV** | Amplitude | Diagnosis | Normoglycemic-Diabetic | 0.972 | -4.190 | 6.135 | 0.892 |
| **HRV** | Amplitude | Diagnosis | Control-Diabetic | 0.223 | -5.839 | 6.284 | 0.996 |
| **HRV** | Amplitude | Diagnosis | Control-Normoglycemic | -0.750 | -6.987 | 5.488 | 0.954 |
